# Malaria transmission modelling and conditional prevention allocation in Uganda

**DOI:** 10.64898/2026.10.23.26365767

**Authors:** Jay Hiteshkumar Patel, John Mulangwa, Milton Musaba

**Affiliations:** Butisema University, Kampala, Uganda

**Keywords:** malaria transmission, structural identifiability, synthetic calibration, prevention allocation, R21/Matrix-M, Uganda, reproducibility

## Abstract

Malaria transmission models can generate precise intervention rankings even when their parameters, observation mappings and programme costs remain uncertain. I evaluated an age-stratified SEIR–SEI framework in a standardised population of one million, using an unchanged 36-month historical synthetic target. The reconciled implementation maintained virtual age-group totals, fitted the transmission composite K = β₀M, integrated infectious onsets and disease deaths, and compared interventions from one phase-aligned baseline state over 720 days. An aggregate vaccine effect entered child infection before simulation, allowing transmission feedback.

Five calibration seeds gave total-series R² 0.9537; subgroup fits were weaker. The closed-form unforced reproduction index, 6.3787, matched the six-state next-generation matrix. The baseline produced 528,041 expected reported episodes. ITN coverage of 50% averted 36,692 episodes; vaccine coverage of 70% averted 2,123 under the assumed aggregate mechanism. At a hypothetical US$2 million additional prevention budget, five allocation seeds and a 10,201-point boundary grid selected 71.43% ITN coverage, averting 52,482.83 reported episodes. The same corner won within a 115-portfolio pool for all 300 assumed parameter draws and remained selected after 40 repeated noisy-target recalibrations. Halving the IRS round price and using two rounds changed the selected mix to 40.07% ITN and 21.95% IRS. All 17 mathematical and numerical assertions passed.

The study demonstrates reproducible conditional calculations and identifies important structural confounding. It does not establish empirical Uganda programme effectiveness, a clinical treatment allocation or a national cost-effectiveness recommendation. Empirical use requires compatible surveillance observations, a cohort-based vaccine mechanism, treatment-specific clinical effects and locally measured programme resources.

## 1 Introduction

Uganda’s malaria burden varies across settings and surveillance systems [1,2,3]. Transmission models can help explain the interaction between human infection, mosquito survival and prevention, but programme decisions also depend on the meaning of observed cases, access to treatment and the resources needed to deliver each intervention. A high correlation between model output and a case series cannot resolve these questions by itself.

Uganda introduced R21/Matrix-M into routine immunisation on 2 April 2025, initially targeting approximately 1.1 million children in 105 high- and moderate-transmission districts. The four doses were scheduled at 6, 7, 8 and 18 months [4]. The phase 3 trial reported 12-month efficacy against a first clinical malaria episode of 75% at seasonal sites and 68% at standard sites [5]. These findings motivate study of combined prevention. They do not supply a direct calibration of infection hazards, transmission feedback or completed-course coverage in a simplified population model.

Established malaria models provide frameworks for infection, immunity and intervention analysis [6,7]. This study examines a compact model in which the mathematical structure, observation rule and budget calculation can be inspected together. Its contribution is a reconciled computational workflow and an explicit account of what each diagnostic establishes. No comparison of scientific performance or operational usability with EMOD, OpenMalaria or another modelling platform was undertaken.

### 1.1 Research questions

Four questions organise the analysis. First, can monthly human case counts identify transmission intensity separately from the mosquito-density scale? Second, how does changing recovery affect episode cycling when temporary immunity and infectious-exit fatality are held in a specified relationship? Third, do separate child exposure and susceptibility labels represent distinguishable mechanisms in these equations? Fourth, which additional prevention portfolios are favoured under a declared budget, and how do that answer and its effect size change under parameter, observation and structural uncertainty?

The first three questions concern the mathematical system. The fourth concerns a conditional budget illustration with explicit programme inputs. They are assessed with algebraic identities, common-state simulations, repeated searches and sensitivity exercises. The analysis does not treat literature compatibility, in-sample synthetic fit or numerical correctness as a substitute for empirical prediction assessment [8,9].

### 1.2 Study status

This is a retrospective computational study. Historical model development and synthetic observations preceded the protocol; the present numerical reference was reconciled and executed on 11 October 2026 under protocol version 0.5. The analyses were specified after earlier data and results had been inspected. No prospective registration or independently validated national policy model is claimed. This paper reports the matching reference calculations, rather than assigning the older manuscript’s results to amended equations.

## 2 Methods

### 2.1 Population model and data

The model represents one homogeneous transmission zone with *N*_h_ = 1,000,000. The fixed shares are *π*_c_ = 0.17 for children under five and *π*_a_ = 0.83 for people aged five and over. The latter group includes school-age children. Each group contains susceptible, exposed, infectious and recovered states, and both share one susceptible–exposed–infectious mosquito population. These maintained virtual populations have no individual ageing process. They standardise the computational illustration rather than reconstructing Uganda’s demographic structure.

The unchanged uganda_synthetic_monthly.csv contains 36 calendar months from January 2021 to December 2023. The fitted fields are reported_cases_children and reported_cases_adults; the adult-labelled field is interpreted as people aged five and over. The target is synthetic. The historical generating implementation counted infection acquisitions at step boundaries and differed in demographic and reporting conventions. Consequently, fitting the current infectious-onset model to that fixture assesses compatibility with the historical series; it cannot be interpreted as recovery of its original generating parameters.

There is no imputation because all 36 target rows are complete. The fitting objective and scenario outputs are deterministic. Observation randomness is contained in the historical fixture or generated as a separate monthly target in the repeated-noise experiment. A separate noiseless fixture generated by the current equations is used for a same-model recovery check. Appendix A gives the full reference parameter dictionary.

**Figure 1.**
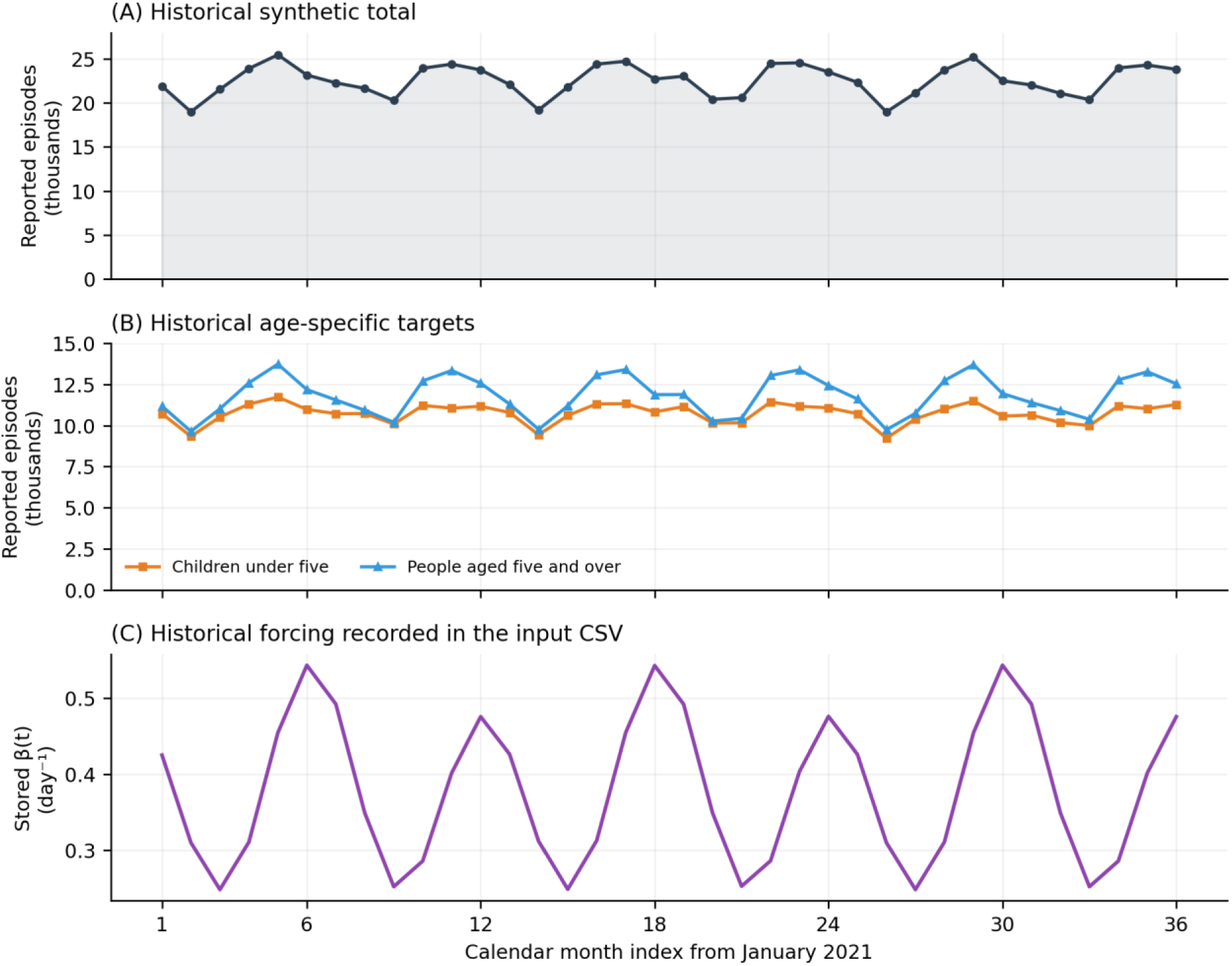
Historical synthetic dataset profile. Panels A and B show the 36 reported-count targets. Panel C uses beta_t exactly as recorded in the historical CSV; it is separate from the fitted forcing and contains no measured rainfall data.

### 2.2 Observation and outcome definitions

The fitted reporting input ρ is a nominal population-share average. A fixed child-to-older reporting ratio of 1.51 determines the age probabilities for every parameter vector through Equation 1. The probabilities are propagated during calibration and scenarios, rather than retained as fixed historical constants when ρ changes. They are model mappings and are not independent estimates of ascertainment.

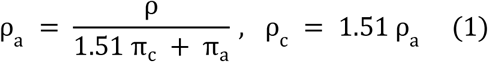

The primary outcome is expected model-reported episode onset, Y. An episode begins at progression from E to I; it is not an infection acquisition, a unique infected individual, a laboratory-confirmed consultation or a severe admission. A person may contribute several episodes during the horizon. Secondary outcomes are onset totals before reporting, age-specific outputs and integrated ODE disease deaths. Clinical severity, asymptomatic infection and diagnostic testing are absent.

### 2.3 Transmission and population dynamics

Time is measured in days. Human and vector states are represented internally as fractions of *N*_h_, and output is returned as counts. Human infection depends on infectious mosquitoes through Equation 2. The child multiplier *q*_c_ = 2.31 is the product of the historical exposure and susceptibility labels. Mosquito infection depends equally on the two human infectious groups through Equation 3. β₀ and *β*_m_ are effective contact–transmission coefficients; no biting-frequency or infection-probability decomposition is inferred.

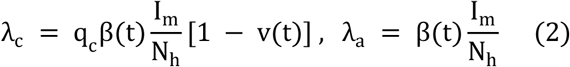

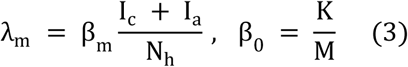

**Figure 2.**
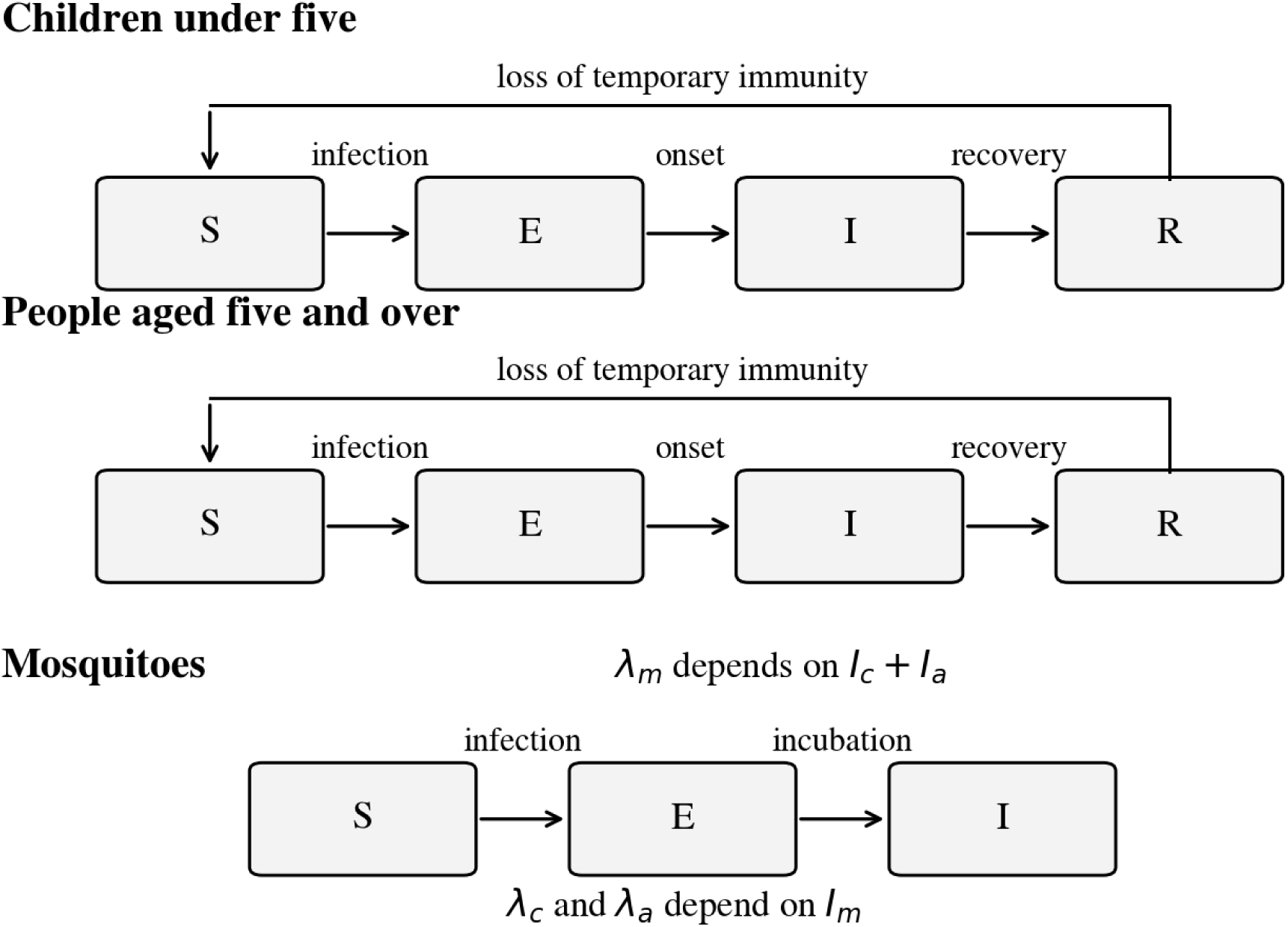
Human SEIR and mosquito SEI structure. Both human groups share the vector pool through Equations 2–3. Recovery, temporary immunity, recruitment and mortality follow Equations 5–7. Vaccine protection modifies child infection before integration.

Seasonality is an assumed 365-day forcing shape with fitted amplitude A and phase φ. The annual and semiannual weights, 0.20 and 0.80, the day-115 anchor and the 0.10 floor are fixed. No rainfall data enter the model, and neither a biological rainfall lag nor district geography is estimated by fitting this curve.

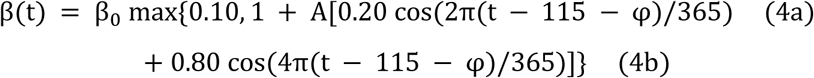

For each human group g, recruitment replaces natural and disease deaths as in Equation 5, preserving *N*_g_ = *π*_g_*N*_h_. Recruitment into the older group is a mathematical replacement flow. CFR_g_ is the assumed probability of disease death among competing exits from *I*_g_. Its hazard includes recovery and natural mortality so that the corresponding probability is exactly CFR_g_. The empirical clinical denominator for these assumed probabilities has not been established.

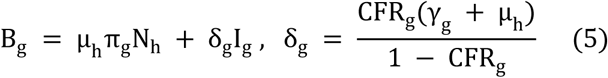

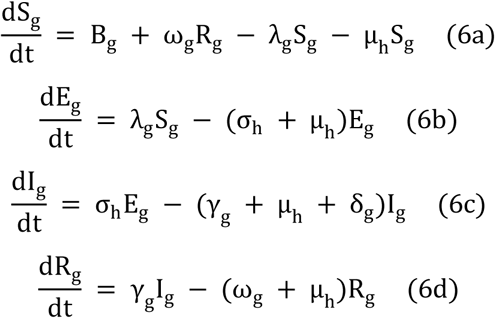

Vector recruitment is *μ*_m,eff_ M *N*_h_. Thus total mosquito density remains M *N*_h_ when survival changes. This convention isolates survival effects without a vector ecology model. It does not estimate a field reduction in mosquito abundance.

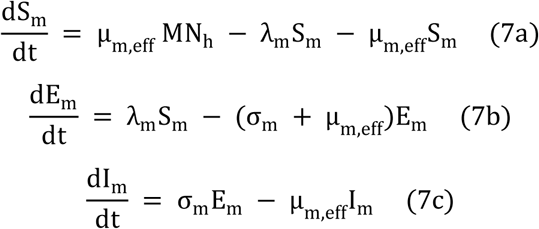

### 2.4 Intervention functions

ITN and IRS coverage fractions act through separate assumed saturating exponential kill probabilities. Their maximum per-cycle probabilities are 0.50 and 0.60; both reference saturation constants are 1.33. Combining their survival probabilities gives the additional daily mortality in Equation 9, with a three-day gonotrophic cycle. These are illustrative response functions. The observational results of Katureebe and colleagues contain site-specific ownership, use and disease measures and are not treated as a fitted coverage-to-kill curve [10].

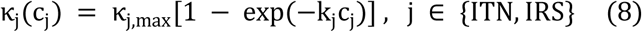

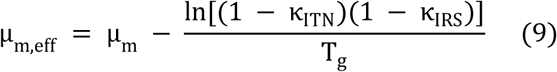

The recovery-only diagnostic interpolates baseline recovery towards a seven-day mean duration (Equation 10). The fatality hazard is recomputed through Equation 5, leaving infectious-exit fatality fixed. This diagnostic does not reuse the vector saturation constant. Treatment-specific reductions in severity, admission and fatality, eligibility for therapy and treatment resources are not represented. Recovery enhancement is therefore excluded from the prevention allocation problem.

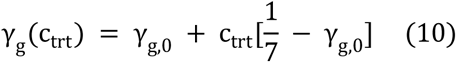

Vaccine coverage refers to completed courses in a fixed eligible share of 40% of children under five. An aggregate protection factor ramps linearly over 90 days and enters *λ*_c_ before integration (Equation 11). Its reference efficacy input is 0.610, calculated as 0.75 × 0.68 + 0.25 × 0.40. The weights and floor are assumptions. The 0.68 clinical-efficacy result supplies context [5], but using it in an infection-hazard modification requires an additional modelling assumption. The trial did not estimate this weighted average or the model’s eligible stock.

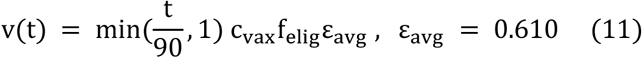

At 70% coverage the plateau child force-of-infection reduction is 17.08%. Reduced child infection can alter mosquito infection and subsequently affect both age groups. This is a mean-field leaky-protection approximation across the child group. It has no vaccinated compartment, dose completion process, births into eligibility, individual waning or booster trajectory [11]. The realised episode reduction is determined by the evolving susceptible and immune pools and is not set equal to 17.08%. There is no post-simulation multiplier on cases or separate vaccine branch that resets caller parameters.

### 2.5 Reproduction and seasonal invasion

The unforced disease-free index R₀ is the spectral radius of FV⁻¹ for infected-state order (*E*_c_,*I*_c_,*E*_a_,*I*_a_,*E*_m_,*I*_m_) [12,13]. For baseline or fixed vaccine plateau protection, *q*_c_* = *q*_c_(1 − *c*_vax_ *f*_elig_ *ε*_avg_). Equations 12a–b give the shared-vector closed form. The age cycle contributions are added inside the square root; an arithmetic demographic average of group-specific roots does not equal this index.

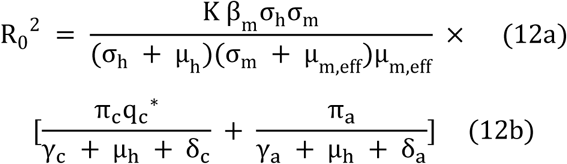

The seasonal invasion diagnostic integrates the six-state linearised system over one 365-day cycle and computes its dominant Floquet multiplier. The logarithm divided by 365 gives a daily growth rate. This is a linear-invasion quantity, not a periodic R₀ or a disease burden forecast. For the vaccine diagnostic, the protection is held at its plateau because the initial 90-day rollout is not a periodic regime.

### 2.6 Numerical integration and comparison horizon

Model onsets and deaths are integrated alongside the compartments at all Runge–Kutta stages, as in Equations 13–14. For constant CFR_g_, integrated deaths satisfy *D*_g_ = CFR_g_[*P*_g_ − {*I*_g_(T) − *I*_g_(0)}]. The terminal infectious stock accounts for the difference between this finite-horizon death flow and CFR_g_ times the total number of onsets.

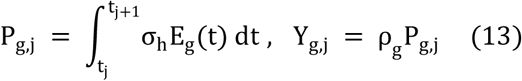

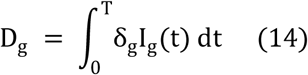

The solver is classical fourth-order Runge–Kutta with a maximum 0.5-day step. The final step is shortened to hit each observation boundary exactly. Calibration uses actual calendar-month boundaries over 1,095 days. Intervention comparisons use 24 consecutive 30-day intervals, totalling 720 days; this is not two calendar years. No stochastic perturbation is inserted into the scenario solver, and calibration and final evaluation use the same numerical operator.

Baseline spin-up iterates phase-aligned 365-day cycles with zero additional intervention. Convergence requires two successive maximum annual changes in normalised states below 10⁻⁸; a failure within 40 cycles raises an error. Each intervention starts from the same baseline state and phase at day zero, rather than its own intervention-specific equilibrium. The counterfactual concerns additional coverage under these background recovery and transmission assumptions. It does not represent withdrawal of existing Uganda services.

### 2.7 Calibration and identifiability

The fitted vector is θ = (K,A,φ,ρ), with bounds 0.30–3.00, 0–0.90, 0–120 days and 0.05–0.60. Equation 15 combines total normalised mean squared error, total Pearson correlation and child normalised mean squared error. NMSE divides squared error by the target-series variance. Older-group diagnostics are reported separately. The weights are retrospective computational choices; this criterion is not an observation likelihood.

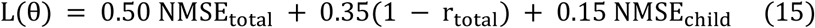

Differential evolution used best1bin, population size 12 per fitted dimension, mutation 0.5–1.0, recombination 0.7, tolerance 10⁻⁷, up to 160 generations and local polish [14]. Five independent runs used seeds 42, 1, 7, 123 and 2024. The minimum-loss solution supplied the reference parameter vector. Every evaluation used the same reporting derivation, spin-up and calendar intervals.

Writing vector states as fractions of M *N*_h_ removes M from their fractional equations and leaves the human-transmission product β₀M = K. M is fixed at 3.5 solely as a scale. A matched rescaling of vector states and β₀ leaves human output unchanged. Similarly, historical *α*_c_ and *φ*_c_ enter only through their product *q*_c_. These symmetries are checked explicitly. Nine-point profiles fixed one fitted input and reoptimised the other three. They describe conditional composite-loss curvature and are not likelihood-ratio confidence intervals [15].

A separate noiseless fixture used generating (K,A,φ,ρ) = (1.33,0.45,55,0.20) under the reconciled equations. Fitting this fixture assessed implementation recovery. It did not test recovery of biological field parameters or compensate for the historical target’s different generating system.

### 2.8 Prevention scenarios and programme costs

The prevention scenarios included ITN at 50% and 80%, IRS at 60%, vaccine at 70% and 85%, ITN 50% plus vaccine 70%, and ITN 50% plus IRS 40% plus vaccine 70%. All used the fitted vector and common 720-day counterfactual. Equation 16 defines expected reported episodes and ODE deaths averted. A combination interaction was calculated as the combined effect minus the sum of constituent effects; it is a model contrast rather than an empirical biological synergy estimate.

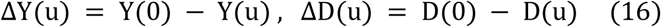

The economic perspective is an illustrative programme budget for additional prevention over one 720-day package. Background essential case management remains outside this additional budget. Household costs, productivity losses, avoided treatment expenditure and health-system capacity are excluded. The currency inputs are hypothetical US-dollar quantities, with no measured base price year or inflation conversion. Nominal prevention package costs are not discounted within the horizon.

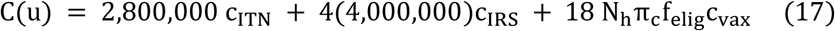

ITN cost is US$28,000 per percentage point for one campaign, assuming coverage is maintained. IRS costs US$40,000 per percentage point per round, with four rounds at days 0, 180, 360 and 540. Its 720-day input is therefore US$160,000 per percentage point. The vaccine target is 68,000 eligible children, matching the target stock in the protection calculation. An assumed US$18 completed four-dose course contains US$11.96 commodity and US$6.04 delivery inputs. One percentage point costs US$12,240; 70% coverage costs US$856,800. Wastage, dropout and later birth cohorts are absent.

Average ratios divide package cost by positive benefit against zero additional spending. They do not establish an incremental cost-effectiveness frontier or an adoption threshold. No flat illness cost, benefit–cost ratio, GDP-per-capita rule or purported WHO per-case threshold is applied [16,17]. Clinical cost-of-illness evidence is considered in the discussion using its original care definitions.

### 2.9 Secondary health loss proxy

A secondary discounted years-of-life-lost proxy assumes remaining lifetimes of 86.7 years for the child group and 61.0 years for the older group, with continuous annual discount rate r = 0.03. Equation 18 gives its weights and change in proxy health loss. The zero-discount limits are the remaining lifetimes. These are hypothetical losses per model disease death, without an observed age-at-death distribution.

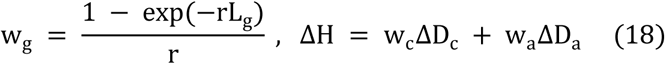

WHO Global Health Estimates use a reference loss function with no time discount or unequal age weighting [18]. The discounted proxy here has a different convention and omits disability, morbidity and sequelae. It is not a complete DALY or standard national YLL estimate. The undiscounted alternative is retained in the numerical package.

### 2.10 Constrained allocation and search checks

The primary budget is US$2,000,000. Equation 19 maximises reported episodes averted subject to intervention caps of 80% for ITN and IRS and 90% for vaccine. Additional recovery enhancement is fixed at zero. A stick-breaking expenditure parameterisation preserves budget feasibility, allocating remaining expenditure to the final intervention up to its cap. This searches budget-exhausting boundaries or cap-limited portfolios; it does not exhaustively prove dominance over all feasible interior allocations.

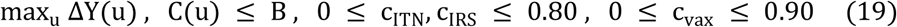

Vector-only and vaccine-inclusive searches each used five seeds, population size 12 per search dimension, at most 100 generations, tolerance 10⁻⁷ and polish. Every candidate and final result used the 0.5-day solver. An independent 101 × 101 expenditure-coordinate grid, including boundaries, supplied 10,201 checks. Explicit single-intervention boundaries and zero additional coverage were retained as benchmarks. Alternative true-onset, death and discounted-proxy objectives were evaluated under the same base assumptions.

### 2.11 Sensitivity and assumed parameter uncertainty

One-at-a-time analyses perturbed 12 non-redundant inputs by ±20%. Global sensitivity used 500 Latin-hypercube samples over independent ±20% ranges, seed 42, and partial rank correlation coefficients (PRCCs) for unforced R₀, reported episodes and ODE deaths [19]. K replaced separately sampled β₀ and M. Local derivative signs at 25 distributed points per input assessed whether the response was monotone over the screening ranges. Small partial correlations for inputs absent from an exact formula were not interpreted mechanistically.

The joint ensemble comprised 300 assumed draws, seed 20261011. K had a lognormal coefficient-scale of 0.15. Its log perturbation and the bounded reporting perturbation had an assumed correlation of −0.6. A and phase had bounded normal standard deviations of 0.08 and 8 days. Log-scale dispersions were 0.10 for mosquito mortality, 0.15 for recovery, 0.20 for immunity loss and 0.15 for *q*_c_. The common k input had a bounded 25% perturbation, aggregate efficacy a bounded 0.06 perturbation and the reporting ratio a bounded 0.10 perturbation. Reporting remained within 0.05–0.60. Age shares, eligible share and fatality probabilities were fixed.

Every draw was evaluated at every member of a 115-portfolio feasible pool assembled from an 11 × 11 expenditure grid and the main search solutions. Selection by expected reported benefit was compared with fitted-point selection and a candidate minimax-regret rule. Regret is the difference from the best member of that same pool for each draw. This is finite-pool optimisation under an assumed joint distribution; it is neither a continuous robust optimum nor a fitted posterior or confidence region.

### 2.12 Repeated noise and structural alternatives

At each monthly multiplicative Gaussian noise SD of 5%, 10%, 15% and 20%, ten independent two-age series were generated from the fitted deterministic expectation. Negative targets were floored at zero. Each target was recalibrated with seed 42 and up to 100 generations; allocation seeds 42, 1 and 7 were then applied to each fitted vector. This produced 40 targets and 120 allocation searches, separating between-target noise from within-target allocation-search randomness. Calibration-search randomness was not exhaustively repeated within each target.

Structural exercises changed the exponential saturation constants to 0.50 or 3.00 and replaced kill functions with linear *κ*_max_ c or Hill *κ*_max_ c/(0.5 + c) forms. Linear and Hill allocations each used three search seeds. Programme alternatives halved or doubled individual cost inputs and compared two or four IRS rounds while retaining the maintained-effect convention. Recovery diagnostics repeated the 70% enhancement after halving or doubling both immunity-loss rates. None of these alternative functions was fitted to an intervention trial.

### 2.13 Verification and external evidence

Seventeen executable assertions assessed matrix agreement, population conservation, positivity, zero coverage, parameter symmetries, reporting propagation, common initial conditions, exact aggregation, step refinement, caller-parameter preservation, vaccine–recovery routing, competing-risk fatality, matched cohort costing and invalid-input handling. An independently implemented Python/DOP853 system checked the RK4 output. Appendix B reports the executed checks. These tests concern mathematical and numerical implementation.

Public Uganda aggregate sources supplied epidemiological and measurement context [1,2,3,20]. They were not used as compatible independent validation observations. The historical compilation of 29 selected weekly records lacked the original bulletin PDFs in the accessible source package and was excluded from formal numerical validation. Routine confirmed cases, severe HMIS cases, estimated burden and survey positivity have different definitions. UMIS tests children aged 6–59 months; microscopy positivity is not the model’s clinical-infectious fraction *I*_c_/*N*_c_. No nominal correlation p values, four-layer empirical validation claim or held-out forecasting accuracy is attached to these comparisons.

## 3 Results

### 3.1 Calibration and synthetic fit

All five calibration runs terminated successfully, with loss values agreeing at the displayed precision (Table 1). The minimum-loss solution, seed 1, gave K = 1.169942, A = 0.459532, φ = 44.4351 days and ρ = 0.195138. These are fitted model inputs conditional on the fixed natural-history and observation assumptions. M was not independently fitted.

**Table 1.**
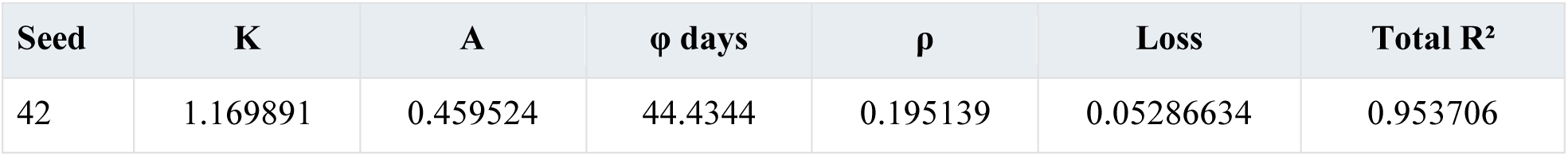
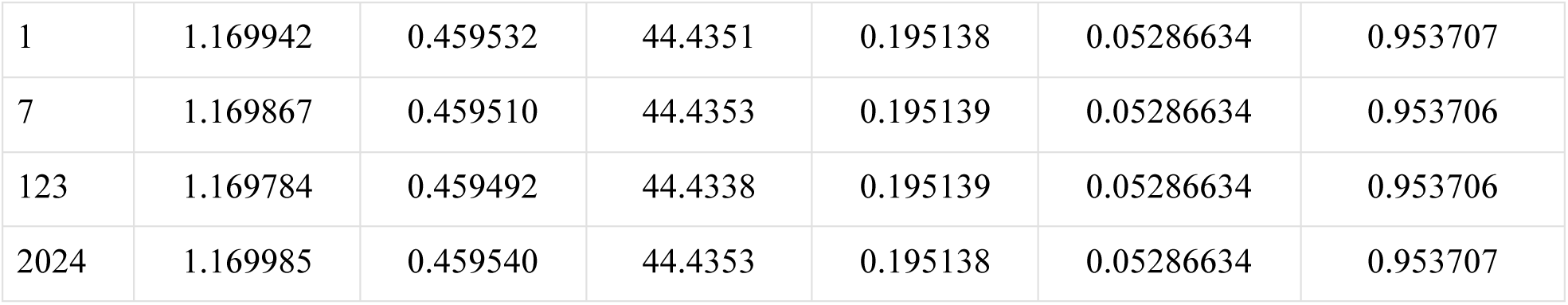
Calibration across five seeds using the historical synthetic target.

The total fit was stronger than either subgroup fit (Table 2 and Figure 3). Agreement in the total series therefore does not imply equivalent agreement in the age distribution. None of these diagnostics estimates out-of-sample accuracy, and none supplies a statistical confidence statement about Uganda. Because the target and current model use different episode and population conventions, the original generating inputs are not used to label this fit as parameter recovery.

**Table 2.**
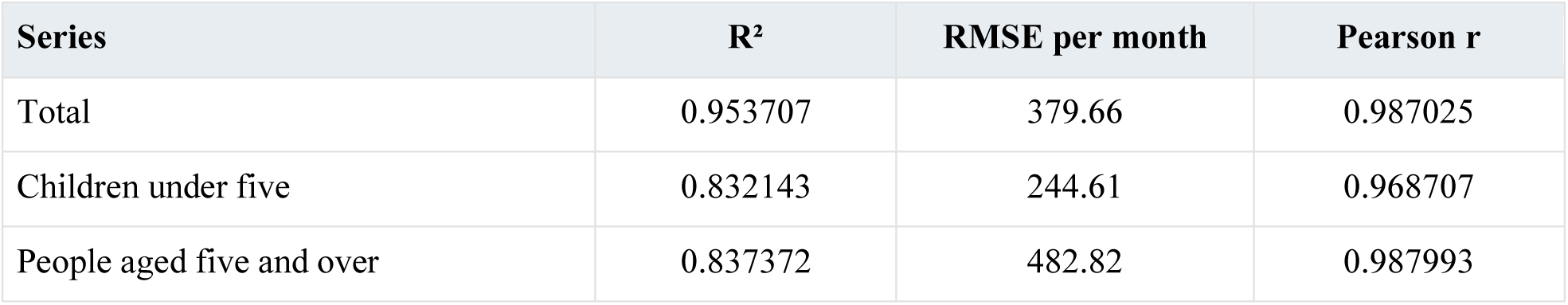
In-sample fit diagnostics on 36 calendar months.

**Figure 3.**
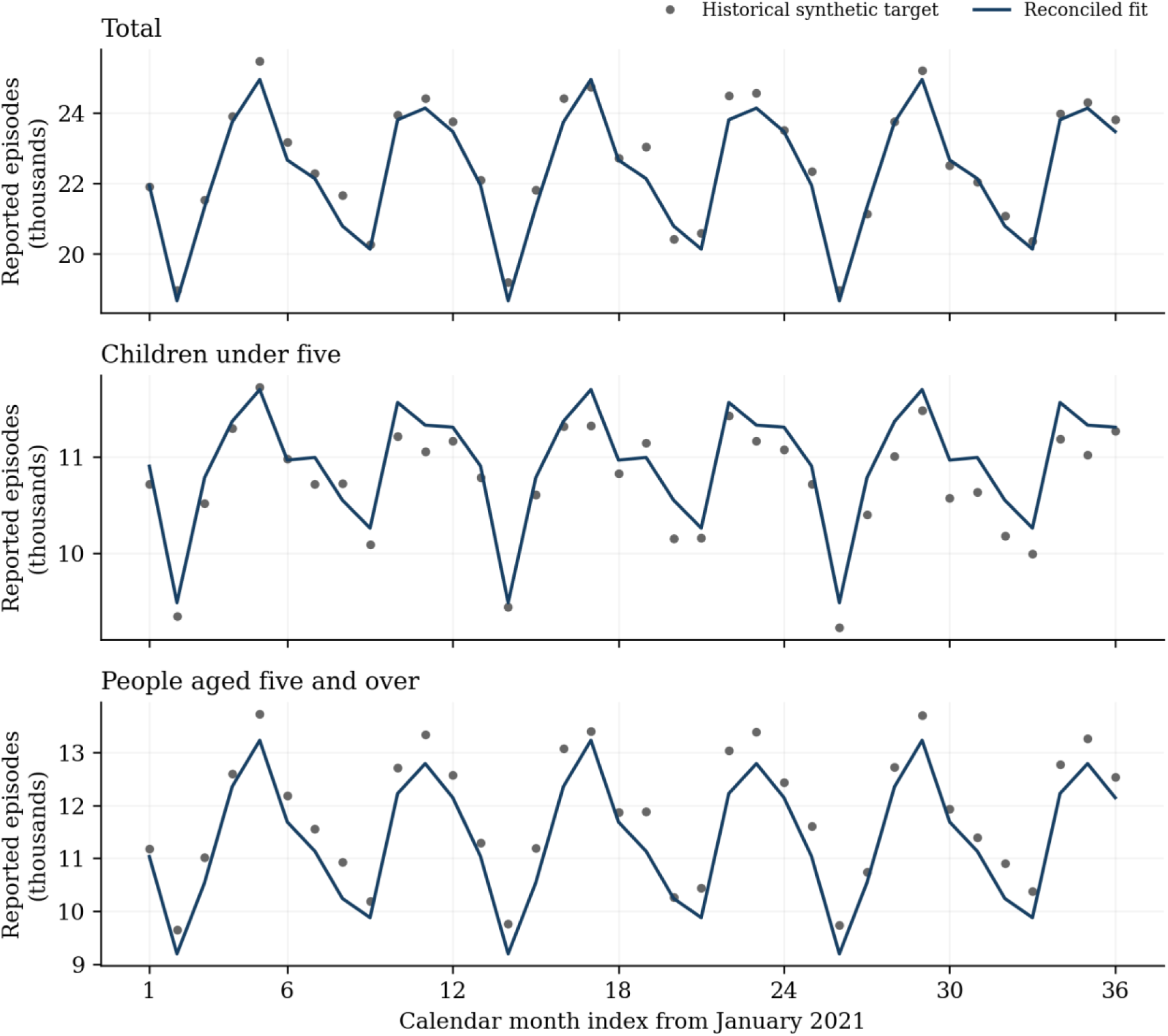
Historical synthetic monthly targets and reconciled model expectations. Total, child and older-group fits use the same parameter vector, reporting rule and calendar boundaries. Points are synthetic observations; lines are deterministic fitted expectations.

### 3.2 Structural confounding and recovery check

The vector-scale symmetry and child-product equivalence checks passed. Doubling M while holding K fixed changed reported human output by zero at displayed numerical precision. Rescaling the historical child factors without changing their product likewise preserved output. The equations therefore cannot distinguish β₀ from M or exposure from susceptibility using these human outputs alone. This is an algebraic result, not a judgement based solely on random-seed agreement.

The profiled composite criterion had conditional curvature over the examined intervals (Figure 4). Such curvature does not remove the separate structural symmetries or establish inferential confidence limits. In the noiseless same-model round trip, (1.33,0.45,55,0.20) was recovered as (1.330000013,0.450000005,55.000000170,0.19999999999), with total monthly RMSE below 0.00001. This establishes internal generation–fitting consistency for the stated fixture.

**Figure 4.**
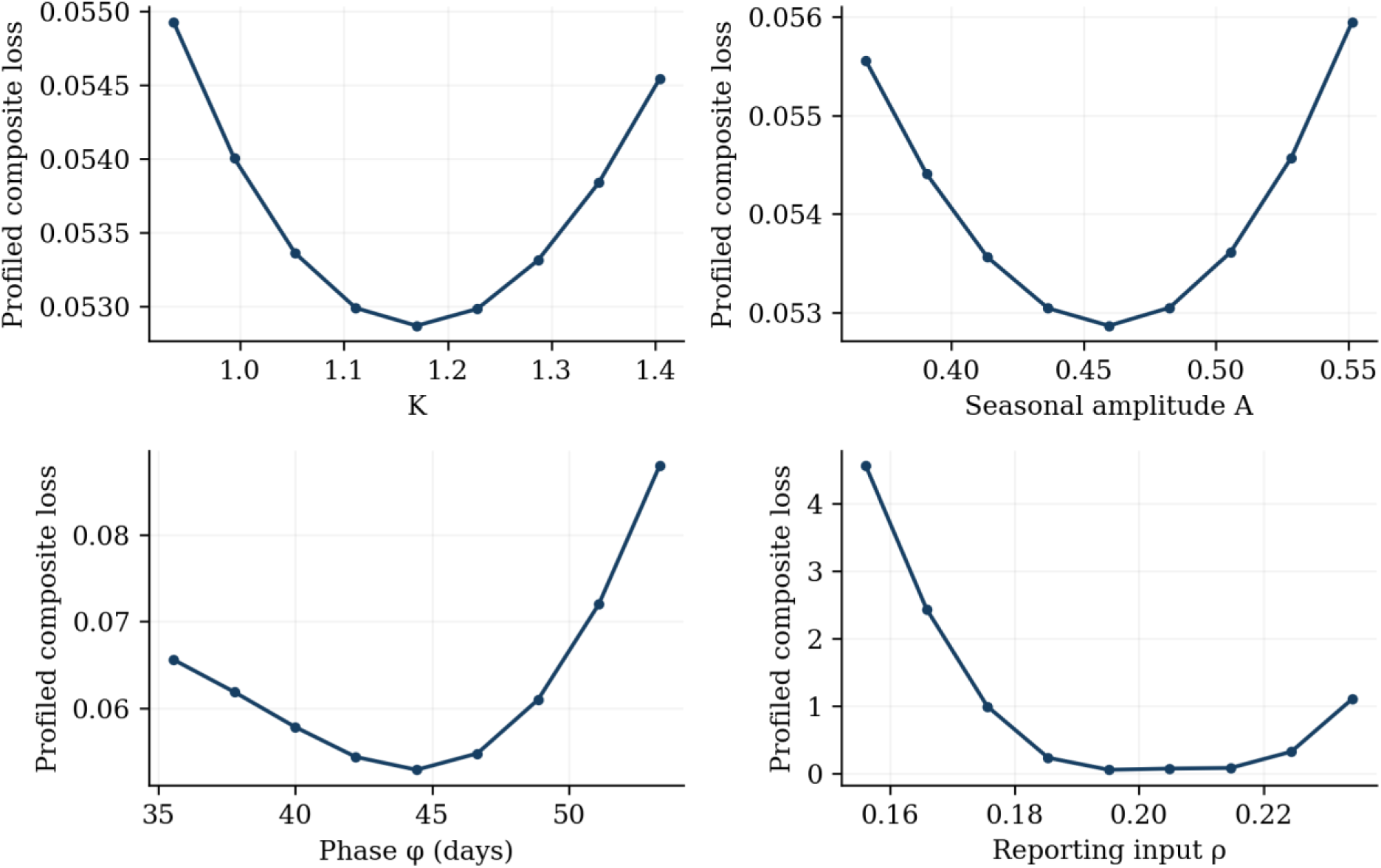
Nine-point profiles of the composite fitting loss. Each point fixes one input and reoptimises the remaining three. These curves are diagnostics under fixed assumptions and are not likelihood-ratio confidence intervals.

### 3.3 Baseline and reproduction diagnostics

The phase-aligned baseline converged in 4 annual cycles. Over 720 days it produced 2,453,677 model onsets before reporting and 528,041 expected reported episodes, comprising 258,877 child and 269,164 older-group episodes. Integrated ODE disease deaths were 3,612.40. These are model expectations rather than observed counts.

The derived reporting probabilities were *ρ*_c_ = 0.271150 and *ρ*_a_ = 0.179570. The episode-weighted baseline reported fraction was 0.215204, differing from the nominal population-share input because incidence rates differ between groups. Children accounted for 49.03% of expected reported episodes. No externally validated burden-share benchmark is inferred from these constructed outputs.

The unforced shared-vector closed form and numerical NGM both gave R₀ = 6.378679572, within the asserted tolerance of 10⁻¹⁰. The baseline annual Floquet multiplier was 1.303 × 10²², with log-growth 0.139511 per day. Under fixed 70% vaccine plateau protection, the multiplier was 3.592 × 10²¹ and growth 0.135981 per day. Both regimes permit invasion in this assumed disease-free seasonal system. No endemicity category or empirically supported elimination threshold follows from these diagnostics.

### 3.4 Prevention and vaccine scenarios

Every reduction in Table 3 uses the same baseline of 528,041 expected reported episodes. ITN 50% averted 36,692 episodes and 262.14 model deaths; ITN 80% averted 58,422 episodes and 413.92 model deaths. IRS 60% averted 59,133 episodes and 418.81 model deaths. The similarity of the last two effects is conditional on the assumed kill probabilities and maintained coverages, not an estimated field equivalence between ITN and IRS.

**Table 3.**
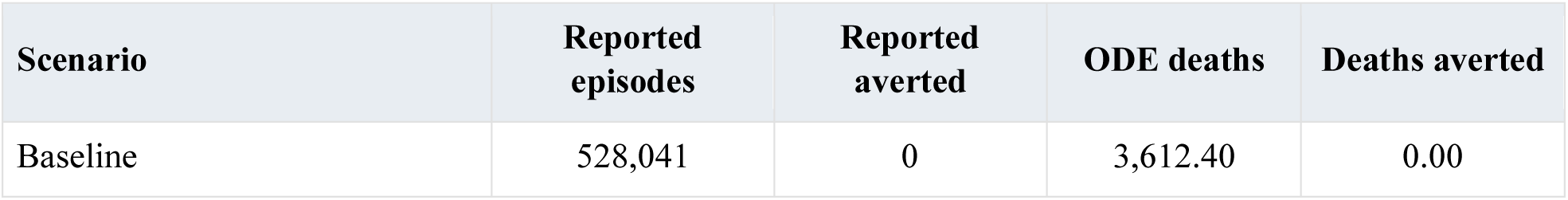
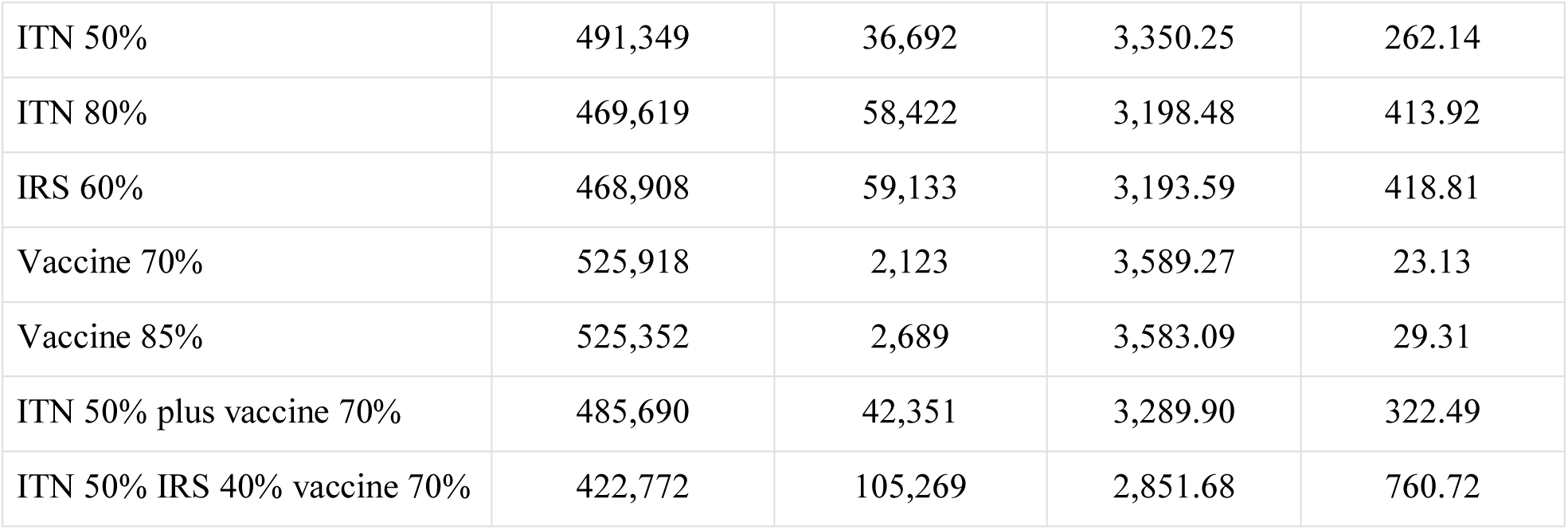
Common-state scenario outcomes over 720 days.

At vaccine coverage 70%, 2,123 reported episodes and 23.13 model deaths were averted. At 85%, 2,689 episodes and 29.31 model deaths were averted. The 70% result includes 2,092.55 child and 30.35 older-group reported episodes averted. The latter reflects indirect feedback through mosquitoes. The aggregate 17.08% plateau force-of-infection modification translated into approximately 0.81% fewer reported child episodes over 720 days, showing that a nominal hazard reduction and a realised episode reduction are different quantities in this immunity-cycling model.

ITN 50% plus vaccine 70% averted 42,351 reported episodes. Subtracting the separate ITN and vaccine benefits left an interaction of +3,536.06 episodes. ITN 50% plus IRS 40% plus vaccine 70% averted 105,269 episodes and 760.72 model deaths. The interaction sign arises from the nonlinear equations, initial conditions and finite horizon. It does not establish biological synergy or the expected effect of Uganda’s actual programme.

**Figure 5.**
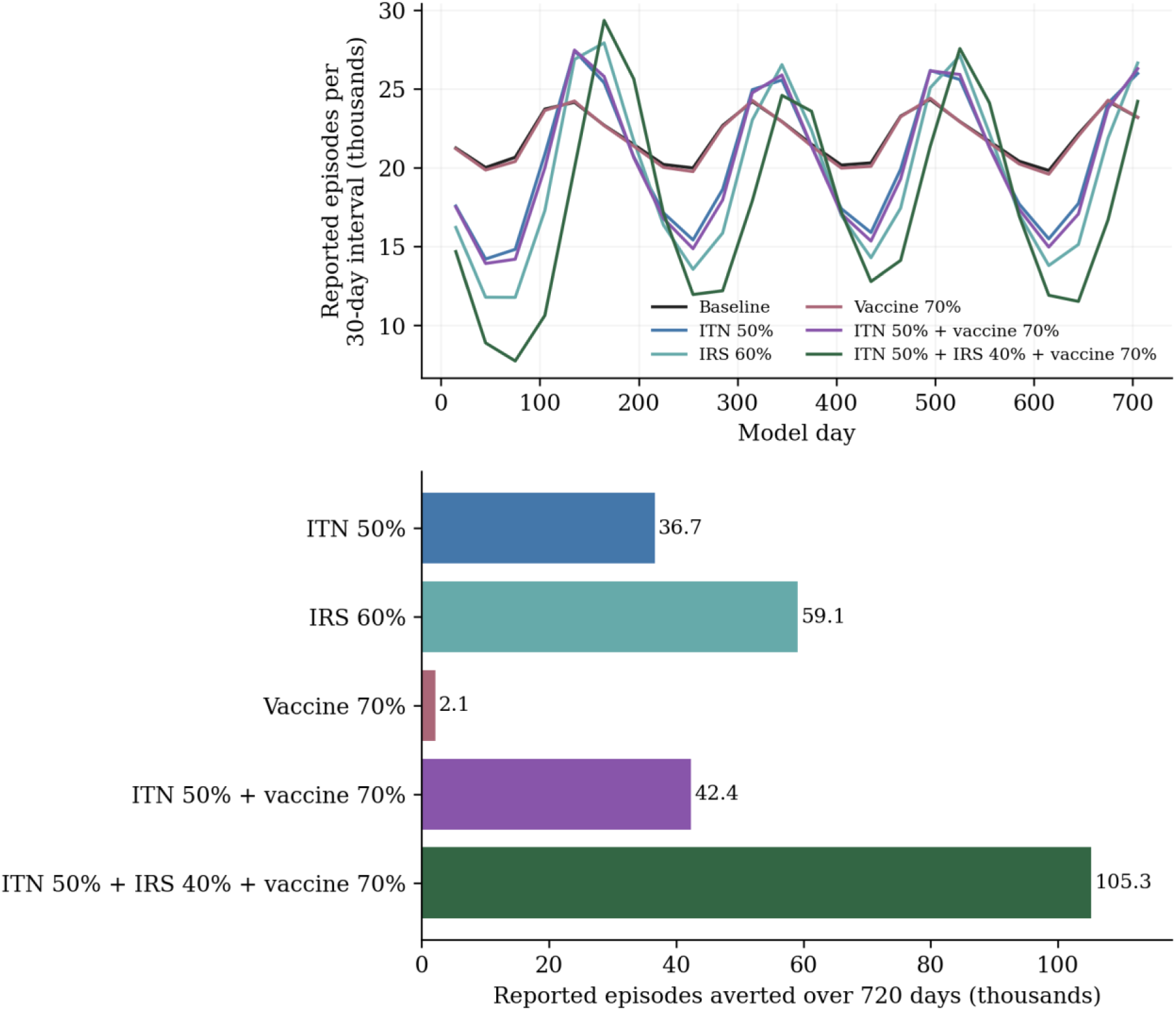
Common-state prevention simulations. Upper panel shows deterministic expected reported episodes in consecutive 30-day intervals; lower panel shows cumulative reductions against the same baseline over 720 days. Vaccine coverage is completed-course coverage of the fixed eligible target.

### 3.5 Recovery and temporary immunity

The 70% recovery-only diagnostic produced 540,820 reported episodes, 12,779 more than baseline, and 3,845.64 ODE disease deaths, 233.24 more than baseline. Its unforced reproduction index fell to 4.4045. Lower infectious duration and a smaller invasion index therefore did not imply fewer finite-horizon episode onsets in these equations. Earlier recovery and subsequent loss of temporary immunity altered the time available for repeated episodes.

The episode contrast changed sign after halving both immunity-loss rates: 4,237 episodes were averted. At the reference rates the contrast was −12,779, and doubling the rates gave −61,420. These calculations used a separate common baseline for each natural-history setting. The mortality response remains affected by holding infectious-exit fatality fixed. The experiment does not represent the clinical benefit or harm of effective antimalarial treatment, and no recovery-diagnostic economic ratio or treatment budget recommendation is reported.

### 3.6 Conditional prevention costs

**Table 4.**
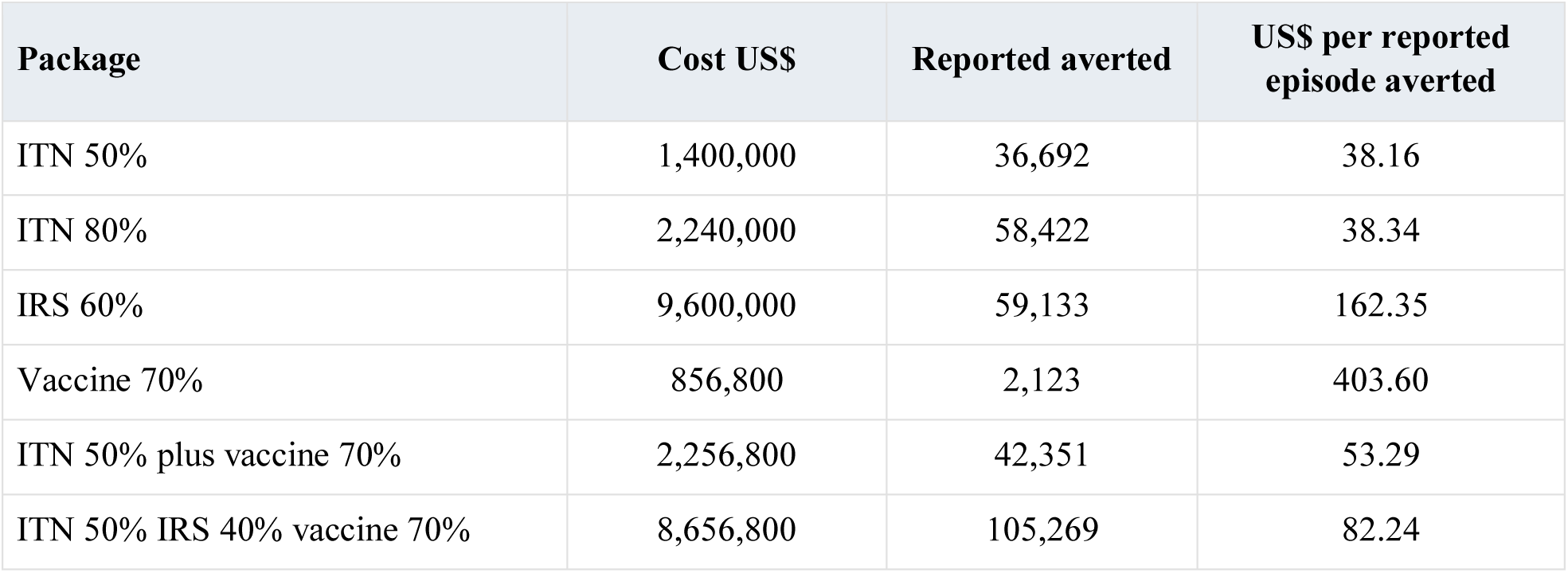
Additional prevention package costs and average episode ratios.

The ITN 50% package cost US$1.4 million and had an average ratio of US$38.16 per reported episode averted. IRS 60% cost US$9.6 million because four rounds were counted, yielding US$162.35 per reported episode averted. Vaccine 70% cost US$856,800, yielding US$403.60 per reported episode averted. The three-intervention combination cost US$8,656,800. Differences in these ratios reflect both the assumed effects and the stated package resources; they do not compare equal-cost programmes or establish an incremental economic frontier.

### 3.7 Budget search and benchmark

**Table 5.**
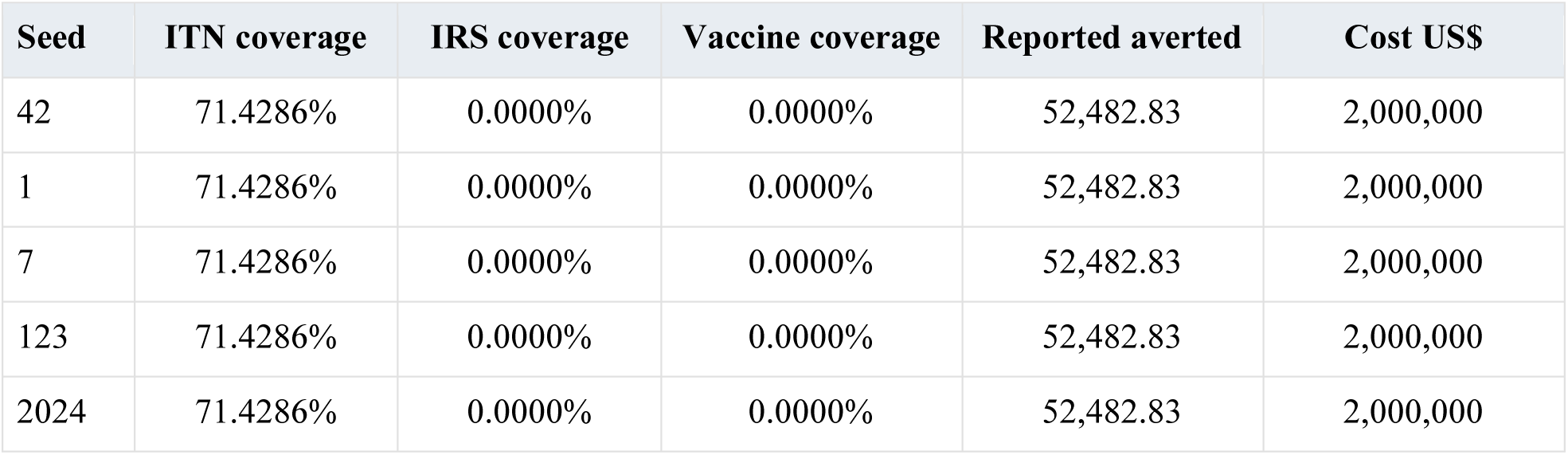
Vaccine-inclusive US$2 million search across five seeds.

All five searches selected the ITN corner to displayed precision. The independent 10,201-point grid selected exactly 71.428571% ITN coverage, with zero IRS and vaccine additions, costing US$2 million and averting 52,482.83 reported episodes. This is a coverage fraction; the corresponding additional spending share is 100% ITN. Vector-only search and the alternative true-onset, death and discounted-proxy objectives also selected this corner under the base assumptions.

The selected boundary averted 242,309.42 model onsets before reporting and 372.85 ODE disease deaths. The discounted proxy weights were 30.860022 and 27.986214 years per model death, giving 11,289.46 proxy years averted. The episode ratio was approximately US$38.11 per reported episode averted. No claim of standard DALYs, marginal cost-effectiveness, a global continuous optimum or a current Uganda allocation is made from these quantities.

### 3.8 Sensitivity of outputs

**Table 6.**
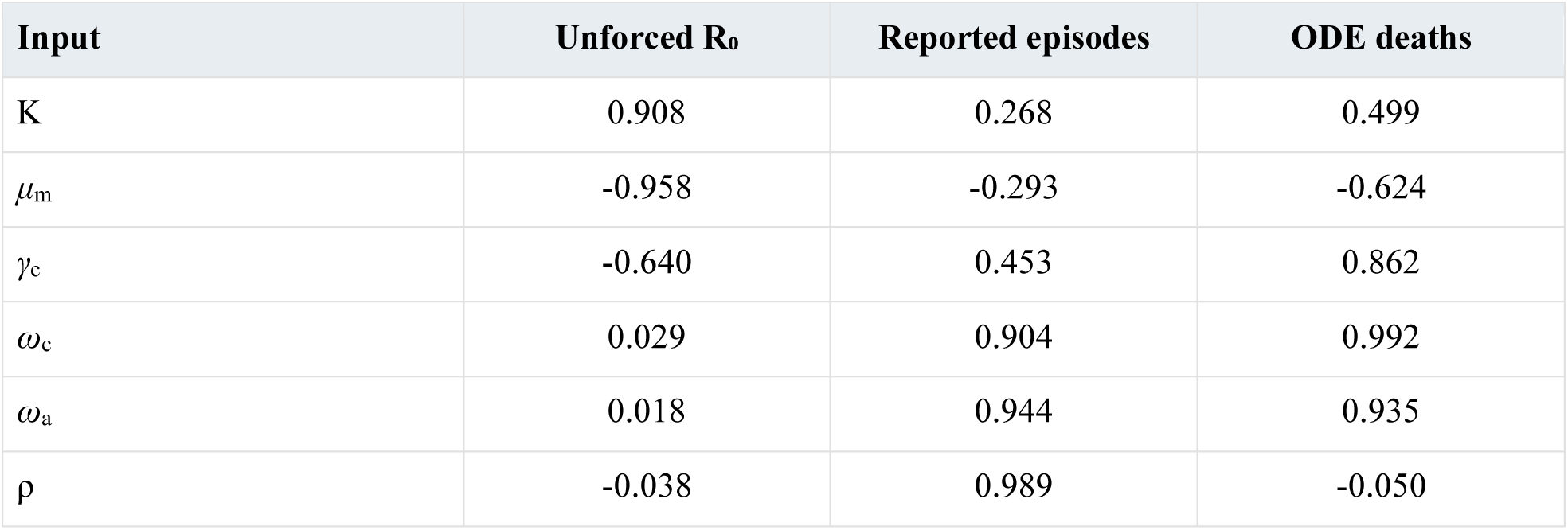
Selected PRCCs from the 500-sample sensitivity screen.

The unforced reproduction index was most strongly associated with mosquito mortality (PRCC−0.958), K (+0.908) and *β*_m_ (+0.906) within the screened ranges. Reported episodes were strongly associated with reporting (+0.989) and immunity loss in the older (+0.944) and child (+0.904) groups. ODE deaths were strongly associated with child immunity loss (+0.992), older immunity loss (+0.935) and child recovery (+0.862). These different rankings show that invasion intensity, observed episodes and model mortality are distinct outputs.

Figure 6 displays all 12 screened inputs. Exact unforced R₀ is independent of reporting, immunity loss and seasonal amplitude, so their small sampled PRCCs in that panel carry no mechanistic meaning. Recovery responses require additional caution where local derivative signs are non-monotone. PRCCs are conditional influence summaries over chosen ranges, not causal effects or measures of empirical parameter importance.

**Figure 6.**
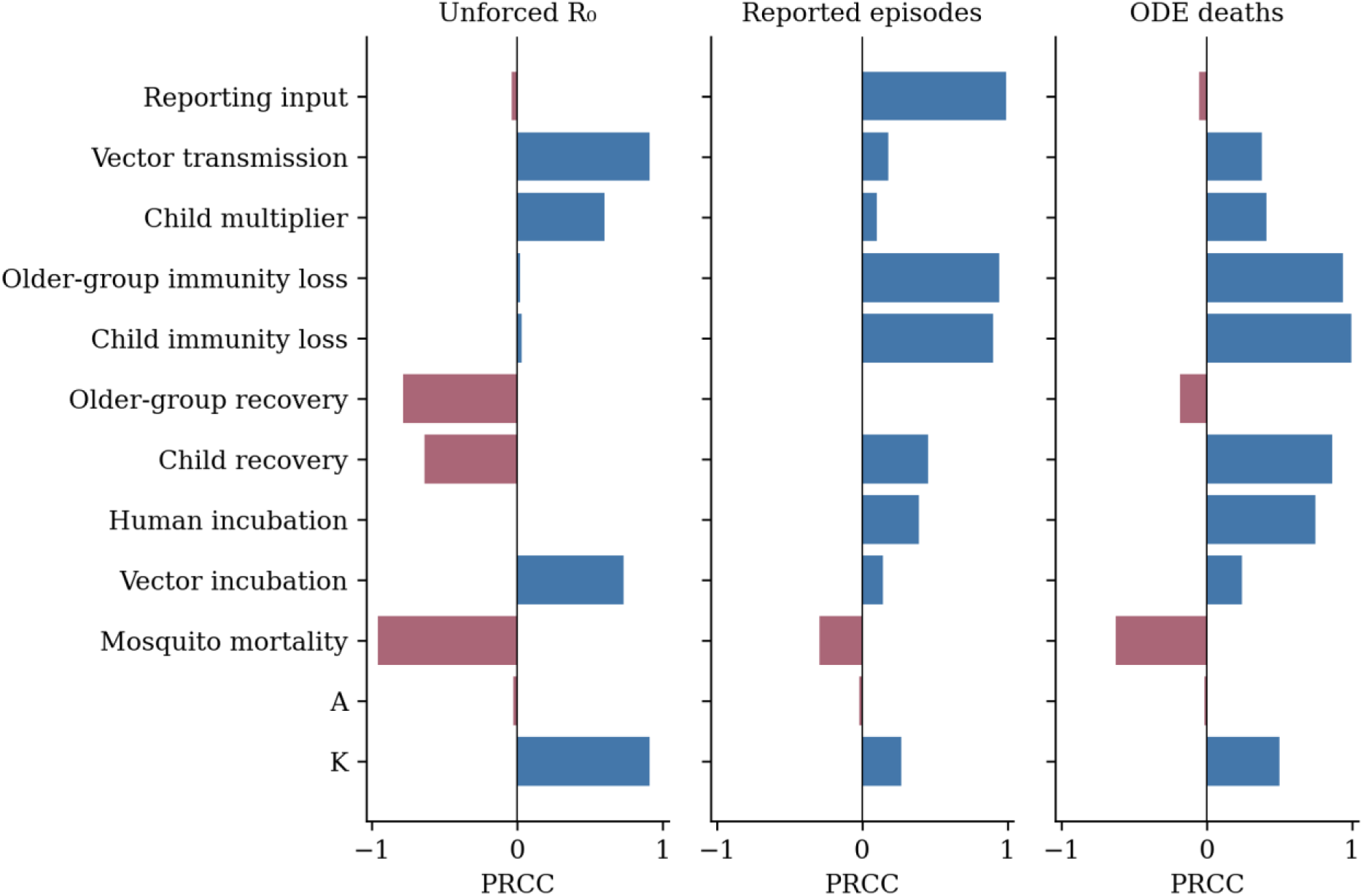
Partial rank correlations for 12 non-redundant inputs varied independently by ±20%, using 500 Latin-hypercube samples. Invasion, reported episodes and ODE deaths have different sensitivities. Small correlations for inputs absent from the exact R₀ formula are sampling effects.

### 3.9 Full finite-pool ensemble selection

**Table 7.**
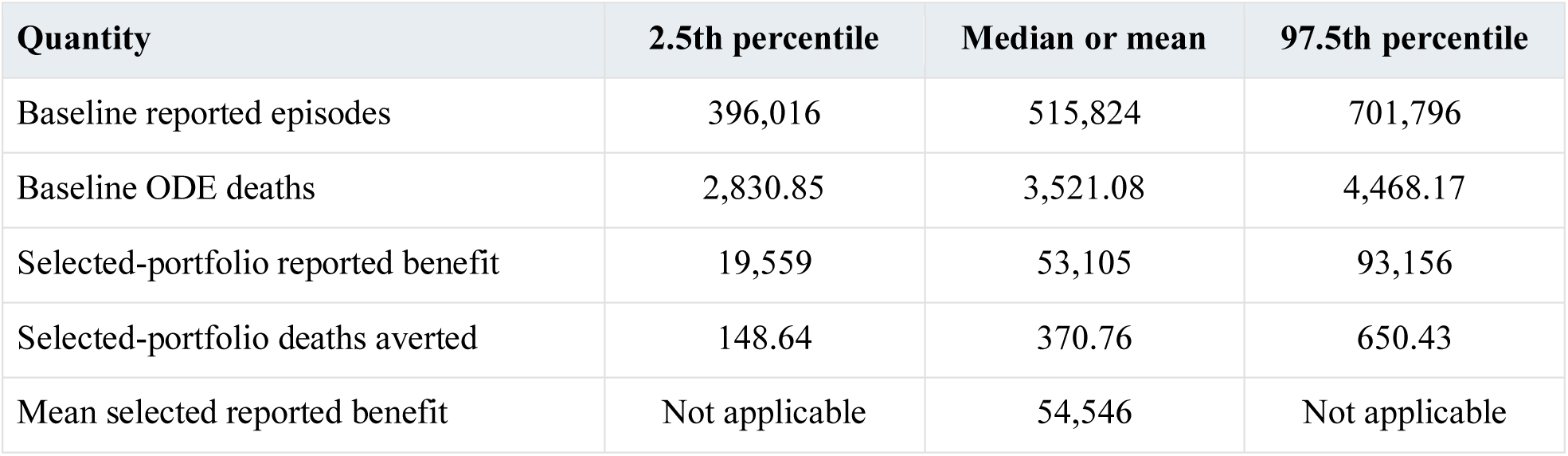
Results across 300 assumed parameter draws.

The fitted-point, expected-benefit and candidate minimax-regret rules all selected the same ITN corner. It was best within the 115-portfolio pool for each of the 300 draws, with zero maximum candidate-relative regret to numerical precision. Mean reported benefit was 54,546.49 episodes. The 2.5th, median and 97.5th percentiles were 19,558.96, 53,105.30 and 93,156.14; the corresponding death benefits were 148.64, 370.76 and 650.43. Baseline reported episodes ranged from 396,016 to 701,796 between the 2.5th and 97.5th percentiles.

Portfolio stability coexisted with large variation in effect size. The distribution held programme prices, eligibility and several clinical assumptions fixed. The percentiles therefore describe this specified experiment and are not 95% confidence limits for Uganda. Candidate-relative zero regret also excludes unexamined portfolios and structural models from its comparison.

### 3.10 Repeated observation noise

**Table 8.**
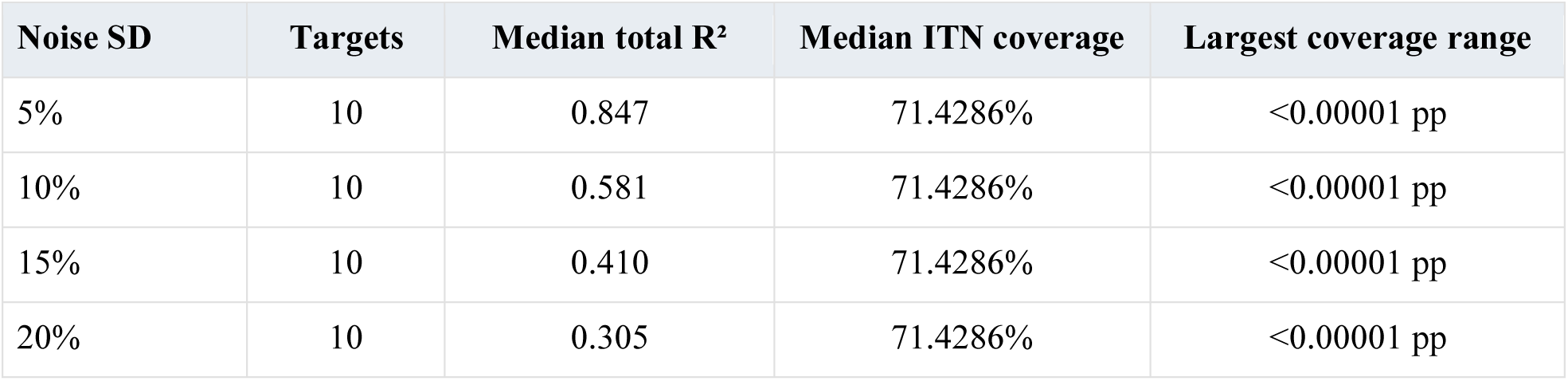
Recalibration and allocation after repeated noisy monthly targets.

All 40 target recalibrations and 120 allocation searches completed successfully. Median total R² fell from 0.847 at 5% noise to 0.581, 0.410 and 0.305 at 10%, 15% and 20%. The ITN corner remained selected to displayed precision, with allocation coverage differences below 0.00001 percentage points both between targets and within-target search seeds. This stability reflects the same strong base-cost boundary despite degraded fit. It does not establish resistance to real reporting-system changes or clinical outcome misspecification.

**Figure 7.**
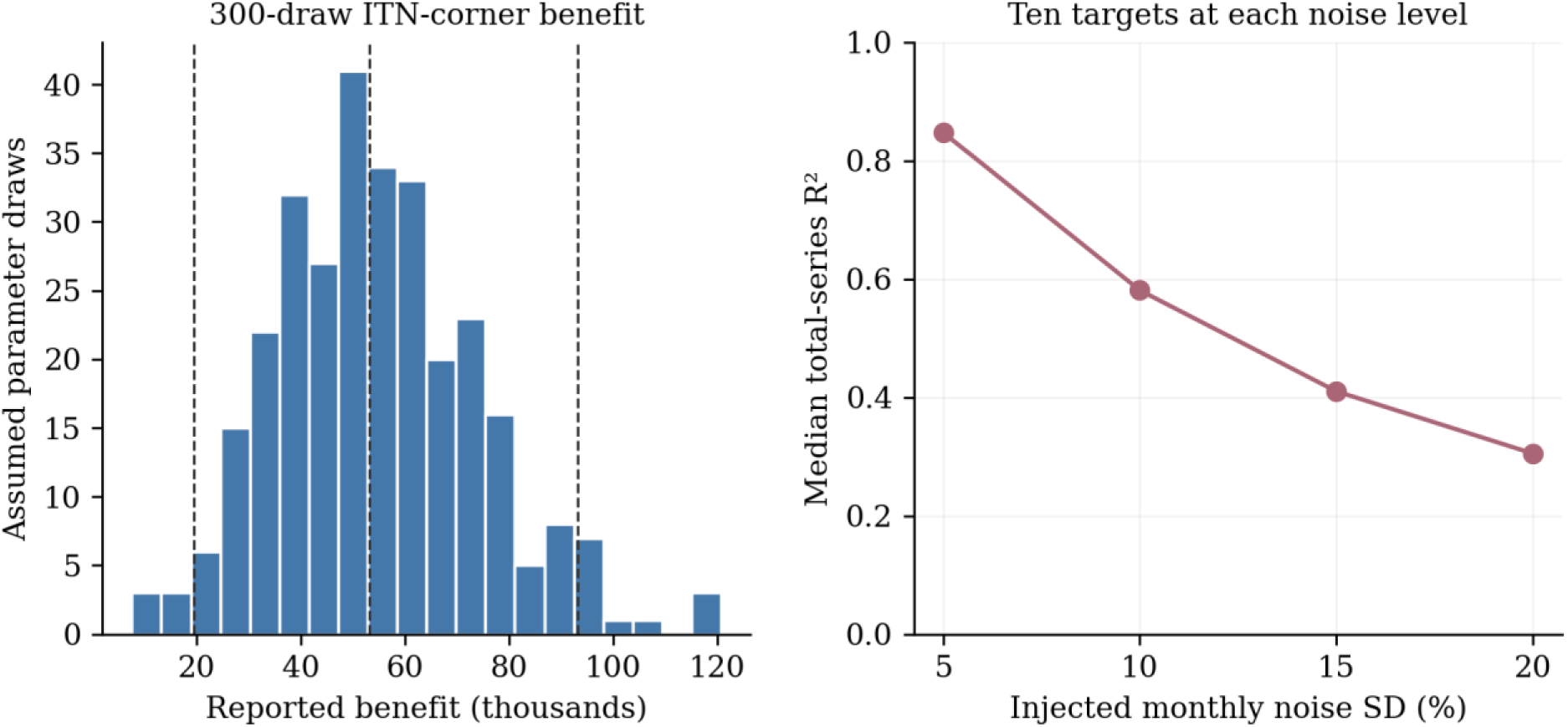
Assumed-parameter and synthetic-noise diagnostics. Left panel shows reported benefit of the selected ITN corner across all 300 parameter draws; dashed lines mark the 2.5th, median and 97.5th percentiles. Right panel shows median fit across ten targets per noise level. Neither panel is an empirical confidence assessment.

### 3.11 Structural and programme alternatives

**Table 9.**
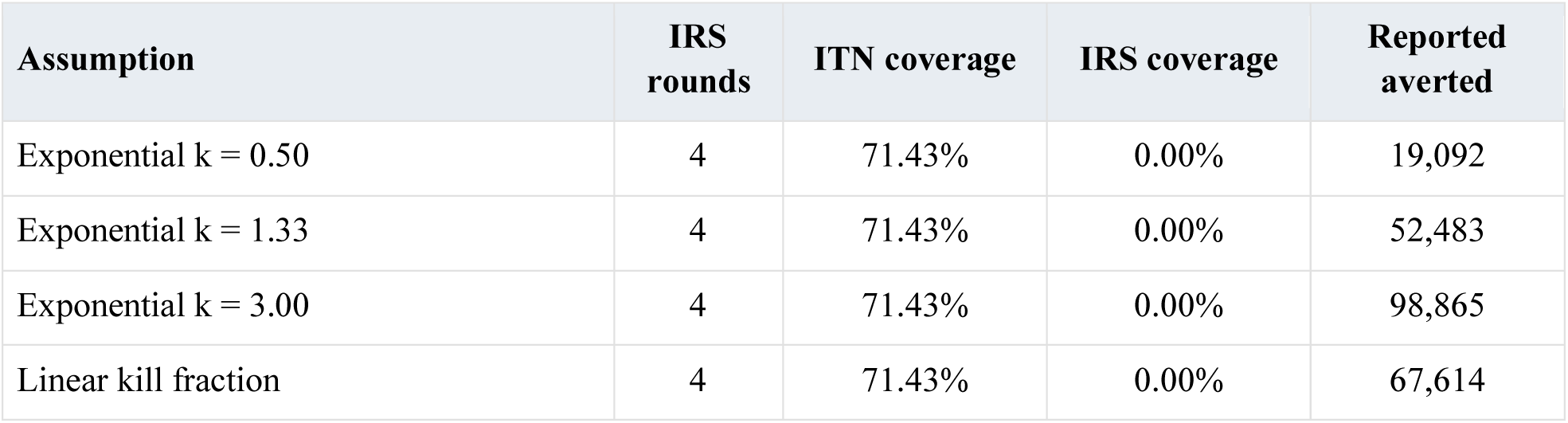
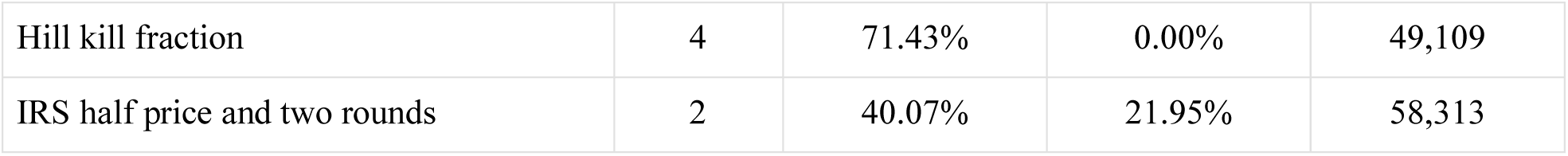
Effects of response-function and IRS package assumptions.

The selected base-cost ITN corner averted 19,092 episodes with exponential k = 0.50, 52,483 with k = 1.33 and 98,865 with k = 3.00. Linear and Hill curves gave approximately 67,614 and 49,109 episodes averted while selecting the same corner. Thus the apparent allocation stability did not imply a stable quantitative benefit.

Halving the IRS round price and counting two rounds changed the searched allocation to approximately 40.07% ITN and 21.95% IRS, with 58,313 episodes averted. The alternative retained maintained IRS efficacy, so it isolates an assumed cost-frequency change rather than estimating a residual-effect programme. The full half-price, double-price and two-/four-round results are retained in the reference package. These shifts rule out a general national claim that the base-cost corner is invariably optimal.

### 3.12 Mathematical and numerical checks

All 17 assertions passed (Appendix B). The closed form and NGM agreed at baseline and under a combined intervention. Maximum population-total error was below 1.4 × 10⁻⁹ persons or mosquitoes. Zero additional coverage matched baseline exactly, and all tested extreme trajectories remained non-negative. Reporting ratios, shared starting states, interval aggregation, caller-input preservation and vaccine–recovery propagation were consistent.

Reducing the step from 0.5 to 0.25 days changed the reported total by a relative 1.87 × 10⁻⁹. The independent DOP853 implementation differed by a relative 2.08 × 10⁻⁹, with a maximum interval discrepancy of 0.000645 episode. The matched-cohort cost test reproduced US$8,656,800 for ITN 50% plus IRS 40% plus vaccine 70%, using 68,000 eligible children. Invalid inputs were rejected, and the main search costs respected the US$2 million budget within floating-point tolerance. These checks verify the stated implementation and do not assess clinical validity. Appendix D provides the additional model and trajectory plots.

## 4 Discussion

### 4.1 What the study establishes

The calculations answer the four research questions within a small assumed system. Human case outputs determine the composite transmission scale K under the present density convention, while β₀ and M remain structurally confounded. The historical child labels similarly reduce to a single multiplier. Recovery can change repeated episode cycling in either direction when temporary immunity changes. The prevention portfolio is stable under the examined parameter and observation perturbations but changes under a declared IRS cost-frequency alternative.

These findings distinguish model identification, implementation checks and conditional intervention comparisons. Repeated searches and agreement with an independent solver strengthen confidence that the reported tables follow the equations. They cannot strengthen an unsupported clinical or resource assumption into a measured quantity. A reproducible answer may still depend on a mechanism that is unsuitable for the decision being considered.

### 4.2 Identifiability and observation compatibility

The vector-scale symmetry explains why collecting more monthly human counts under the same observation rule cannot separately recover β₀ and M. Independent entomology could constrain density, but converting that evidence into a model input would require a matching definition of vector abundance, sampling and contact. The same caution applies to splitting child risk into exposure and immunological susceptibility. Distinct names in a parameter table do not identify distinct effects when the equations use only their product.

The historical target’s different acquisition definition also explains why the current refit and a same-model recovery test must be reported separately. A model can recover its own generating inputs almost exactly and still provide imperfect subgroup fit to another synthetic implementation. An empirical study would need to define how infection, symptom onset, testing, consultation and reporting lead to each recorded count. Reporting completeness is not interchangeable with the probability of an episode appearing in surveillance.

### 4.3 Vaccine and recovery interpretation

Putting vaccine protection inside the infection process preserves feedback that is lost when simulated child cases are reduced only after the transmission equations have been solved. The resulting small vaccine-only episode effect is a property of this aggregate structure: susceptible-pool expansion, temporary immunity and the chosen horizon partly offset the nominal hazard reduction. It is not a contradiction of clinical R21 efficacy [5] or a prediction of programme failure. The positive combination interaction is equally conditional.

Programmatic vaccine evaluation requires eligibility entry, age and dose cohorts, completion, waning, boosters and compatible clinical outcomes [11]. Spreading a fixed eligible stock’s protection across all child states cannot reproduce those processes. The same stock was used for current protection and costs to avoid internal denominator disagreement, but matching internal denominators does not establish that the stock approximation is empirically appropriate.

The recovery-only experiment cannot rank clinical treatment investments. It fixes the disease-fatality probability at infectious exit and omits treatment-specific protection from severe disease and death. A rise in its model mortality therefore reflects repeated onsets and the imposed fatality convention. An intervention model intended to guide case management would have to represent clinical access, severity, treatment eligibility, treatment-specific outcomes and resources. The present prevention search maintains background recovery and gives no basis for reducing essential diagnosis or treatment.

### 4.4 Budget assumptions and health outcomes

The base selection follows a hypothetical 720-day package in which one maintained net campaign is compared with four IRS rounds. Changing an IRS price and frequency assumption changes the selected mix without changing the fitting data. This is evidence that resource definitions belong inside sensitivity analysis. A coverage fraction, a spending share and a realised health effect are also different quantities: 71.43% ITN coverage consumes the entire additional US$2 million budget in the base corner.

The study does not establish a societal benefit–cost ratio. Orem and colleagues report historical Uganda morbidity costs [21], while Snyman and colleagues estimate care-specific societal costs in constant 2022 US dollars, including US$19.02 per confirmed outpatient episode and US$29.29 per confirmed inpatient episode [22]. Applying either amount to every model onset would require an explicit confirmation and care pathway. Inflating a historical national illness estimate with an unverified index would not supply current Ugandan programme prices.

The discounted YLL proxy is a secondary assumption-based output. Its correctly computed weights still lack an age-at-death distribution, and two broad age groups cannot support a standard age-specific burden calculation. WHO’s undiscounted health-loss convention differs from this illustration [18]. A full economic evaluation would also need morbidity and sequela states, dated unit prices, recurrence and implementation constraints, a clear perspective, incremental dominance analysis and a locally relevant decision threshold [16].

### 4.5 Robustness and external evaluation

Using all 300 parameter draws at every candidate is more informative than selecting three draws by reproduction-index percentile: portfolios can behave differently even when a single scalar index is similar. The finite-pool result nonetheless remains conditional on the assumed joint distribution and pool. The broad benefit interval and unchanged corner show that stable allocation does not imply precise benefit. Repeated noise targets similarly identify an allocation boundary that persists as synthetic fit deteriorates.

No compatible independent clinical observation set was available to validate the current calculations. WHO burden estimates, routine confirmed and severe HMIS cases, and UMIS infection testing each require their own observation mapping [1,2,3,20]. Selecting a different seasonal comparator after inspecting a failed comparison cannot establish prespecified predictive performance. Autocorrelated monthly values and sparse pooled weeks also require more than nominal correlation significance. Future evaluation should specify outcomes and assessment criteria before examining an independent period or location, and compare performance with simple baseline predictions [8,9,23].

### 4.6 Software and remaining limits

The numerical reference uses one C++17 RK4 core and Python interface for fitting, scenarios, reporting and programme arithmetic. It rejects invalid inputs and preserves caller parameters. The ordered reproduction runner, source hashes, complete output arrays and executable assertions permit inspection of the reported calculations [24,25,26]. This contribution concerns the scientific reference implementation. The historical interactive dashboard, conversational assistant, browser runtime and multi-year planner were not reconciled or performance-tested as part of this release.

Other limitations include homogeneous mixing, maintained virtual demographics, fixed vector density, assumed kill functions, exponential return to full susceptibility, and omission of asymptomatic infection, severe disease, pregnancy, resistance, spatial movement and distributional objectives. The budget search emphasises expenditure boundaries and does not prove a global continuous optimum. The study reports a single fixed portfolio over 720 days; it makes no five-year optimal-control claim, district resource recommendation or comparison with the current national strategy.

### 4.7 Next empirical study

The next step is to obtain governed observations with compatible definitions: age-specific confirmed case series, population denominators, test volume, reporting completeness, care pathways, intervention access and use, dose records and programme resource quantities. Entomology and measured rainfall would then constrain the vector and seasonal mechanisms. A new observation likelihood and independently defined evaluation period should precede parameter estimation, rather than replacing the synthetic target without changing the measurement model. Appendix C identifies the corresponding data requirements.

A clinical and vaccine extension should be developed before expanding the prevention result into an operational recommendation. Cohorts, severity and treatment outcomes should be linked to the costed population. Spatial and equity objectives, campaign delivery, decay and actual programme recurrence can then be evaluated with appropriate local data. This sequence follows the limitations demonstrated here; no Ministry partnership, funded timetable or future validation result is assumed.

## 5 Conclusions

The reconciled SEIR–SEI framework produces traceable, numerically verified synthetic calculations. It demonstrates structural confounding of transmission and vector density, equivalence of the historical child-risk product, and the dependence of recovery cycling on temporary immunity. At the stated US$2 million additional budget, the base-cost prevention searches select an ITN corner that remains best within the examined ensemble pool and repeated-noise experiments. A different IRS cost-frequency assumption changes that selection, and assumed intervention curves materially change the estimated benefit.

These are conditional results from an explicit model and programme package. They establish an executable starting point for empirical work, while leaving clinical effectiveness, current national costs, forecast performance and operational allocation unvalidated. Compatible observations and evidence-supported intervention and resource mechanisms are required before the framework can support Uganda programme decisions.

## Supporting information

Protocol

## Data Availability

The study uses a 36-month historical synthetic calibration dataset and model-generated numerical results. No individual-level patient data were analysed. The accompanying reproducibility archive, UMAPS_Research_Paper_Analysis_v05.zip, contains the unchanged synthetic target, reference source code, numerical outputs, dependencies, validation assertions and input/output hashes. The supporting materials are available from the corresponding author upon reasonable request, subject to applicable third-party source rights. Publicly published Ugandan epidemiological information was used for context and is cited in the manuscript; it was not used as an independent empirical validation dataset.

## Declarations

### Ethics and data

The activity analysed synthetic observations and publicly published aggregate information. It recruited no participants and used no identifiable individual-level health records. No ethics approval or exemption determination is documented for this version. Future restricted-data access requires appropriate review, custodian permission and an authorised governance arrangement; no institutional affiliation or clearance is implied.

### Author responsibility and assistance

Jay Hiteshkumar Patel is the named investigator and is responsible for interpretation, source permissions and submission. AI assistance was used to reconcile equations, implement and execute the reference analysis, and edit and format this manuscript. This assistance does not constitute independent clinical expert review or institutional endorsement. Disclosure must follow the receiving journal’s requirements.

### Data and code availability

The accompanying UMAPS_Research_Paper_Analysis_v05.zip contains the matching reference source, unchanged synthetic target, complete numerical outputs, dependencies, assertions, input and result hashes, and manuscript authoring code. The newly written numerical routines carry an MIT licence; historical data and source documents are not relicensed. No public repository, OSF deposit or persistent DOI is claimed. Appendix C describes reproduction and release scope.

## Appendix A Reference parameters and symbols

Table A1 distinguishes fitted quantities, fixed scales and illustrative assumptions. A source may support a mechanism or provide relevant context without measuring the exact value used here. The executed numerical vector is retained in results/parameters.json.

**Table A1.**
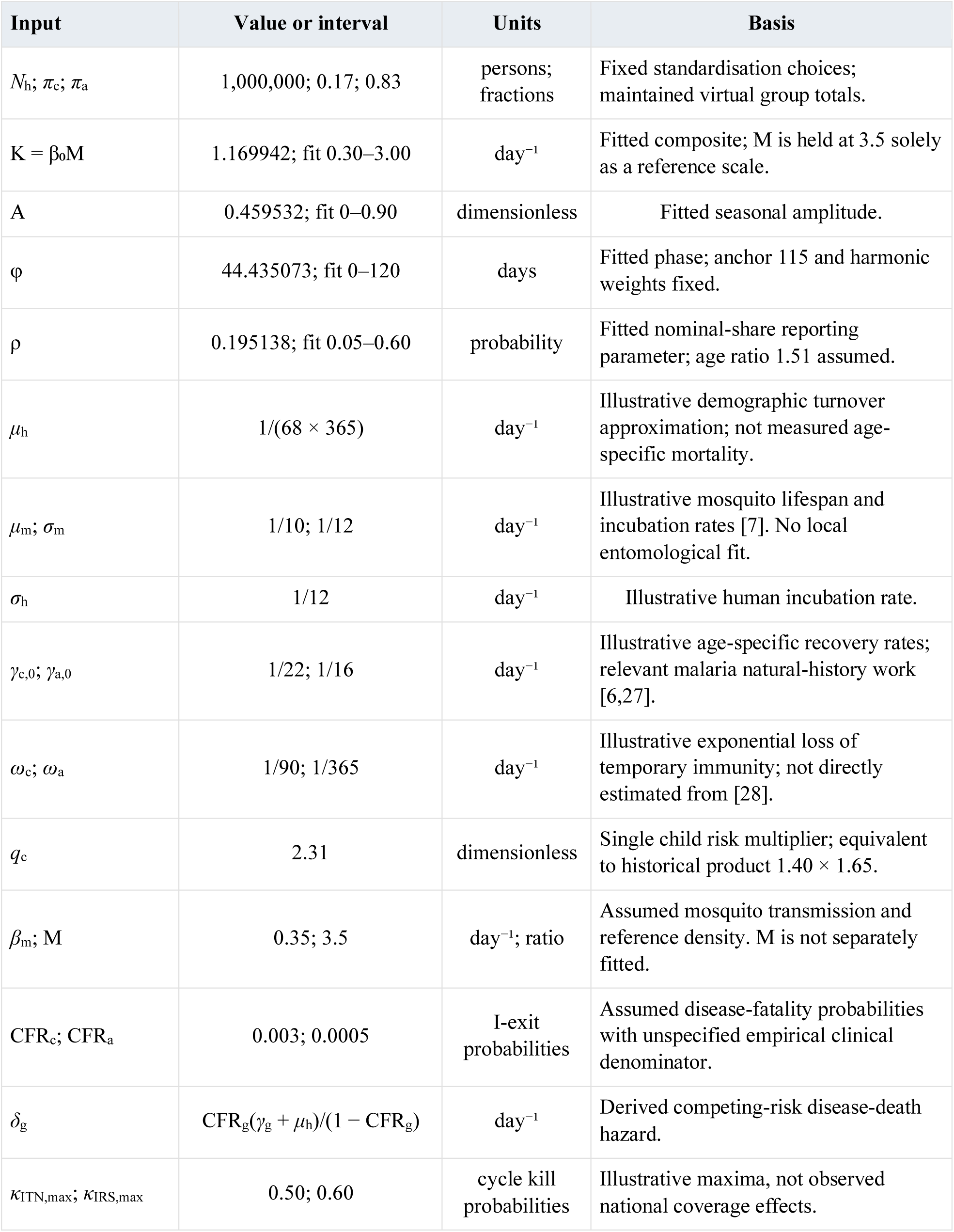
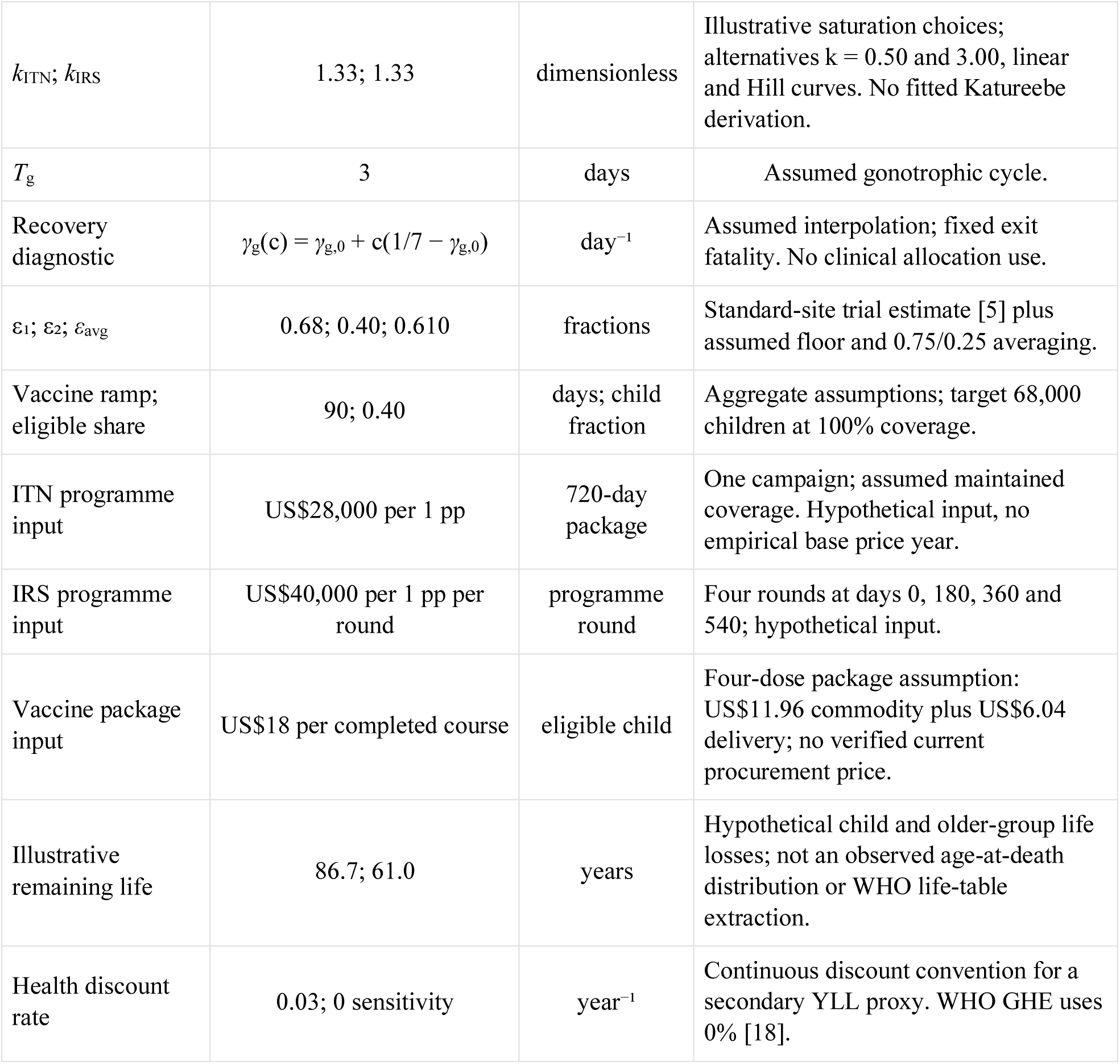
Full reference parameter dictionary and evidence basis.

**Table A2.**
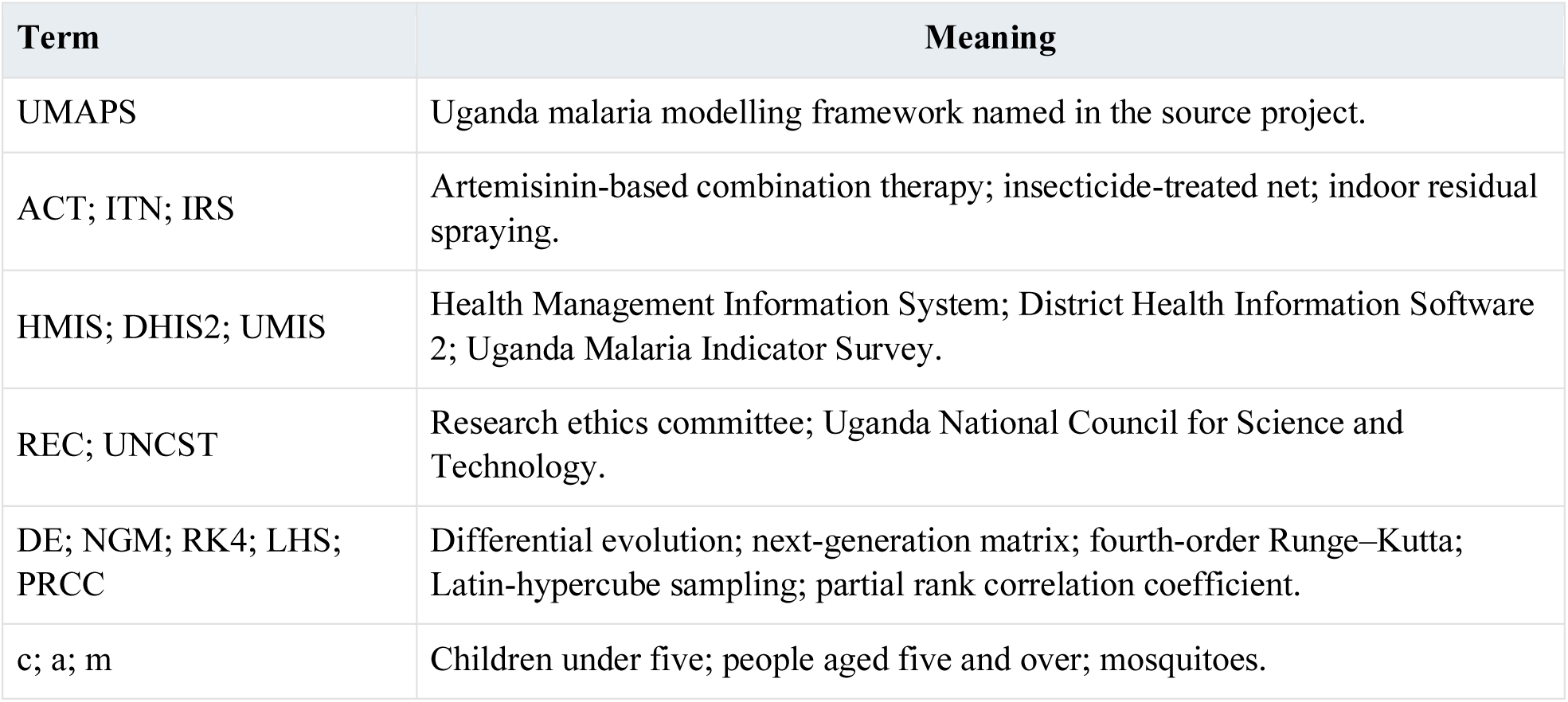
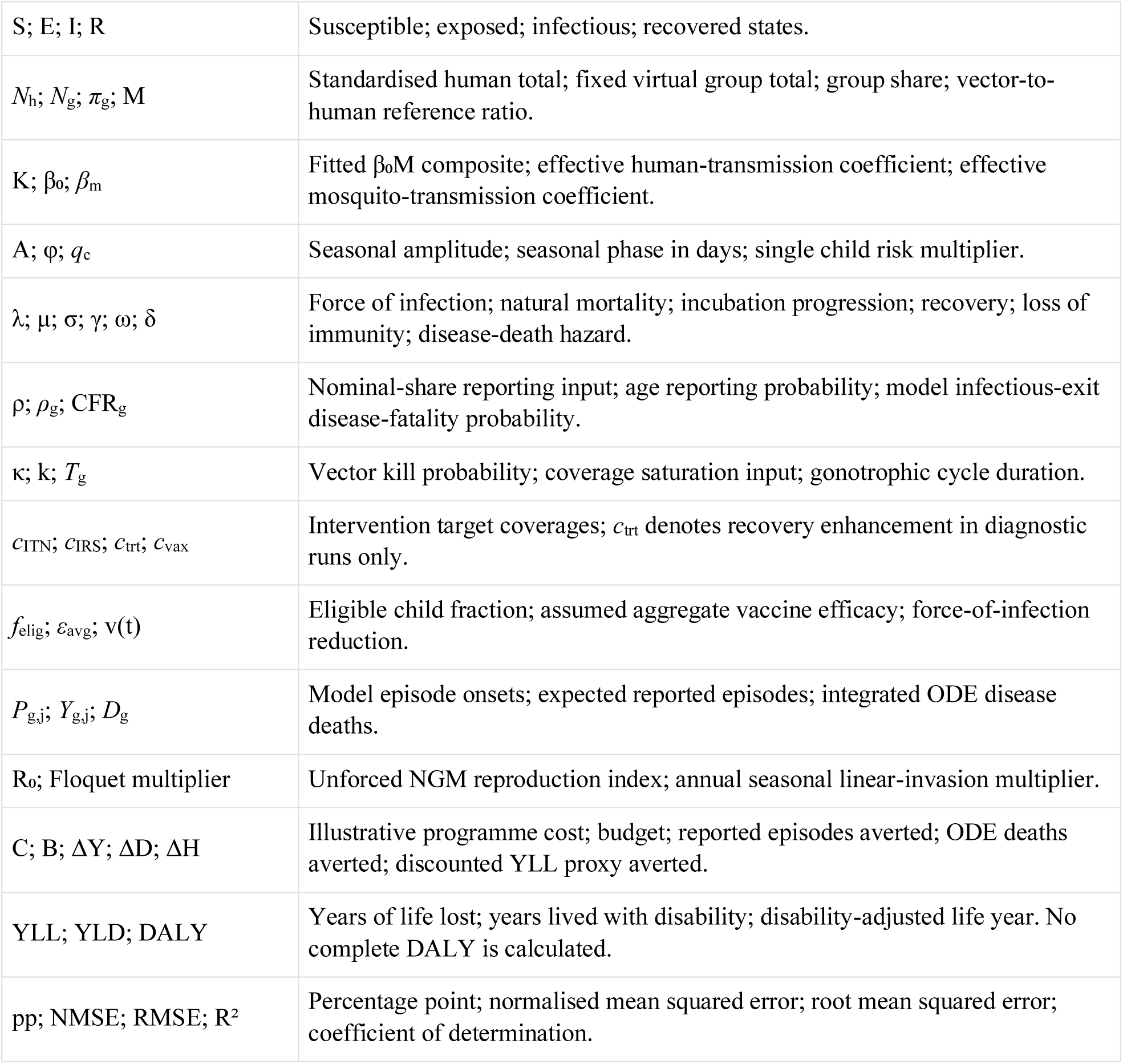
Abbreviations and mathematical symbols.

## Appendix B Executed verification checks

The 17 assertions were executed against numerical reference version 0.5 on 11 October 2026. They concern the stated mathematical and computational implementation. Tolerances and detailed values are retained in results/verification.json. They are not clinical validation or independent expert endorsement.

**Table B1.**
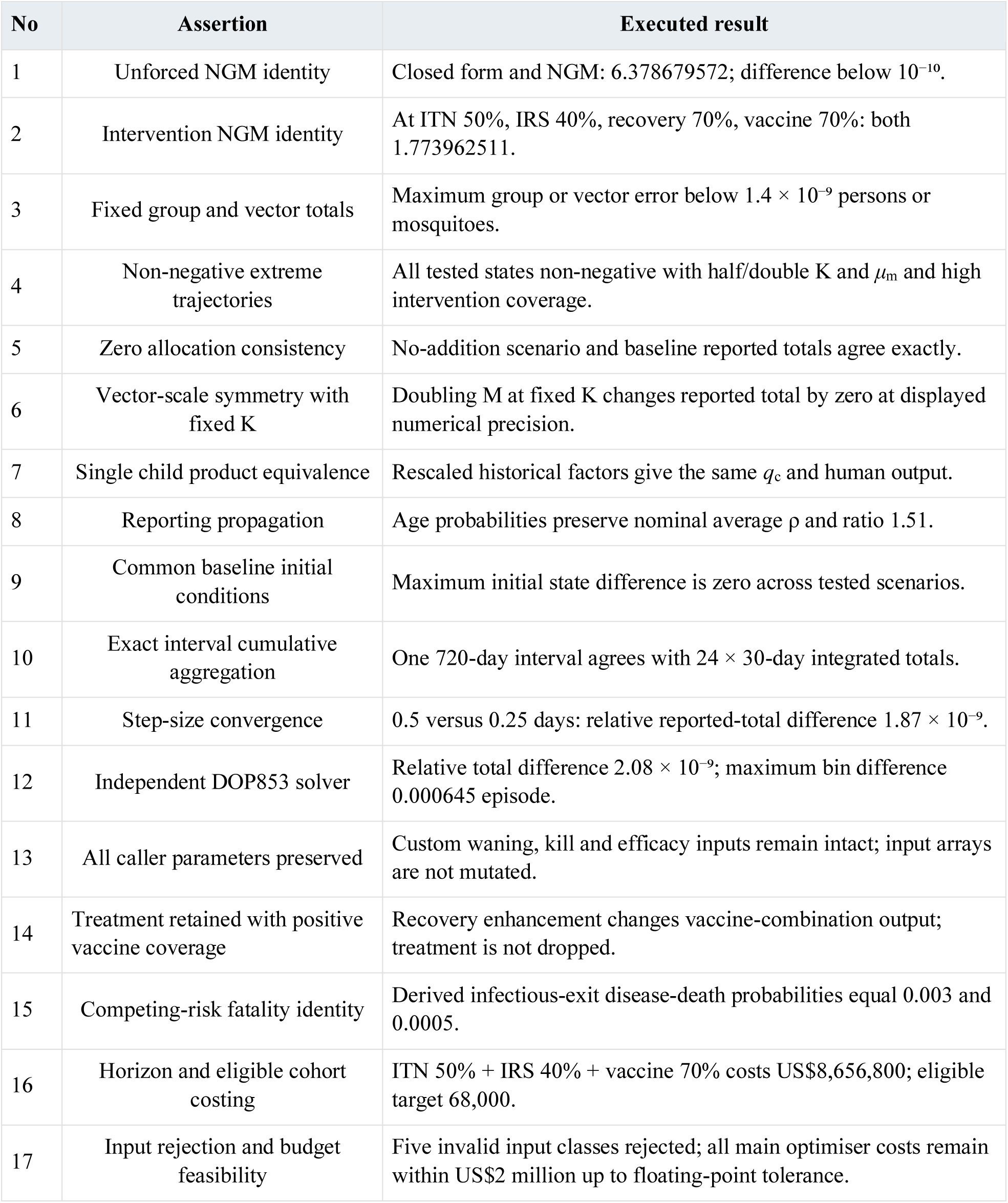
Mathematical and numerical assertions.

## Appendix C Reproduction and empirical data requirements

The supporting archive contains analysis_release/core.cpp, model.py, run_analysis.py, checks.py, supplement.py and reproduce_all.py; results/ contains full calibration, scenarios, seed searches, profiles, ensembles, noise replicates, sensitivity and structural alternatives. Inputs, exact dependency pins, compiler and platform information, licence and hash manifests are supplied. Authoring files contain this manuscript builder, its figure sources and the protocol reference needed to reproduce the editable equations and tables.

With Python 3.12, the recorded NumPy and SciPy dependencies and a C++17 compiler, run python reproduce_all.py from analysis_release to execute calibration, checks and supplementary diagnostics in order. Run python reproduce_all.py --verify-existing to check supplied output hashes and assertions. The platform-specific shared core is compiled locally. Small floating-point or search differences on another platform should be assessed against the documented tolerances, rather than treated as a changed scientific method.

The main reference analysis ran in Python 3.12.14 with NumPy 2.3.5 and SciPy 1.17.0, Linux x86_64 and a C++17 compiler. Its recorded runtime was 348.8 seconds in that environment. This is a single recorded run, not a benchmark of the historical dashboard, the author’s computer or another optimiser. The unchanged synthetic input SHA-256 is:

`e34443d00b7ffa618a0f2144f76c3c118905147ad6ca4d933b265724954f83db`

**Table C1.**
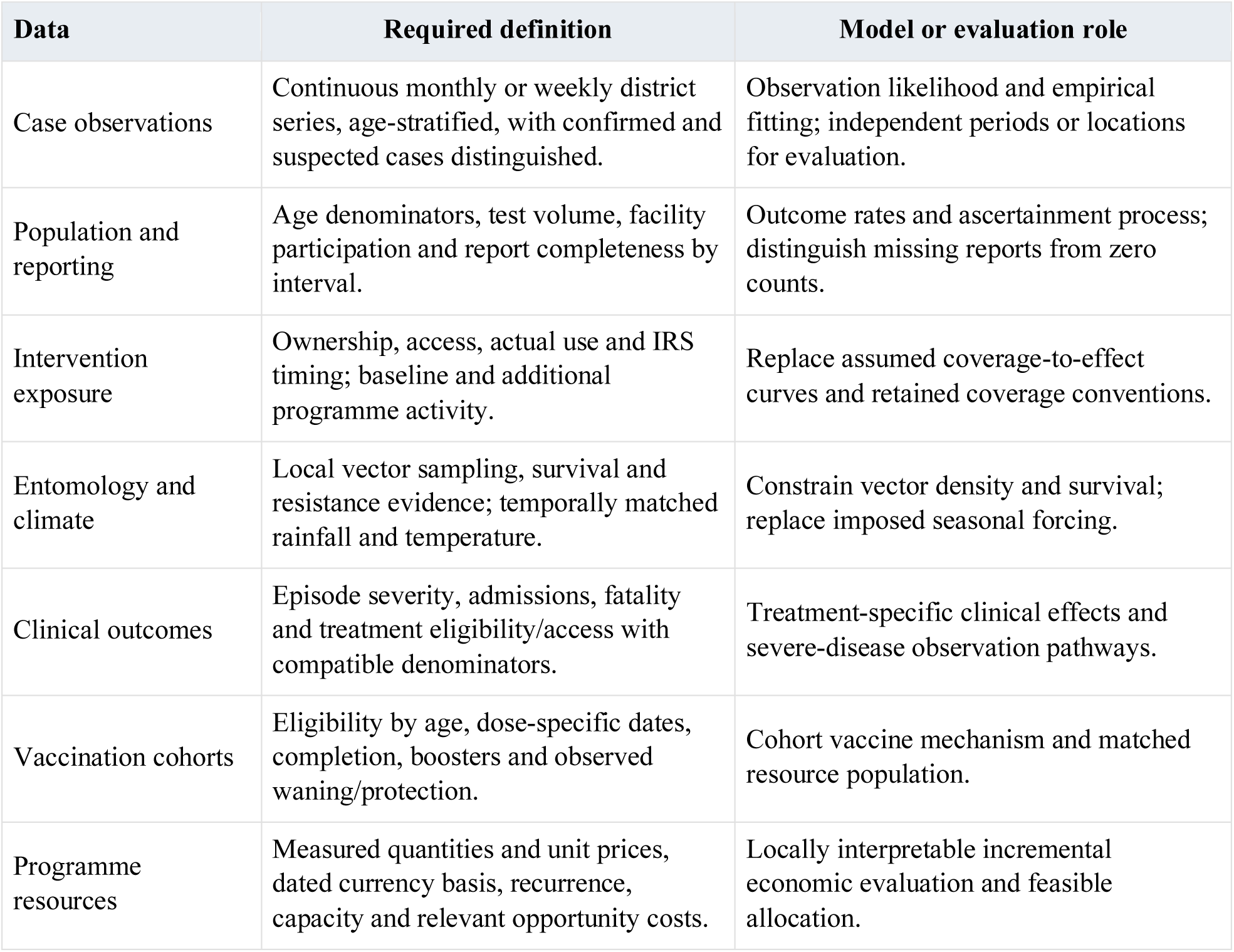
Data needed for an empirical extension.

| Data | Required definition | Model or evaluation role |
| --- | --- | --- |
| Case observations | Continuous monthly or weekly district series, age-stratified, with confirmed and suspected cases distinguished. | Observation likelihood and empirical fitting; independent periods or locations for evaluation. |
| Population and reporting | Age denominators, test volume, facility participation and report completeness by interval. | Outcome rates and ascertainment process; distinguish missing reports from zero counts. |
| Intervention exposure | Ownership, access, actual use and IRS timing; baseline and additional programme activity. | Replace assumed coverage-to-effect curves and retained coverage conventions. |
| Entomology and climate | Local vector sampling, survival and resistance evidence; temporally matched rainfall and temperature. | Constrain vector density and survival; replace imposed seasonal forcing. |
| Clinical outcomes | Episode severity, admissions, fatality and treatment eligibility/access with compatible denominators. | Treatment-specific clinical effects and severe-disease observation pathways. |
| Vaccination cohorts | Eligibility by age, dose-specific dates, completion, boosters and observed waning/protection. | Cohort vaccine mechanism and matched resource population. |
| Programme resources | Measured quantities and unit prices, dated currency basis, recurrence, capacity and relevant opportunity costs. | Locally interpretable incremental economic evaluation and feasible allocation. |

## Appendix D Supplementary figures

These figures provide additional visual diagnostics for numerical reference 0.5, covering the stored input, model dynamics, intervention assumptions, reproduction, vaccine coverage, age-specific effects and assumed parameter trajectories. They use the same equations, fitted vector and uncertainty draws as the main analysis.

**Figure D1.**
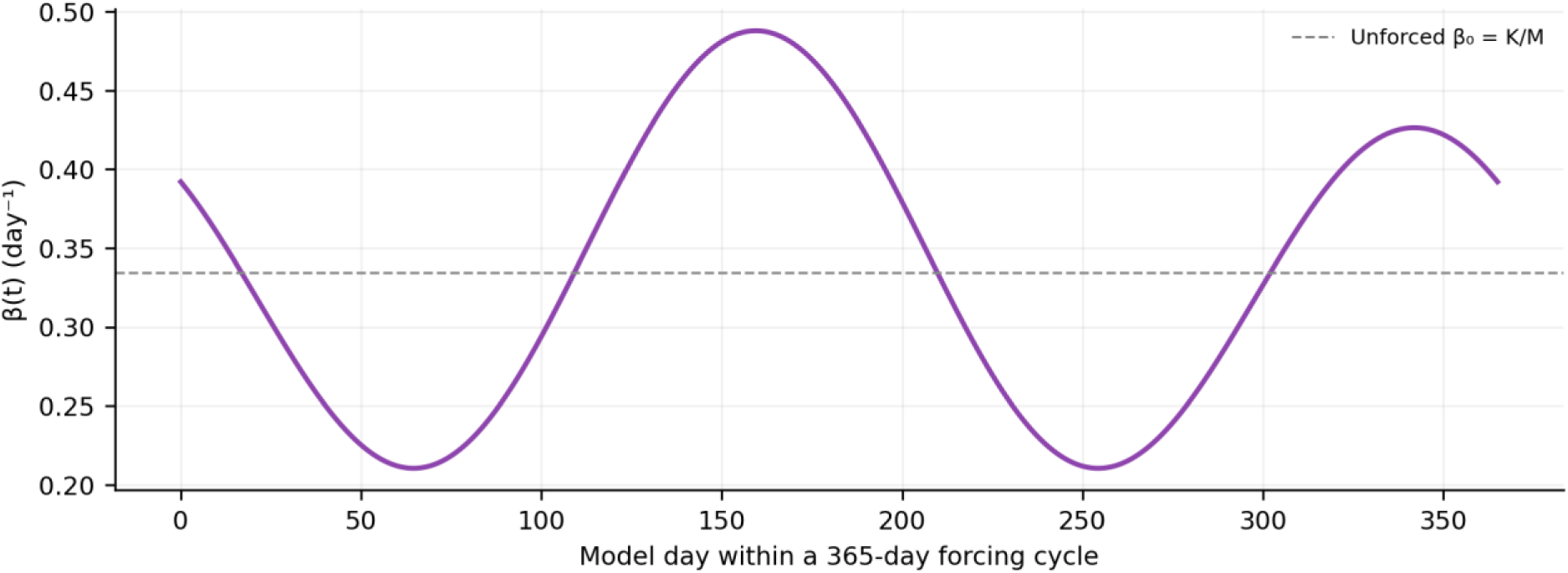
Fitted seasonal forcing over one 365-day cycle. The unforced coefficient is K/M with the reference scale M = 3.5. The harmonic shape and phase anchor are model assumptions; the curve is not an observed rainfall series.

**Figure D2.**
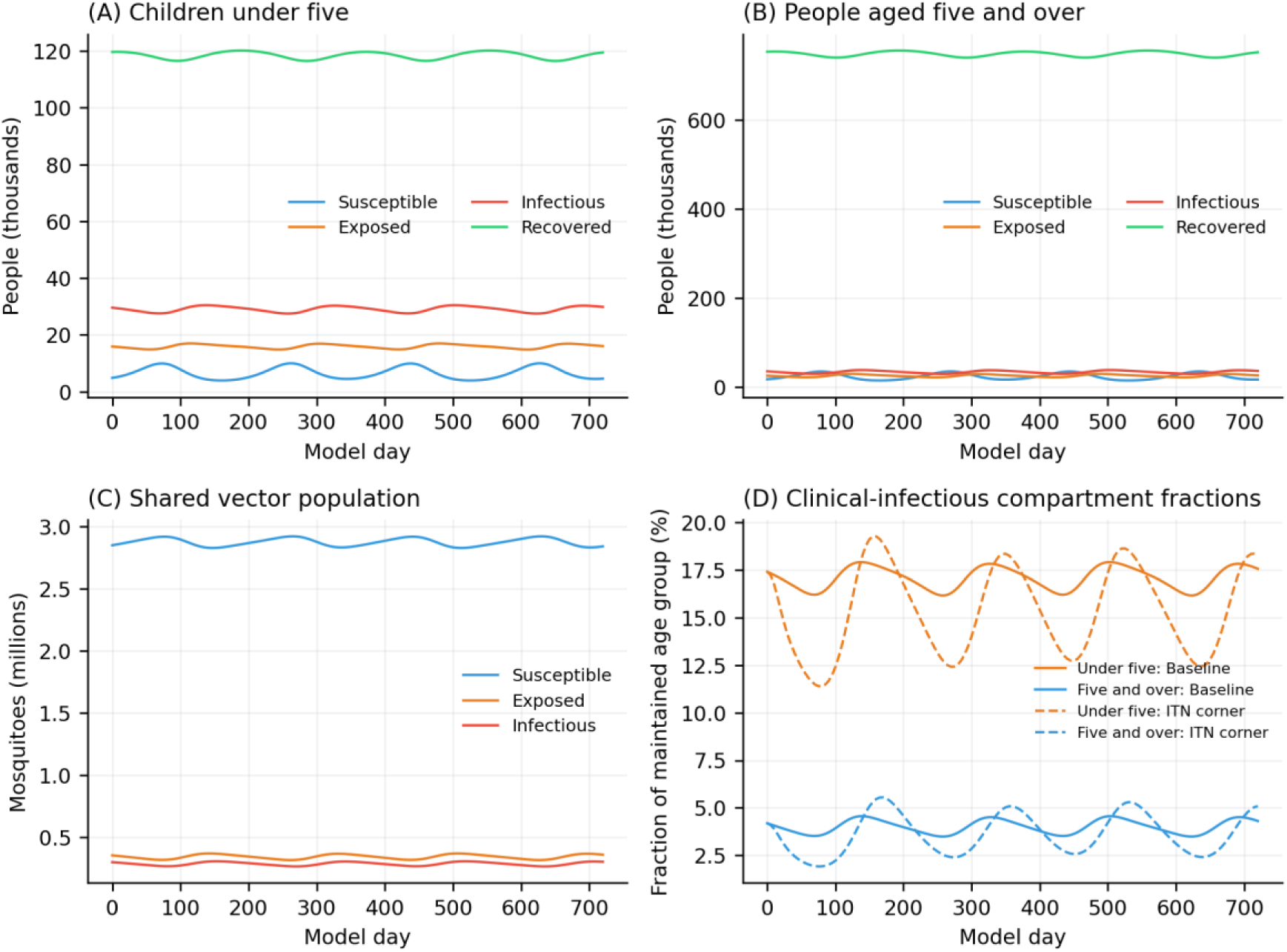
Common-baseline compartment dynamics over 720 days. Panels A–C show maintained human and vector counts. Panel D compares age-specific clinical-infectious fractions for baseline and the selected ITN corner. These fractions are not microscopy prevalence or laboratory-confirmed disease.

**Figure D3.**
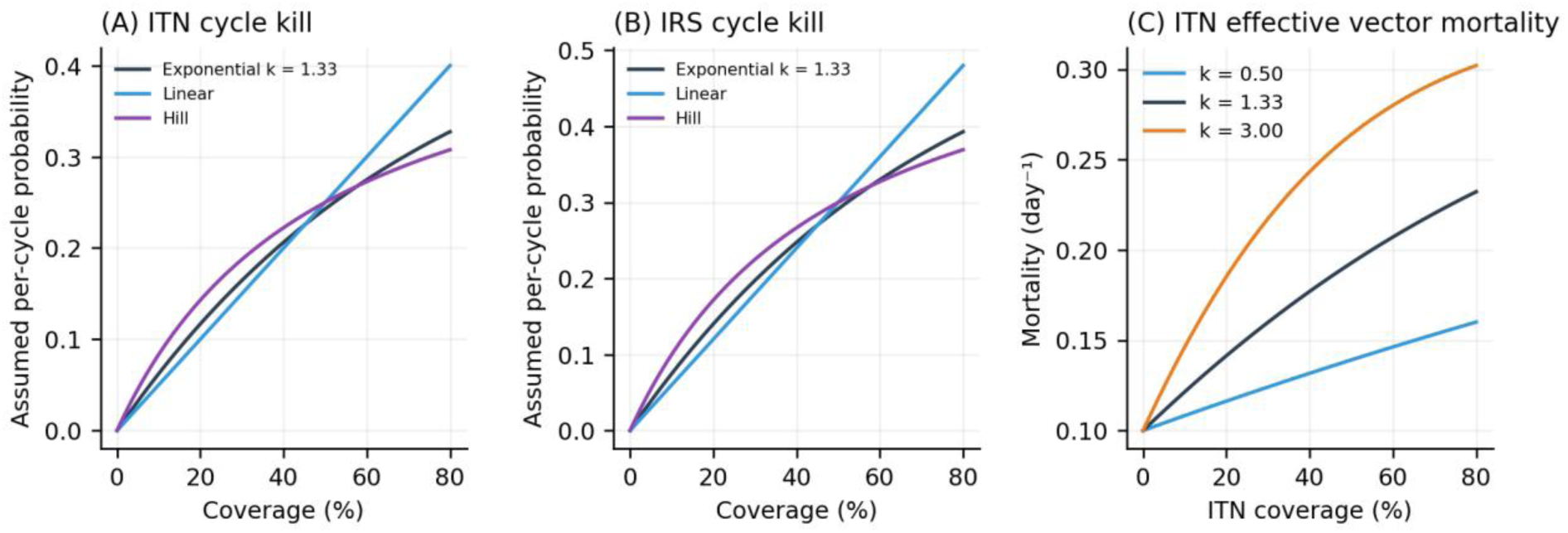
Assumed vector response functions. Exponential, linear and Hill curves show per-cycle kill probabilities, while the right panel shows effective mosquito mortality under three exponential saturation inputs. All curves are assumed response functions and are not fitted to field observations.

**Figure D4.**
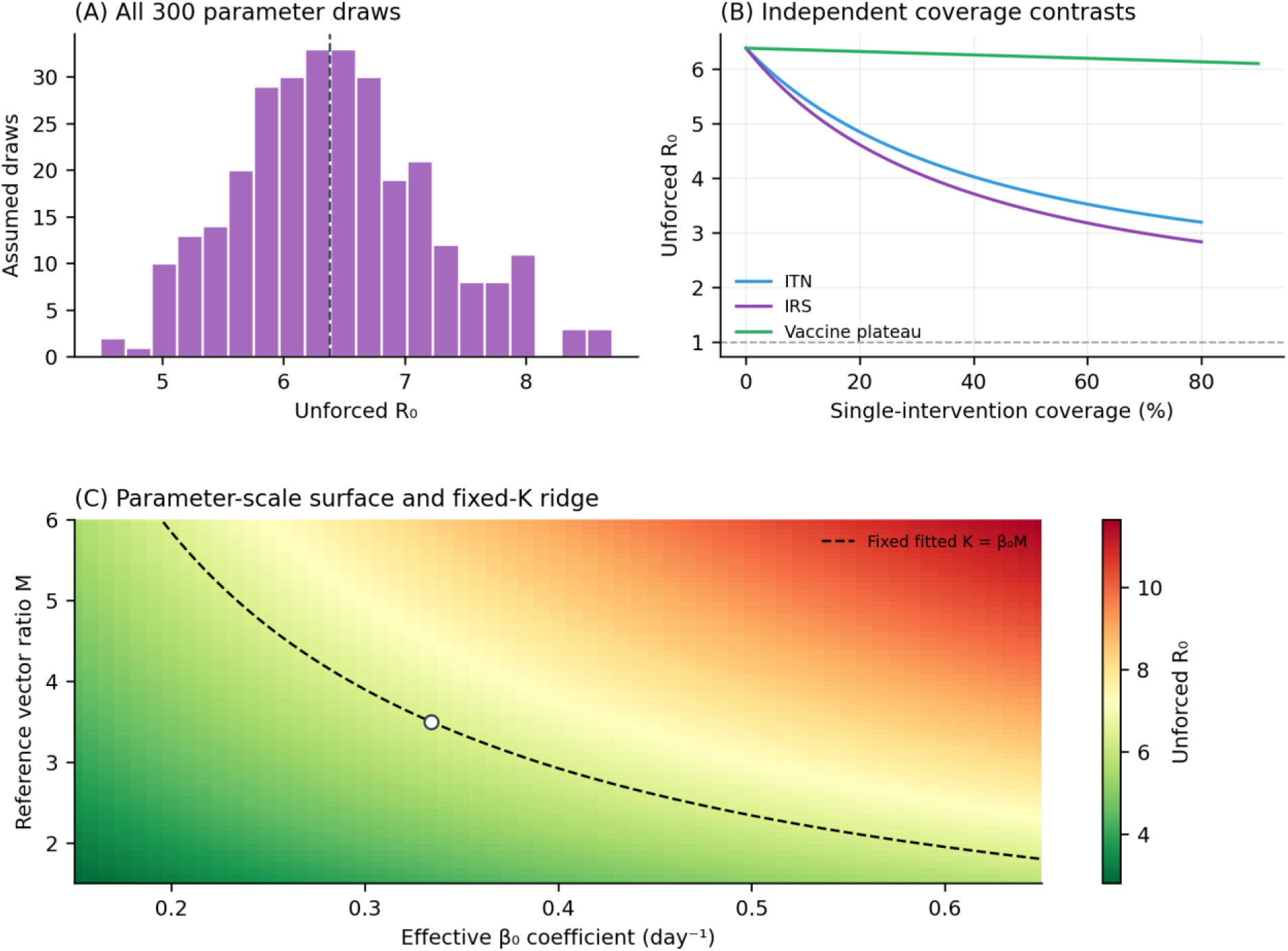
Reproduction diagnostics. Panel A uses all 300 assumed parameter draws; panel B varies one prevention coverage at a time, using vaccine plateau protection. Panel C varies the effective coefficient and reference density with their product defining K; the dashed fixed-K ridge has identical human-transmission scale. All panels concern unforced R₀, not the seasonal Floquet multiplier or an empirical elimination forecast.

**Figure D5.**
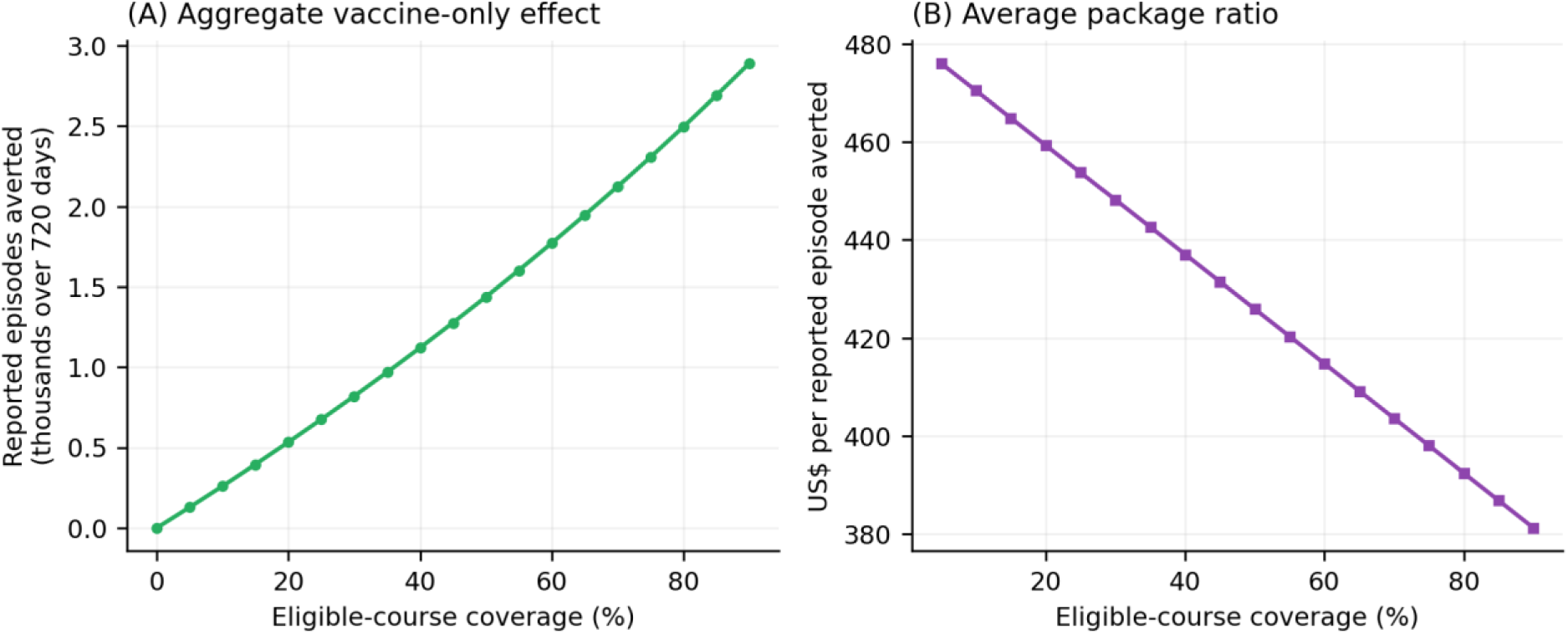
Vaccine-only coverage sensitivity under the current aggregate mechanism. Reported episodes averted and average additional-package ratios use the same 720-day baseline, eligible stock and course input. Zero-coverage benefit is zero and its ratio is undefined. No individual efficacy-waning curve or clinical effectiveness estimate is inferred.

**Figure D6.**
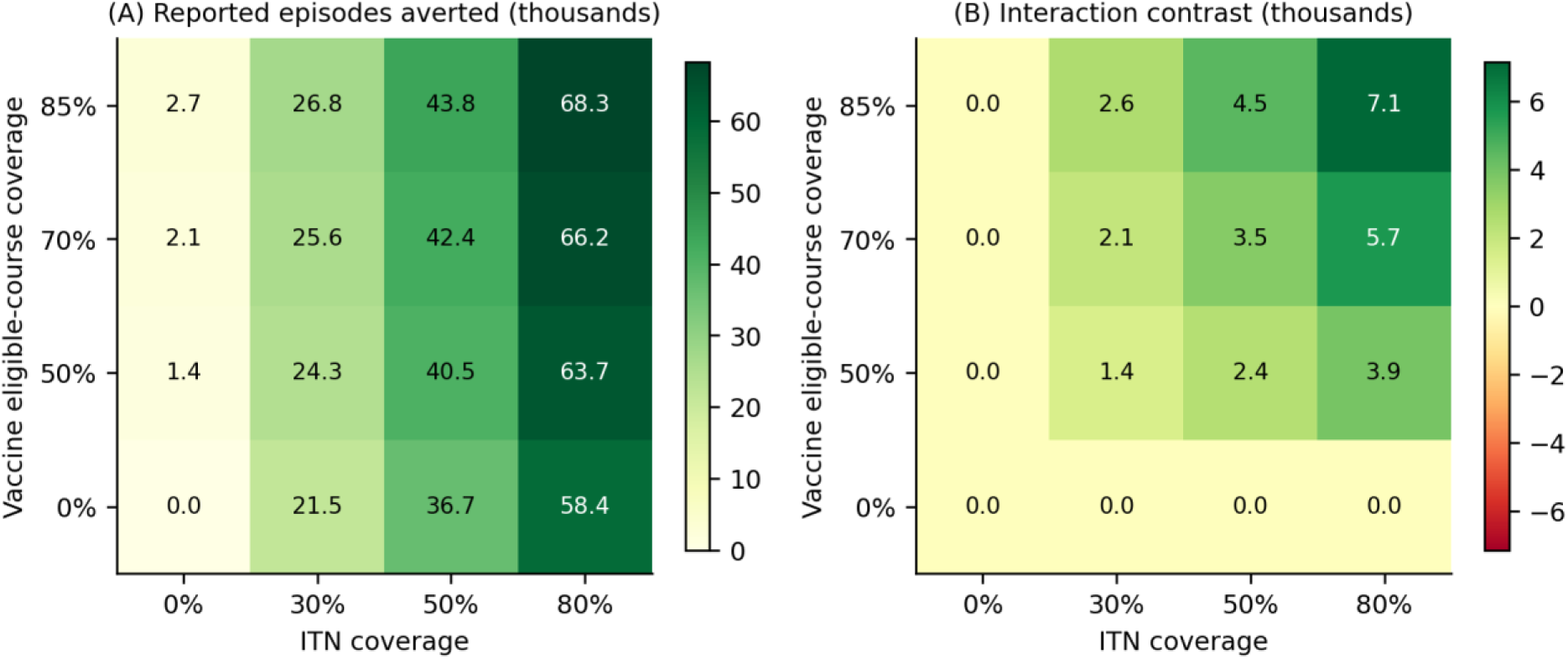
ITN–vaccine coverage grid with zero IRS and recovery enhancement. The right panel is combined reported benefit minus the two separate benefits against the common baseline. It is a model interaction contrast, without a claim of empirical biological synergy. The ITN 50% and vaccine 70% cell gives 42,351.26 reported episodes averted and an interaction of +3,536.06.

**Figure D7.**
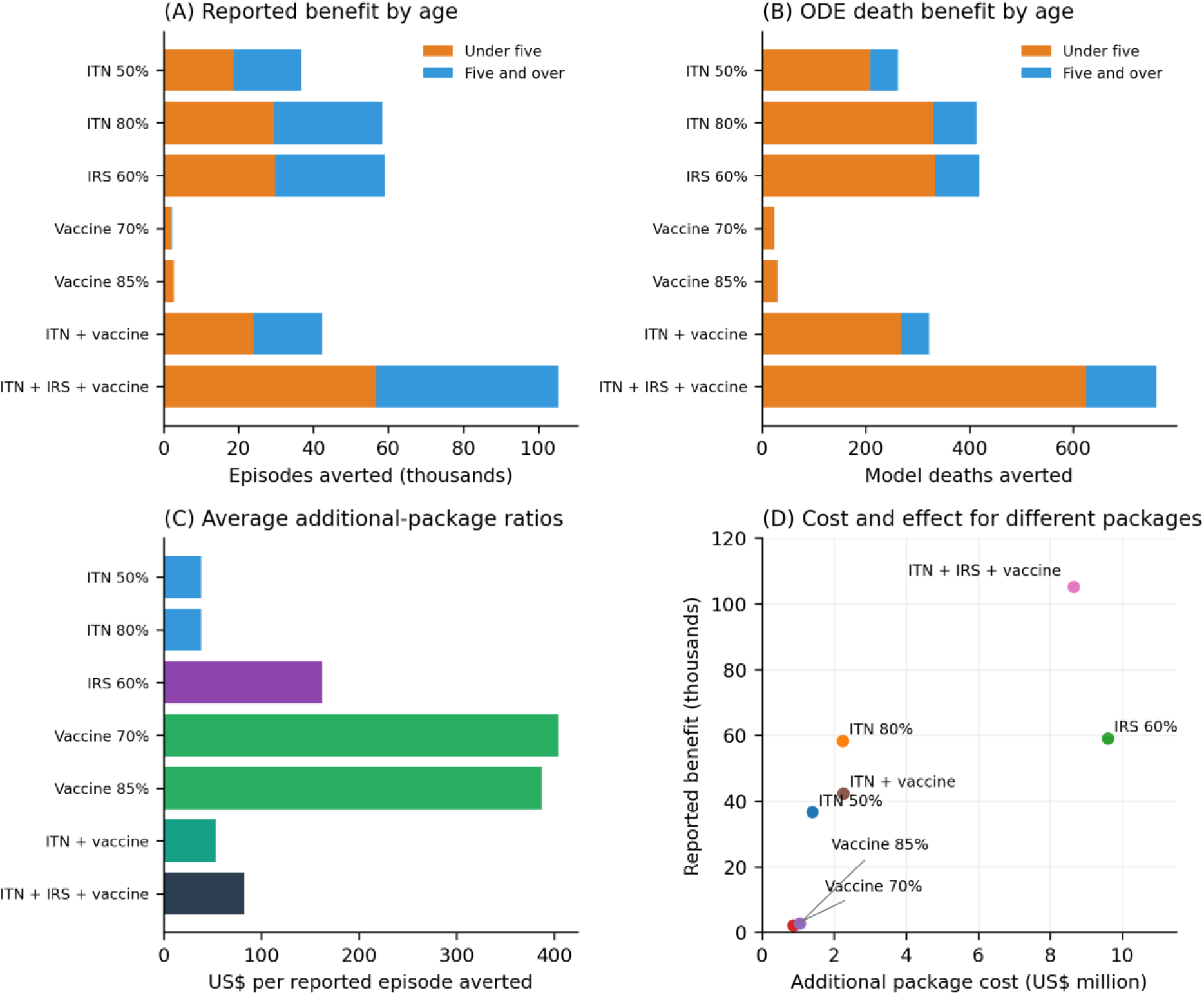
Age-specific reported-episode and ODE-death benefits and illustrative package costs. All packages use the common 720-day baseline. Ratios are average comparisons against zero additional spending. The cost–effect scatter is not an incremental efficiency frontier and includes no adoption threshold. ITN, IRS and vaccine percentages match Table 3.

**Figure D8.**
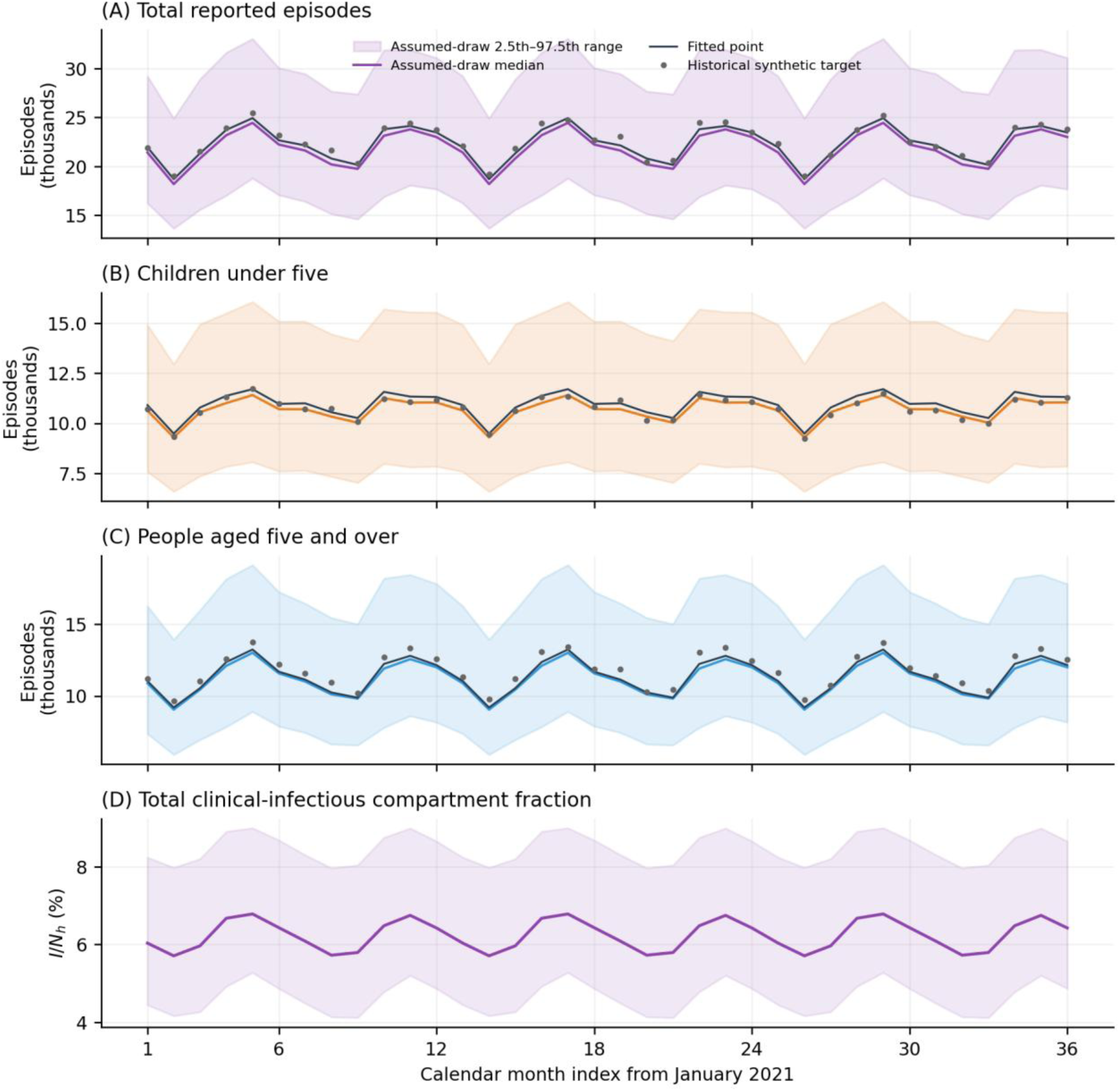
Complete 300-draw monthly trajectory summaries over the 36 calendar months. Bands are pointwise 2.5th–97.5th percentiles under the stated assumed distribution, not empirical confidence limits or fitted posterior intervals. The historical target and fitted point are plotted separately. Panel D shows the total clinical-infectious fraction at month ends; this compartment measure is distinct from survey positivity. Calendar intervals here differ from the 720-day prevention horizon.

