## Supplementary material for "Malaria transmission modelling and conditional prevention allocation in Uganda": Protocol

### Malaria intervention modelling and illustrative budget allocation in Uganda

Retrospective computational research protocol

An age stratified SEIR SEI framework with reconciled synthetic calibration and a vaccine transmission module

Jay Hiteshkumar Patel – Author

John Mulangwa – Co-author

Milton Musaba – Co-author

Butisema University, Kampala, Uganda

Version 0.5
11 October 2026

This protocol records completed computational work and the reconciled reference analysis executed on 11 October 2026. It specifies the equations, observation rule, scenario estimand, illustrative programme budget, uncertainty analyses and reproducibility package for this version. It is retrospective and has no claim of prospective registration.

The study examines model behaviour in a standardised population. Its fitted parameters and portfolios are conditional computational results. They do not establish a validated national programme allocation or the clinical effect of changing malaria treatment.

### Contents

1 Administrative information
2 Structured summary
3 Background and rationale
4 Objectives and research questions
5 Study design and workflow
6 Setting population and estimands
7 Data and observation definitions
8 Model specification
9 Parameters and provenance
10 Calibration and identifiability
11 Scenarios economics and optimisation
12 Uncertainty sensitivity and evaluation
13 Software and reproducibility
14 Limitations
15 Ethics governance and data protection
16 Dissemination and future work
References
Appendix A Abbreviations and symbols
Appendix B Mathematical and numerical checks
Appendix C Reporting crosswalk
Appendix D Reproducibility package
Appendix E Amendments and deviations

### 1 Administrative information

| Item | Specification |
|---|---|
| Protocol identifier | UMAPS protocol version 0.5 |
| Author and study responsibility | Jay Hiteshkumar Patel, independent researcher, Kampala, Uganda |
| Correspondence | |
| Version date | 11 October 2026 |
| Study type | Retrospective deterministic modelling study with synthetic calibration and exploratory sensitivity analyses |
| Setting | Illustrative homogeneous Uganda transmission zone; standardised population of 1,000,000 |
| Primary computational outcome | Model-reported episode onsets averted during 720 days from a common baseline state |
| Registration and ethics record | No prospective registration, approval or exemption determination is claimed |
| Reference implementation | UMAPS 0.5 source and numerical outputs accompany this revision in the supporting reproducibility archive |
| Financial and conflict declarations | Funding, sponsor involvement and competing interests were not specified in the available author records; no absence of funding or conflicts is inferred |

This version supersedes the numerical specifications in protocol 0.4 for the present analysis. The historical manuscript and dashboard archive remain source records. Current tables were computed with the reference implementation described here; historical output values are not relabelled as current results. Appendix E records the substantive changes.

### 2 Structured summary

##### Purpose

The study assesses how a malaria transmission model responds to intervention, observation and costing assumptions, and whether its calculations can be reproduced. It addresses parameter confounding, recovery and immunity cycling, aggregate child risk, prevention allocation, vaccine feedback and sensitivity to uncertainty. The aim is a transparent methodological demonstration for subsequent empirical work.

##### Design and methods

The human model has susceptible, exposed, infectious and recovered states in two virtual age groups, linked to susceptible, exposed and infectious mosquitoes. Group totals are held fixed by compensating recruitment. Calibration uses 36 months of historical synthetic observations, one deterministic observation rule and four fitted composite or observation parameters. A separate noiseless fixture tests recovery under the reconciled equations. Every scenario starts from the same phase-aligned baseline periodic state and runs for 720 days.

ITNs and IRS increase vector mortality through stated assumed kill curves. The vaccine reduces the force of infection in children and therefore participates in human–mosquito feedback. It is an aggregate leaky-protection approximation. A recovery-only treatment experiment is analysed separately and excluded from prevention budget selection, because the model does not represent treatment-specific reductions in clinical severity or fatality.

##### Executed reference results

The reconciled fit gives total-series $R^2$ 0.9537, monthly RMSE 379.66 and correlation 0.9870. The baseline produces 528,041 model-reported episodes over 720 days. The unforced next-generation reproduction index is 6.3787; its closed form agrees with the numerical matrix calculation. These are synthetic and mathematical checks, not external clinical validation.

At a hypothetical US$2 million additional prevention budget, the searched base-cost portfolios select ITN coverage 71.43%, with IRS and vaccine additions zero to reported precision. The boundary benchmark gives 52,483 reported episodes averted. The result depends on the assumed intervention functions, four IRS rounds, aggregate vaccine mechanism and unit costs. It provides no basis for withdrawing existing vaccination, vector control or essential case management.

The analysis includes five calibration seeds, five seeds for each main budget search, a 10,201-point grid benchmark, all 300 assumed parameter draws across 115 candidate portfolios, 40 synthetic noise realisations and 500 Latin-hypercube samples. All 17 mathematical and numerical assertions passed. No independent clinical validation or public release deposit is claimed.

#### 3 Background and rationale

##### Public health context

Malaria remains an important health burden in Uganda, with substantial geographical variation in transmission and surveillance performance [1,6,7]. National-average models can show the implications of explicit assumptions. Local programme planning also requires district epidemiology, access to care, delivery constraints and resource evidence that this illustration does not contain.

Uganda launched routine R21/Matrix-M vaccination on 2 April 2025, initially targeting approximately 1.1 million children in 105 high- and moderate-transmission districts, with doses at 6, 7, 8 and 18 months [2]. The phase 3 trial reported 12-month efficacy of 75% at seasonal sites and 68% at standard sites [3]. The present model uses the standard-site estimate as one component of an explicitly assumed aggregate protection input. It does not reproduce the trial dose histories or estimate Ugandan programme effectiveness.

##### Scientific rationale

Interventions change different parts of the human–mosquito transmission cycle. Their apparent ranking can also change when the observation model, population dynamics or cost horizon changes. A strong synthetic fit is insufficient if several parameter combinations generate the same trajectory or if the outcome used in optimisation does not represent the clinical objective. The study therefore evaluates the equations and computational pathway alongside scenario outputs.

The SEIR–SEI architecture follows established malaria modelling approaches [11,14]. The contribution of this work is its explicit integration of calibration, identifiability diagnostics, common-state scenarios, prevention budget search and reproducible sensitivity calculations. The model is deliberately small so that structural assumptions and their effects can be examined directly.

##### Retrospective chronology

Model development and the historical analyses preceded the original protocol. The reconciled analyses in this version were undertaken after the historical data and results were available. They are retrospective exploratory work. Synthetic thresholds, ranges and search settings have no status as prospectively registered success criteria [4,5]. The clean protocol describes the implemented version and preserves this chronology in Appendix E.

#### 4 Objectives and research questions

The general objective is to evaluate the reproducibility and inferential limits of an integrated malaria modelling framework in an illustrative Uganda setting. The four original research questions are retained, with their operational form aligned to what the equations can answer.

**Table 1. Research questions and implemented analyses**

| Question | Implemented analysis | Scope of inference |
|---|---|---|
| Q1 Separate transmission and vector density | Fit $K = \beta_0 M$; test the vector-scale symmetry; examine composite-loss profiles and a noiseless round trip. | Monthly human outputs cannot identify $\beta_0$ and M separately under the stated recruitment convention. |
| Q2 Recovery and episode cycling | Compare fixed recovery enhancement with the common baseline; vary immunity waning. | Describe behaviour of the recovery-only equations; no treatment benefit or harm estimate is inferred. |
| Q3 Child exposure and susceptibility | Use the single multiplier $q_c = \alpha_c \varphi_c$; demonstrate product equivalence. | The model identifies no separate exposure and immunological mechanisms. |
| Q4 Prevention portfolios and vaccine contribution | Run common-state scenarios, constrained budget searches, finite-pool ensemble selection and structural cost alternatives. | Conditional prevention portfolios within the model and declared cost assumptions. |

Additional objectives are to examine parameter sensitivity, separate noise realisations from search randomness, assess mathematical and numerical implementation, document the role of external aggregate sources, and provide an executable source package. The primary allocation objective is cumulative model-reported episode onsets averted. Model episode onsets before reporting, age-specific outputs, ODE disease deaths, a discounted YLL proxy and search stability are secondary or diagnostic outcomes.

There is no participant sample, recruited comparison group or confirmatory hypothesis test. The computational sample sizes are 36 observation intervals, five main search seeds, 300 assumed parameter draws, 40 noise realisations and 500 sensitivity samples. These counts define diagnostic precision and computational coverage; they are not epidemiological sample-size calculations or a power analysis.

### 5 Study design and workflow

The workflow begins with an explicit model and parameter dictionary. The historical synthetic observation fixture is retained unchanged. The same deterministic forward solver and age-specific reporting probabilities are used for fitting and evaluation. Calibration output is then passed intact to the scenario, intervention and budget routines. Figure 1 shows the principal dependencies.

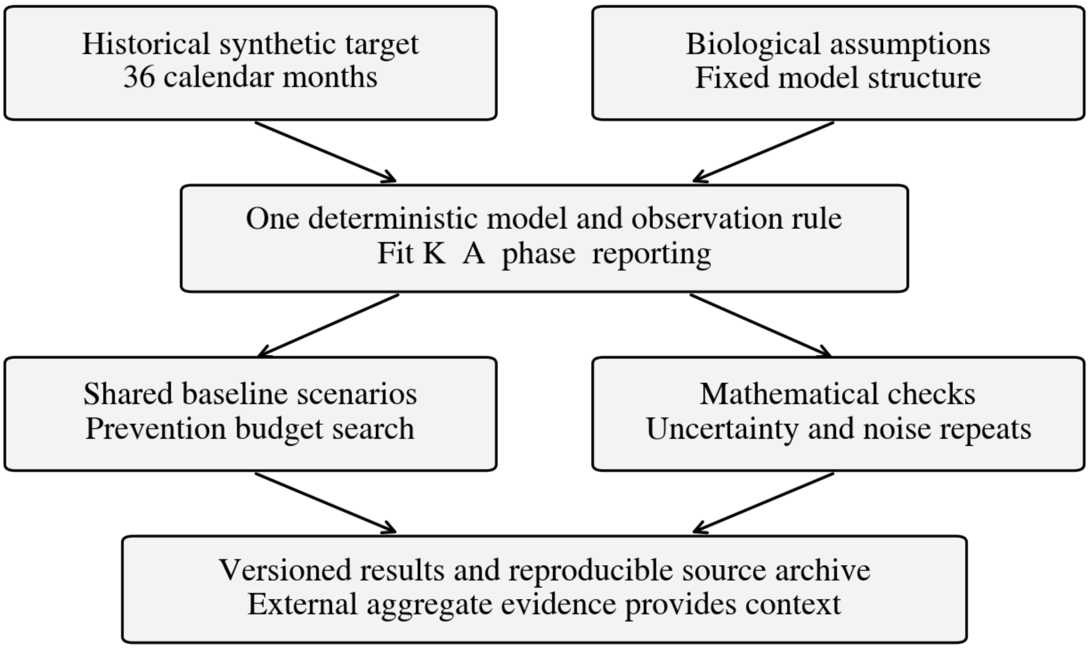


**Figure 1. Computational workflow for version 0.5. External aggregate evidence supplies context; it is not included in the synthetic fitting objective or treated as a held-out validation set.**

The baseline periodic state is computed before intervention scenarios. Scenario effects are compared over identical intervals and initial conditions. Budget search is evaluated across seeds and against explicit feasible boundaries. Parameter ensembles, structural alternatives and repeated observation-noise experiments are reported separately so that their distinct meanings remain visible. Source, input and result hashes identify the calculations used in the final tables.

### 6 Setting population and estimands

#### Setting and virtual population

The model represents one homogeneous mixing zone with $N_h$ = 1,000,000. Children under five account for $\pi_c$ = 0.17 and people aged five and over for $\pi_a$ = 0.83. The symbol a denotes the latter group and includes school-age children. Its outputs must not be described as exclusively adult outcomes. These fixed shares standardise the illustration and are not estimates obtained from a verified census table.

Each virtual age-group total is maintained by compensating recruitment for natural and disease deaths. No individual ageing process is modelled. Recruitment into the older group is a mathematical replacement flow, not a literal adult birth process. The model therefore represents two maintained populations with different natural-history assumptions. Prevalence is calculated using their actual, conserved totals of 170,000 and 830,000.

##### Model boundary

The model omits spatial movement, pregnancy, household exposure, asymptomatic and submicroscopic states, diagnostic test characteristics, severe disease, admission, drug resistance and mosquito species or insecticide resistance. Recovered individuals return to full susceptibility at exponential rates. Vector recruitment maintains M $N_h$ even when mortality increases, so vector control changes survival and infectious composition without an independently estimated fall in vector density.

##### Scenario estimand

The principal contrast is an implementation counterfactual in the synthetic system. Additional intervention coverage begins at model day zero from a common baseline state and seasonal phase. The comparator uses zero additional intervention coverage with the stated baseline recovery rates. It does not represent withdrawal of existing health services or an estimate of the actual untreated Ugandan population.

Effects are evaluated over T = 720 days in 24 consecutive 30-day intervals. This is not two calendar years. Calibration instead uses the actual boundaries of January 2021 to December 2023, totalling 1,095 days. Both use the same differential equations, RK4 operator and reporting function. Time is measured in days; periodic forcing has a 365-day period. No result is interpreted as a forecast of the present 2026 population.

#### 7 Data and observation definitions

##### Historical synthetic observations

The unchanged input file uganda_synthetic_monthly.csv contains 36 rows. It provides calendar labels, compartment summaries, prevalence labels, transmission summaries, infection-acquisition fields and synthetic reported counts by age. The fitted target comprises reported_cases_children and reported_cases_adults; the latter column is interpreted as the group aged five and over. The filename and original labels are preserved for traceability rather than rewritten in place.

The historical solver counted infection acquisitions $\lambda_g S_g$ at a step boundary. Version 0.5 defines episode onset as progression $\sigma_h E_g$ into the infectious state and integrates that flow within RK4. The historical population and reporting paths also differed from the present implementation. Consequently, refitting the unchanged synthetic target assesses compatibility with a historical fixture; it is not recovery of the historical generating $\beta_0$ and M values. A separate noiseless fixture generated by version 0.5 tests same-model recovery.

Synthetic observation randomness belongs to the original fixture. There is no new noise inside the fitting objective, fitted output or intervention simulator. The repeated-noise analysis generates separate noisy monthly targets from the fitted deterministic expectation and records a distinct random seed for each realisation. It does not change the forward equations or introduce solver-step-dependent observation variance.

#### External aggregate evidence

**Table 2. External sources and their role in the present study**

| Source | Outcome and scope | Role and restriction |
|---|---|---|
| Aregawi and colleagues [6] | Uganda national routine-service indicators and the 2022 epidemic. | Epidemiological context. Confirmed, outpatient and inpatient series are not interchangeable with model infectious onsets. |
| Zalwango and colleagues [7] | HMIS severe malaria cases, 2017–2021. | Context for disease severity and surveillance. No severe-disease compartment is fitted. |
| UMIS 2024–25 [8] | Survey infection testing in children aged 6–59 months and intervention coverage indicators. | Context for measurement and coverage. Microscopy positivity is not clinical-stage prevalence $I_c/N_c$; ownership, access and use have different denominators. |
| Historical weekly compilation | 29 selected weekly records across 2020, 2021, 2023 and 2024; original bulletin PDFs were unavailable in the accessible source package. | Excluded from formal external validation and numerical significance claims. The years and epidemiological weeks are retained as source metadata only. |
| WHO reports [1] | Estimated national malaria burden using WHO methods. | Burden context. Estimates are not substituted for laboratory-confirmed HMIS cases or the model reporting process. |
| Cost-of-illness studies [30,31] | Uganda illness costs with different dates and care definitions. | Evidence context for future economic input collection; no flat per-model-episode cost is imported. |

No compatible independent observation set was available for empirical calibration or prediction assessment. Seasonal similarity partly reflects an imposed forcing shape, and annual rescaling can conceal outcome mismatches. The present protocol therefore omits the historical claims of four independent validation layers, nominal correlation p values and biological calibration from pooled weekly records. No hold-out performance is asserted.

#### Observation rule and missingness

The age-specific expected reported episode count in interval j is the integral of $\sigma_h E_g$ multiplied by $\rho_g$. The fitted reporting parameter ρ is a nominal population-share average, with a fixed child-to-older ratio $r_\rho = 1.51$. Both probabilities are derived anew for every parameter vector. They are not kept at historical hard-coded fractions when ρ changes.

$$\rho_a = \frac{\rho}{1.51\,\pi_c + \pi_a}, \quad \rho_c = 1.51\,\rho_a \qquad (1)$$

At the fitted point, $\rho_c = 0.271150$ and $\rho_a = 0.179570$, while ρ = 0.195138. The baseline episode-weighted reported fraction is 0.215204; it differs from the nominal population-weighted value because episode incidence differs by age. The reporting parameters are model mappings, not independently observed ascertainment probabilities.

The 36 fitted observation rows are complete and no imputation is performed. Future empirical work must distinguish zero cases, a missing report and a reported aggregate without known completeness. Reporting completeness, test volume and facility coverage would require their own observation model. Missing values must never be coded as zero merely to obtain a full time series.

### 8 Model specification

##### Compartments and transmission

For g ∈ {c,a}, human states are $S_g$, $E_g$, $I_g$ and $R_g$. Mosquito states are $S_m$, $E_m$ and $I_m$. Counts are represented internally as fractions of $N_h$ to improve numerical conditioning, then returned as persons or mosquitoes. The force of infection links both age groups to one mosquito population. The sole child multiplier $q_c$ represents the product of the historical exposure and susceptibility labels; no separate mechanism is inferred.

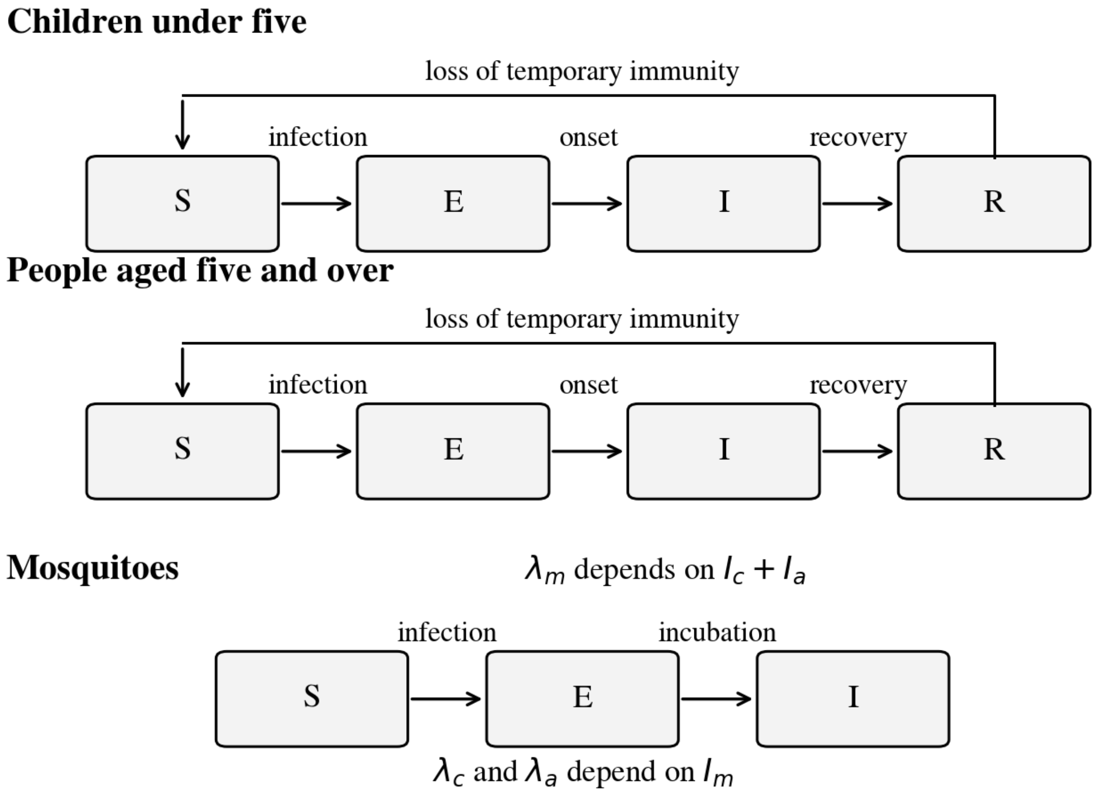


**Figure 2. Human SEIR and mosquito SEI structure. The forces of infection link both human groups to mosquitoes through Equations 2–3. Recruitment, natural death and disease-death flows appear in Equations 5–7; the vaccine acts on child infection and treatment on recovery.**

$$\lambda_c = q_c \beta(t) \frac{I_m}{N_h}[1 - v(t)], \ \lambda_a = \beta(t)\frac{I_m}{N_h} \quad (2)$$

$$\lambda_m = \beta_m \frac{I_c + I_a}{N_h}, \ \beta_0 = \frac{K}{M} \quad (3)$$

Human-to-mosquito infectivity is equal for the two infectious groups in the present model. The terms $\beta_0$ and $\beta_m$ are effective contact–transmission coefficients in the stated units; their values do not separate biting frequency from infection probability. The fitted composite K is the estimable human-transmission scale under the vector-density symmetry.

#### Seasonal forcing

$$\beta(t) = \beta_0 \max\{0.10, 1 + A[0.20\cos(2\pi(t - 115 - \varphi)/365) \quad (4a)$$

$$+ 0.80\cos(4\pi(t - 115 - \varphi)/365)]\} \quad (4b)$$

The period is 365 days, the anchor is day 115, and the annual and semiannual weights are 0.20 and 0.80. A and φ are fitted; the shape, anchor and floor are fixed assumptions. No rainfall time series is fitted. The floor remains explicit even though it is inactive within the fitted amplitude range. A seasonal fit does not by itself corroborate rainfall mechanisms or geography.

#### Population and disease dynamics

$$B_g = \mu_h \pi_g N_h + \delta_g I_g, \ \delta_g = \frac{CFR_g(\gamma_g + \mu_h)}{1 - CFR_g} \quad (5)$$

$$\frac{dS_g}{dt} = B_g + \omega_g R_g - \lambda_g S_g - \mu_h S_g \quad (6a)$$

$$\frac{dE_g}{dt} = \lambda_g S_g - (\sigma_h + \mu_h)E_g \quad (6b)$$

$$\frac{dI_g}{dt} = \sigma_h E_g - (\gamma_g + \mu_h + \delta_g)I_g \quad (6c)$$

$$\frac{dR_g}{dt} = \gamma_g I_g - (\omega_g + \mu_h)R_g \quad (6d)$$

These equations preserve $N_g = \pi_g N_h$ when initialised at the fixed shares. Disease deaths are compensated within the same virtual group. The disease-death hazard is derived so that the probability of disease death among competing exits from $I_g$ is exactly $CFR_g$. The assumed fatality input refers to that model exit process. Its empirical clinical denominator is not established.

$$\frac{dS_m}{dt} = \mu_{m,eff} M N_h - \lambda_m S_m - \mu_{m,eff} S_m \quad (7a)$$

$$\frac{dE_m}{dt} = \lambda_m S_m - (\sigma_m + \mu_{m,eff})E_m \quad (7b)$$

$$\frac{dI_m}{dt} = \sigma_m E_m - \mu_{m,eff} I_m \quad (7c)$$

Vector recruitment equals $\mu_{m,eff}$ M $N_h$, preserving total vector density. This convention makes survival changes visible without a separate mosquito ecology model. It is also the basis of the $\beta_0$–M confounding described in Section 10. It should be replaced by an empirically supported vector population process before entomological or density-reduction claims.

#### Vector control and treatment diagnostic

$$\kappa_j(c_j) = \kappa_{j,max}[1 - \exp(-k_j c_j)], \ j \in \{ITN, IRS\} \quad (8)$$

$$\mu_{m,eff} = \mu_m - \frac{\ln[(1 - \kappa_{ITN})(1 - \kappa_{IRS})]}{T_g} \quad (9)$$

Coverage is a fraction of the specified intervention target. ITN and IRS kill probabilities combine multiplicatively per gonotrophic cycle. Their maxima and saturation constants are illustrative inputs. The value k = 1.33 is not presented as an estimate fitted to Katureebe and colleagues: that observational study used different site-specific ownership, utilisation and burden measures [15]. Separate $k_{ITN}$ and $k_{IRS}$ inputs are retained, and linear and Hill alternatives are evaluated.

$$\gamma_g(c_{trt}) = \gamma_{g,0} + c_{trt}[\frac{1}{7} - \gamma_{g,0}] \quad (10)$$

The treatment diagnostic linearly interpolates baseline recovery to a seven-day mean duration. It does not reuse the vector saturation constant. Disease-death hazard is recomputed with Equation 5, leaving the infectious-exit fatality probability fixed. Treatment-specific reductions in severity, admission and fatality, treatment eligibility and treatment resources are absent. Any increase in model episodes or deaths therefore describes this artificial recovery–immunity experiment and has no interpretation as clinical harm from effective treatment.

#### Aggregate vaccine mechanism

$$v(t) = \min(\frac{t}{90}, 1)\, c_{vax} f_{elig} \varepsilon_{avg}\,, \quad \varepsilon_{avg} = 0.610 \quad (11)$$

For $t \geq 0$, the vaccine effect ramps over 90 days. The eligible share $f_{elig}$ is 0.40 of children under five. At 70% coverage the plateau reduction in the aggregate child force of infection is 0.1708, or 17.08%. Coverage denotes completed-course coverage of that fixed eligible target. The 0.75/0.25 efficacy weights, the 0.40 floor and the 40% eligible fraction are illustrative assumptions; they do not arise from observed dose completion or individual antibody kinetics [3,32].

The effect enters $\lambda_c$ before simulation. Child susceptible, exposed and infectious states therefore change, and reduced human infectivity can feed back through mosquitoes to both age groups. The implementation retains all caller parameters and passes treatment enhancement through the same scenario routine when a diagnostic combination is requested. There is no separate vaccine branch that drops an intervention or rebuilds default biological inputs.

This is a mean-field leaky-protection approximation across the child group, without vaccinated compartments, age progression, dose timing, waning trajectories, births into eligibility or boosters. The nominal force-of-infection reduction is not the realised reduction in reported child episodes: susceptible-pool and immunity feedback can attenuate or amplify the latter. A cohort and dose model is necessary for programme impact claims. Unused booster and efficacy-half-life fields are omitted from the reference implementation.

#### Reproduction and seasonal invasion

The unforced disease-free system uses infected-state order ($E_c$,$I_c$,$E_a$,$I_a$,$E_m$,$I_m$). $R_0$ is the spectral radius of $FV^{-1}$ [9,10]. For zero vaccine or its fixed plateau effect, let $q_c^* = q_c[1 - c_{vax} f_{elig} \varepsilon_{avg}]$. The correct shared-vector closed form is Equation 12; it adds the age-specific cycle contributions inside the square root.

$$R_0{}^2 = \frac{K\,\beta_m \sigma_h \sigma_m}{(\sigma_h + \mu_h)(\sigma_m + \mu_{m,eff})\mu_{m,eff}} \times \quad (12a)$$

$$[\frac{\pi_c q_c{}^*}{\gamma_c + \mu_h + \delta_c} + \frac{\pi_a}{\gamma_a + \mu_h + \delta_a}] \quad (12b)$$

At the fitted baseline, the closed form is 6.378679572 and the matrix calculation is 6.378679572. The arithmetic weighted mean of group-specific square roots is not used. The quantities for each age group are components of the shared transmission cycle, not independent epidemics. No endemicity category is assigned from this index.

The seasonal invasion diagnostic integrates the six-state linearised system over a full 365-day cycle and obtains its dominant Floquet multiplier. The baseline multiplier is 1.303e+22, with log-growth rate 0.139511 per day. Both indicate invasion in the assumed disease-free seasonal system. This multiplier is not labelled a periodic $R_0$. The 90-day rollout is not a periodic regime; the vaccine seasonal diagnostic uses its fixed plateau effect.

##### Episodes deaths and numerical solution

$$P_{g,j} = \int_{t_j}^{t_{j+1}} \sigma_h E_g(t)\,dt\,, \quad Y_{g,j} = \rho_g P_{g,j} \quad (13)$$

$$D_g = \int_0^T \delta_g I_g(t)\,dt \quad (14)$$

P denotes model episode onsets and Y expected reported episodes. A person may contribute more than one episode during the horizon. Counts are not unique infected people. Disease deaths D are integrated from the ODE flow, not calculated by multiplying infection acquisitions by a fatality fraction. For constant infectious-exit CFR, $D_g = CFR_g[P_g - \{I_g(T) - I_g(0)\}]$; the terminal infectious stock explains why a simple CFR × onset total can differ over a finite interval.

The solver is classical fourth-order Runge–Kutta with maximum step 0.5 days. It shortens the last step to hit every observation boundary exactly. Cumulative episode and mortality states are integrated at the same RK4 stages as the compartments. There is no dropped endpoint, duplicate step-length multiplication, post-fit solver switch or stochastic scenario perturbation.

Baseline spin-up iterates phase-aligned 365-day cycles with zero additional intervention. Two successive maximum annual changes in normalised compartment states must be below $10^{-8}$; failure within 40 cycles raises an error. The fitted baseline converged in 4 years. All scenario states at t = 0 match that baseline. The 0.25-day comparison and an independently implemented DOP853 solver provide numerical checks in Appendix B.

#### 9 Parameters and provenance

Table 3 distinguishes fitted values, reference choices and illustrative biological or programme assumptions. Literature references identify relevant mechanisms and contexts; they are not used to assert that an unverified exact local parameter was measured. Every changed assumption is propagated through the same model, rather than altering a results cell after simulation.

**Table 3. Parameter dictionary and evidence basis**

| Input | Value or interval | Units | Basis |
|---|---|---|---|
| $N_h$; $\pi_c$; $\pi_a$ | 1,000,000; 0.17; 0.83 | persons; fractions | Fixed standardisation choices; maintained virtual group totals. |
| $K = \beta_0 M$ | 1.169942; fit 0.30–3.00 | $day^{-1}$ | Fitted composite; M is held at 3.5 solely as a reference scale. |
| A | 0.459532; fit 0–0.90 | dimensionless | Fitted seasonal amplitude. |
| φ | 44.435073; fit 0–120 | days | Fitted phase; anchor 115 and harmonic weights fixed. |
| ρ | 0.195138; fit 0.05–0.60 | probability | Fitted nominal-share reporting parameter; age ratio 1.51 assumed. |
| $\mu_h$ | $1/(68 \times 365)$ | $day^{-1}$ | Illustrative demographic turnover approximation; not measured age-specific mortality. |
| $\mu_m$; $\sigma_m$ | 1/10; 1/12 | $day^{-1}$ | Illustrative mosquito lifespan and incubation rates [14]. No local entomological fit. |
| $\sigma_h$ | 1/12 | $day^{-1}$ | Illustrative human incubation rate. |
| $\gamma_{c,0}$; $\gamma_{a,0}$ | 1/22; 1/16 | $day^{-1}$ | Illustrative age-specific recovery rates; relevant malaria natural-history work [11,12]. |
| $\omega_c$; $\omega_a$ | 1/90; 1/365 | $day^{-1}$ | Illustrative exponential loss of temporary immunity; not directly estimated from [13]. |
| $q_c$ | 2.31 | dimensionless | Single child risk multiplier; equivalent to historical product 1.40 × 1.65. |
| $\beta_m$; M | 0.35; 3.5 | $day^{-1}$; ratio | Assumed mosquito transmission and reference density. M is not separately fitted. |
| $CFR_c$; $CFR_a$ | 0.003; 0.0005 | I-exit probabilities | Assumed disease-fatality probabilities with unspecified empirical clinical denominator. |
| $\delta_g$ | $CFR_g(\gamma_g + \mu_h)/(1 - CFR_g)$ | $day^{-1}$ | Derived competing-risk disease-death hazard. |
| $\kappa_{ITN,max}$; $\kappa_{IRS,max}$ | 0.50; 0.60 | cycle kill probabilities | Illustrative maxima, not observed national coverage effects. |
| $k_{ITN}$; $k_{IRS}$ | 1.33; 1.33 | dimensionless | Illustrative saturation choices; alternatives k = 0.50 and 3.00, linear and Hill curves. No fitted Katureebe derivation. |
| $T_g$ | 3 | days | Assumed gonotrophic cycle. |

| Input | Value or interval | Units | Basis |
|---|---|---|---|
| Recovery diagnostic | $\gamma_g(c) = \gamma_{g,0} + c(1/7 - \gamma_{g,0})$ | $day^{-1}$ | Assumed interpolation; fixed exit fatality. No clinical allocation use. |
| $\varepsilon_1$; $\varepsilon_2$; $\varepsilon_{avg}$ | 0.68; 0.40; 0.610 | fractions | Standard-site trial estimate [3] plus assumed floor and 0.75/0.25 averaging. |
| Vaccine ramp; eligible share | 90; 0.40 | days; child fraction | Aggregate assumptions; target 68,000 children at 100% coverage. |
| ITN programme input | US$28,000 per 1 pp | 720-day package | One campaign; assumed maintained coverage. Hypothetical input, no empirical base price year. |
| IRS programme input | US$40,000 per 1 pp per round | programme round | Four rounds at days 0, 180, 360 and 540; hypothetical input. |
| Vaccine package input | US$18 per completed course | eligible child | Four-dose package assumption: US$11.96 commodity plus US$6.04 delivery; no verified current procurement price. |
| Illustrative remaining life | 86.7; 61.0 | years | Hypothetical child and older-group life losses; not an observed age-at-death distribution or WHO life-table extraction. |
| Health discount rate | 0.03; 0 sensitivity | $year^{-1}$ | Continuous discount convention for a secondary YLL proxy. WHO GHE uses 0% [34]. |

Exact biological rates and programme inputs are retained as transparent reference assumptions. They require empirical replacement before a policy analysis. The source studies demonstrate important heterogeneity: ownership, access, use, clinical disease and infection prevalence cannot be collapsed into one coverage-to-kill calibration [13,15]. Alternative curves are a structural sensitivity exercise, not a substitute for such evidence.

### 10 Calibration and identifiability

#### Fitting criterion and search

$$L(\theta) = 0.50\,\mathrm{NMSE}_{\mathrm{total}} + 0.35(1 - r_{\mathrm{total}}) + 0.15\,\mathrm{NMSE}_{\mathrm{child}} \quad (15)$$

$\theta = (K, A, \varphi, \rho)$. NMSE is the mean squared error divided by the variance of the corresponding target series; r is Pearson correlation. This is a deterministic composite criterion, not a likelihood. Total and child terms are jointly weighted, while older-group error is reported separately. The weights are fixed retrospective modelling choices and have no interpretation as measurement-error probabilities.

Differential evolution uses best1bin, population size 12 per parameter, mutation range 0.5–1.0, recombination 0.7, tolerance $10^{-7}$, at most 160 generations and a final local polish [16]. Five runs use seeds 42, 1, 7, 123 and 2024. The parameter dictionary in Table 3 supplies the bounds. Every objective evaluation and every final metric uses the same periodic baseline, 0.5-day solver, calendar intervals and derived reporting probabilities.

**Table 4. Reconciled calibration across five search seeds**

| Seed | K | A | φ days | ρ | Loss | Total $R^2$ |
|---|---|---|---|---|---|---|
| 42 | 1.169891 | 0.459524 | 44.4344 | 0.195139 | 0.05286634 | 0.953706 |
| 1 | 1.169942 | 0.459532 | 44.4351 | 0.195138 | 0.05286634 | 0.953707 |
| 7 | 1.169867 | 0.459510 | 44.4353 | 0.195139 | 0.05286634 | 0.953706 |
| 123 | 1.169784 | 0.459492 | 44.4338 | 0.195139 | 0.05286634 | 0.953706 |
| 2024 | 1.169985 | 0.459540 | 44.4353 | 0.195138 | 0.05286634 | 0.953707 |

All five runs terminated successfully. The minimum-loss run is used in the reference scenarios; fitted output is not recomputed through an alternative reporting path. Seed agreement describes this numerical criterion and does not establish parameter uniqueness or empirical validity. The fit is conditional on fixed natural-history, population and observation assumptions.

**Table 5. Fit diagnostics on the unchanged historical synthetic target**

| Series | $R^2$ | RMSE per month | Pearson r |
|---|---|---|---|
| Total | 0.953707 | 379.66 | 0.987025 |
| Children under five | 0.832143 | 244.61 | 0.968707 |
| People aged five and over | 0.837372 | 482.82 | 0.987993 |

The total fit is stronger than the two subgroup fits because the composite objective and structural amendments affect the age pattern. The historical fixture was generated with another incidence definition and population convention. Its original generating values are therefore not used to claim successful recovery in this refit. Correlations and $R^2$ are descriptive; no nominal significance tests are attached to dependent monthly observations.

#### Structural and practical identifiability

Writing mosquito states as fractions of M $N_h$ removes M from the vector fractional equations. Human infection then depends on $\beta_0 M = K$. Scaling M and dividing $\beta_0$ by the same factor leaves the human trajectories invariant under matched vector initialisation. The reference implementation therefore fits K and fixes M = 3.5 as a scale. It neither reports separate $\beta_0$ or M recovery nor treats the fitted K as a unique empirical transmission coefficient.

Likewise, the historical child factors $\alpha_c$ and $\varphi_c$ entered only through their product. The reconciled model uses $q_c = 2.31$ and records the algebraic equivalence. Exposure and susceptibility could be separated only with external information or an extended mechanism in which they act differently. The present two labels do not establish biological necessity or separate identification.

Nine-point profiles for each fitted parameter fix that value and reoptimise the remaining three with the same composite objective. The profile values and full settings are retained in the source package. They illustrate conditional objective curvature. They are not likelihood-ratio confidence limits. Any future inferential fit should define an observation likelihood and perform an appropriate identifiability assessment with independent entomological and age-specific constraints [21].

The separate noiseless round-trip fixture uses generating $(K,A,\varphi,\rho) = (1.33,0.45,55,0.20)$. Differential evolution recovered (1.330000013,0.450000005,55.000000170,0.19999999999), with total monthly RMSE below 0.00001. This is an implementation consistency check using the same equations for generation and fitting. It is not evidence that field parameters can be recovered from surveillance observations.

### 11 Scenarios economics and optimisation

#### Scenario comparisons

All prevention scenarios use the fitted point, the same baseline t = 0 state and 720-day horizon. Coverages in Table 6 are intervention target fractions. ITN and IRS values are maintained assumed coverages; vaccine coverage refers to completed courses in the eligible target. The zero-addition baseline retains the stated recovery and background model assumptions.

$$\Delta Y(u) = Y(0) - Y(u), \; \Delta D(u) = D(0) - D(u) \quad (16)$$

**Table 6. Common-state reference scenarios over 720 days**

| Scenario | Reported episodes | Reported averted | ODE deaths | Deaths averted |
|---|---|---|---|---|
| Baseline | 528,041 | 0 | 3,612.40 | 0.00 |
| ITN 50% | 491,349 | 36,692 | 3,350.25 | 262.14 |
| ITN 80% | 469,619 | 58,422 | 3,198.48 | 413.92 |
| IRS 60% | 468,908 | 59,133 | 3,193.59 | 418.81 |
| Vaccine 70% | 525,918 | 2,123 | 3,589.27 | 23.13 |
| Vaccine 85% | 525,352 | 2,689 | 3,583.09 | 29.31 |
| ITN 50% plus vaccine 70% | 485,690 | 42,351 | 3,289.90 | 322.49 |
| ITN 50% IRS 40% vaccine 70% | 422,772 | 105,269 | 2,851.68 | 760.72 |

The baseline has 2,453,677 model onsets before reporting, of which 258,877 and 269,164 are reported child and older-group episodes. These are model expectations in a synthetic population. Table 6 mortality values come from integrated disease-death flows; they are not observed deaths.

The vaccine-only 70% scenario reduces reported episodes by 2,123 despite a 17.08% plateau change in the child force of infection. The difference reflects the aggregate susceptible and immunity feedback in this particular model. It should not be read as a field effectiveness estimate. The combination effect minus the sum of ITN 50% and vaccine 70% effects is 3,536 episodes. This interaction contrast is conditional and is not evidence of biological synergy or antagonism.

#### Recovery and immunity cycling diagnostic

With 70% recovery enhancement alone, the model produces 540,820 reported episodes, an increase of 12,779 relative to baseline. Infectious duration falls, but time spent in the infectious state is replaced by earlier recovery and subsequent loss of temporary immunity. Because fatality is held fixed, repeated episodes can also raise the integrated mortality output. This experiment omits the clinical mortality benefit of treatment.

The direction changes when immunity waning changes. Multiplying both waning rates by 0.5 yields 4,237 episodes averted; the reference rates yield −12,779 averted; doubling them yields −61,420 averted. Faster treatment is therefore not intrinsically harmful in the model; the observed cycling is conditional on the chosen recovery–immunity structure. Treatment diagnostic costs and economic ratios are not calculated.

#### Programme perspective and horizon

The economic component is an illustrative programme-budget calculation for additional prevention. It covers one 720-day package, compared with zero additional programme spending. Existing essential case management is outside the additional budget and is held in place. Household costs, productivity losses, treatment savings and health-system capacity costs are excluded. No societal cost-effectiveness or benefit–cost ratio is inferred.

Currency inputs are hypothetical US-dollar-denominated programme quantities assigned for this illustration. There is no measured base price year, Uganda procurement survey or inflation conversion. They are not reported as real 2022 or 2026 programme prices. Costs are not discounted within the 720-day package; all nominal rounds and course resources are counted. Any economic decision analysis would need priced resource use, a base year and discounted cash flows specified prospectively.

$$C(u) = 2{,}800{,}000\, c_{ITN} + 4(4{,}000{,}000) c_{IRS} + 18\, N_h \pi_c f_{elig} c_{vax} \quad (17)$$

The ITN input is US$28,000 per percentage point for one campaign with assumed coverage maintained throughout the horizon. IRS costs US$40,000 per percentage point per round, with four rounds at days 0, 180, 360 and 540. IRS therefore costs US$160,000 per percentage point over this package. A two-round sensitivity changes the cost schedule while retaining the stated maintained-effect assumption; it is a price-frequency scenario, not a mechanistic residual-effect model.

The vaccine target is $N_h \pi_c f_{elig}$ = 68,000 children, the same fixed population used in the protection calculation. At US$18 per completed four-dose course, one percentage point costs US$12,240 and 70% coverage costs US$856,800. The course input contains US$11.96 commodity and US$6.04 delivery as illustrative components. Eligibility is not inferred from life expectancy or an annual-birth proxy. No later birth cohort, wastage, dropout or repeat course is costed or protected.

**Table 7. Illustrative prevention package costs and average episode ratios**

| Package | Cost US$ | Reported averted | US$ per reported episode averted |
|---|---|---|---|
| ITN 50% | 1,400,000 | 36,692 | 38.16 |
| ITN 80% | 2,240,000 | 58,422 | 38.34 |

| Package | Cost US$ | Reported averted | US$ per reported episode averted |
|---|---|---|---|
| IRS 60% | 9,600,000 | 59,133 | 162.35 |
| Vaccine 70% | 856,800 | 2,123 | 403.60 |
| ITN 50% plus vaccine 70% | 2,256,800 | 42,351 | 53.29 |
| ITN 50% IRS 40% vaccine 70% | 8,656,800 | 105,269 | 82.24 |

The ratios in Table 7 are average ratios against the zero-addition comparator. They are defined only for positive benefit and do not establish an incremental cost-effectiveness frontier. Portfolios with different costs require incremental cost and effect differences and dominance assessment before decision use. A negative benefit is not assigned a favourable ratio. No GDP-per-capita threshold, per-case willingness-to-pay rule or claimed WHO adoption criterion is applied [23,33].

#### Mortality and health loss

$$w_g = \frac{1 - \exp(-rL_g)}{r}, \quad \Delta H = w_c \Delta D_c + w_a \Delta D_a \quad (18)$$

A secondary discounted YLL proxy uses assumed remaining lifetimes $L_c$ = 86.7 and $L_a$ = 61.0 years with continuous r = 0.03. The resulting weights are 30.860022 and 27.986214 years per model disease death. At r = 0 the limits are $L_c$ and $L_a$. These weights are correctly calculated from the stated assumptions; they are not drawn from an observed age-at-death distribution. The two broad age groups are insufficient to calculate standard age-specific burden without additional data.

WHO Global Health Estimates use a standard reference loss function with no time discount or unequal age weighting [34]. That convention differs from this illustrative discounted health proxy. The protocol therefore does not call ΔH a complete DALY, a WHO-standard YLL estimate or a validated national burden. No morbidity disability weights or sequela durations are modelled. The US$2 million reference allocation averts approximately 372.85 model deaths and 11,289.46 discounted proxy years; corresponding ratios are descriptive assumptions rather than adoption rules.

The previous flat illness-cost inputs are not used. Orem and colleagues studied Uganda with a historical cost basis [30]; an unverified US CPI conversion cannot establish a present local cost. Snyman and colleagues provide care-specific societal costs in constant 2022 US dollars, including US$19.02 per confirmed outpatient episode and US$29.29 per confirmed inpatient episode [31]. These observations have explicit care definitions and cannot be multiplied by all model onsets without an admission and confirmation mapping. They are retained as evidence for future resource collection, not silently substituted into the current budget.

#### Constrained prevention search

$$\max_u \Delta Y(u), \quad C(u) \le B, \quad 0 \le c_{ITN}, c_{IRS} \le 0.80, \quad 0 \le c_{vax} \le 0.90 \quad (19)$$

The primary hypothetical budget is B = US$2,000,000. The prevention decision variables are ITN, IRS and optional vaccine coverage; recovery enhancement is fixed at zero additional change. A stick-breaking expenditure parameterisation preserves feasibility and assigns the remaining budget to the final intervention up to its cap. The search focuses on budget-exhausting boundaries or portfolios limited by the final cap. It does not prove that all interior portfolios have been globally examined.

Differential evolution uses population size 12 per search dimension, at most 100 generations, tolerance $10^{-7}$ and local polish. Five seeds are run for the vector-only and vaccine-inclusive searches. The same 0.5-day solver evaluates both candidate and final output. A 101 × 101 grid in expenditure coordinates, including boundaries, is evaluated independently of search randomness. The zero-addition comparator and single-intervention boundaries remain explicit benchmarks.

**Table 8. Vaccine-inclusive US$2 million search across five seeds**

| Seed | ITN coverage | IRS coverage | Vaccine coverage | Reported averted | Cost US$ |
|---|---|---|---|---|---|
| 42 | 71.4286% | 0.0000% | 0.0000% | 52,482.83 | 2,000,000 |
| 1 | 71.4286% | 0.0000% | 0.0000% | 52,482.83 | 2,000,000 |
| 7 | 71.4286% | 0.0000% | 0.0000% | 52,482.83 | 2,000,000 |
| 123 | 71.4286% | 0.0000% | 0.0000% | 52,482.83 | 2,000,000 |
| 2024 | 71.4286% | 0.0000% | 0.0000% | 52,482.83 | 2,000,000 |

All five runs converge to the same corner to the displayed precision. The grid boundary has ITN coverage 71.428571%, IRS 0% and vaccine 0%, costing US$2 million and averting 52,482.83 reported episodes. The searched coverage fractions are not budget shares: this reference corner assigns 100% of additional programme spending to ITNs. Vector-only searches and the true-onset, death and discounted-proxy objectives also select this corner under the base assumptions.

The result is driven by the hypothetical cost horizon and transmission mechanism. Four recurring IRS rounds are substantially more costly than the single net campaign; mean-field vaccine protection is partly offset by the model's immunity and susceptible feedback. A zero selected vaccine addition is not evidence against the Ugandan vaccination programme. Clinical efficacy, equity, feasibility and real programme resources lie outside this optimisation. No unique global optimum or validated current allocation is claimed.

#### Planning boundary

This protocol evaluates a single fixed portfolio over 720 days. It contains no five-year optimal control claim. The historical annual planner's 15-percentage-point net floor, infection-dependent density rescaling, 12-percentage-point annual cap and campaign-years rule are not part of the reference analysis. Zero additional net coverage produces zero additional net kill effect. A future planner would require explicit delivery and decay states, recurrence costs, initial coverage and a dynamic objective before whole-path optimisation could be evaluated.

### 12 Uncertainty sensitivity and evaluation

#### Parameter sensitivity

One-at-a-time calculations vary 12 non-redundant inputs by ±20% and recompute the corrected unforced reproduction index, reported episodes and ODE deaths. Global sensitivity uses 500 Latin-hypercube samples over the same independent ±20% ranges, with seed 42 and the fixed reference scenario [17]. $\beta_0$ and M are not sampled as separately identified human-transmission factors; K is used instead. The complete input and output arrays are retained.

**Table 9. Selected partial rank correlations in the 500-sample sensitivity screen**

| Input | Unforced $R_0$ | Reported episodes | ODE deaths |
|---|---|---|---|
| K | 0.908 | 0.268 | 0.499 |
| $\mu_m$ | -0.958 | -0.293 | -0.624 |
| $\gamma_c$ | -0.640 | 0.453 | 0.862 |
| $\omega_c$ | 0.029 | 0.904 | 0.992 |
| $\omega_a$ | 0.018 | 0.944 | 0.935 |
| ρ | -0.038 | 0.989 | -0.050 |

PRCC is a conditional rank-based influence screen within the stated ranges, not a causal effect or probability of empirical importance. Local derivative signs were checked at 25 distributed sample points per input. Non-monotonic responses require caution, particularly for recovery rates in an immunity-cycling system. The exact unforced $R_0$ expression does not depend on reporting, immunity waning or seasonal amplitude; small sampled partial correlations for such inputs have no mechanistic interpretation. No multiple-testing p values are reported.

#### Assumed joint parameter ensemble

The ensemble comprises 300 draws with seed 20261011. K is lognormal with coefficient-scale 0.15; the perturbation of ρ is correlated −0.6 with the K log perturbation and bounded to 0.05–0.60. A and phase have bounded normal perturbations with standard deviations 0.08 and 8 days. Mosquito mortality has log-scale 0.10; recovery rates 0.15; waning rates 0.20; and $q_c$ 0.15. The common reference k has a bounded 25% perturbation, vaccine average efficacy a bounded 0.06 perturbation, and the reporting ratio a bounded 0.10 perturbation.

These joint distributions are illustrative assumptions. They are not a fitted posterior, empirical confidence region or formal propagation of calibration sampling error. The correlation is a specified sensitivity choice, not an estimated parameter covariance. M remains a scale, eligible share and age shares are fixed, and fatality probabilities are not sampled in this ensemble. Cost and intervention functional forms are evaluated in separate structural exercises.

Every draw is evaluated at each of 115 unique feasible portfolios formed from an 11 × 11 expenditure grid and the five main search solutions. The portfolio maximising mean reported benefit is compared with a candidate minimax-regret rule and the fitted-point selection. This is optimisation over a finite candidate pool, not proof of a continuous robust optimum. Maximum regret is defined relative only to the best portfolio in that same pool for each draw.

**Table 10. Full 300-draw finite-pool ensemble results**

| Quantity | 2.5th percentile | Median or mean | 97.5th percentile |
|---|---|---|---|
| Baseline reported episodes | 396,016 | 515,824 | 701,796 |
| Baseline ODE deaths | 2,830.85 | 3,521.08 | 4,468.17 |
| Selected-portfolio reported benefit | 19,559 | 53,105 | 93,156 |
| Selected-portfolio deaths averted | 148.64 | 370.76 | 650.43 |
| Mean selected reported benefit | Not applicable | 54,546 | Not applicable |

The expected-benefit and minimax candidate rules select the ITN corner to reported precision. The selected candidate is best within the examined pool at all 300 draws; its maximum candidate-relative regret is zero to numerical precision. This apparent stability concerns a restricted parameter distribution with fixed programme assumptions. It does not establish stability to omitted biology, prices, spatial heterogeneity or an empirically estimated vaccine mechanism. The percentiles are assumed-parameter intervals, not 95% confidence intervals for Uganda.

#### Repeated observation noise and search randomness

For each monthly multiplicative Gaussian noise level 5%, 10%, 15% and 20%, ten independent two-age target series are generated from the fitted deterministic expectation. Negative counts are floored at zero. Each target is recalibrated using seed 42 with at most 100 generations. Three allocation seeds, 42, 1 and 7, are run at that fitted point. The procedure yields 40 targets and 120 allocation searches, separating between-target noise variation from within-target allocation-search variation. Calibration-search randomness is not exhaustively sampled within each noise realisation.

**Table 11. Repeated monthly observation-noise diagnostics**

| Noise SD | Targets | Median total $R^2$ | Median ITN coverage | Largest coverage range |
|---|---|---|---|---|
| 5% | 10 | 0.847 | 71.4286% | <0.00001 pp |
| 10% | 10 | 0.581 | 71.4286% | <0.00001 pp |
| 15% | 10 | 0.410 | 71.4286% | <0.00001 pp |
| 20% | 10 | 0.305 | 71.4286% | <0.00001 pp |

Fit quality falls as synthetic noise rises, while the selected portfolio stays at the same base-cost boundary to displayed precision. Search-induced coverage differences within a fitted target are also below 0.00001 percentage points. A stable corner can reflect strong assumed relative prices and structural constraints even when parameters or observations are poorly determined. It should not be interpreted as evidence of reliable real-world allocation under noisy surveillance.

##### Structural and economic alternatives

The saturation constants are varied to 0.50 and 3.00. Linear kill fractions $\kappa_{max}$ c and Hill fractions $\kappa_{max}$ c/(0.5 + c) are also evaluated with the same mortality, solver and cost operators. At US$2 million, the linear and Hill searches use three seeds each. These are alternative mathematical response functions; no alternative is claimed to have been fitted to a Uganda intervention trial.

**Table 12. Selected structural and programme-cost alternatives**

| Assumption | IRS rounds | ITN coverage | IRS coverage | Reported averted |
|---|---|---|---|---|
| Exponential k = 0.50 | 4 | 71.43% | 0.00% | 19,092 |
| Exponential k = 1.33 | 4 | 71.43% | 0.00% | 52,483 |
| Exponential k = 3.00 | 4 | 71.43% | 0.00% | 98,865 |
| Linear kill fraction | 4 | 71.43% | 0.00% | 67,614 |
| Hill kill fraction | 4 | 71.43% | 0.00% | 49,109 |
| IRS half price and two rounds | 2 | 40.07% | 21.95% | 58,313 |

The structural curve changes materially affect the size of the model benefit even where the selected budget corner is unchanged. Cost changes can also change the portfolio: halving the IRS round input and using two rounds selects approximately 40.07% ITN and 21.95% IRS coverage. The complete half-price, double-price and two-/four-round runs are retained. This demonstrates why the base allocation cannot be treated as a generally optimal national intervention mix.

##### Evaluation and validation status

Mathematical evaluation includes NGM agreement, population identities, parameter symmetries and the exact competing-risk fatality relation. Numerical evaluation includes positivity, zero coverage, exact interval aggregation, step refinement and an independent DOP853 implementation. Regression checks verify caller-parameter preservation, treatment propagation with positive vaccine coverage and matched cohort costing. Appendix B provides the executed results.

The study has no independent human expert validation record or compatible external predictive evaluation. Synthetic recovery and code verification address implementation. Empirical validation would require clearly defined observed outcomes, independent time periods or locations, baseline comparators, reporting uncertainty and criteria fixed before inspecting evaluation results [26,27]. No present table is used to claim that those steps have been completed.

#### 13 Software and reproducibility

##### Reference implementation

The new reference package contains one C++17 RK4 core, a Python parameter and observation interface, ordered analysis routines, numerical assertions and result files. Scenario and fitting functions preserve the complete parameter vector. Coverage validation rejects non-finite or out-of-range input; interval validation rejects duplicate or non-finite boundaries; invalid solver steps are rejected. Runtime failures are surfaced instead of silently clipping negative populations.

The reference implementation is separate from the historical dashboard. Dashboard callbacks, deployment, conversational assistants, planner heuristics and browser performance were not repaired or retested by this protocol revision. The scientific tables here do not depend on a dashboard session or external language-model service. Historical source files are retained as context rather than declared to be the matching executable release.

##### Executed environment and files

The calculations used Python 3.12.14, NumPy 2.3.5, SciPy 1.17.0, Linux 6.18.44 on x86_64 with glibc 2.39, and a C++17 compiler. The main analysis run took 348.8 seconds in this environment; this is not a benchmark of the author’s original machine or browser. Exact relevant dependencies, compiler identification, seeds, input hashes and output hashes are recorded in the package.

The run scripts execute calibration, scenarios, search benchmarks, profiles, round-trip recovery, full finite-pool uncertainty, repeated noise, sensitivity and structural alternatives. Assertion checks and supplementary diagnostics use the same parameter and results directory. The complete reproduction entry point is documented in Appendix D. The package includes a licence for the newly written reference code; it does not relicense the historical dashboard or source data.

##### Availability and release scope

The supporting UMAPS_Reference_Analysis_v05.zip archive is retained with this revision and contains the matching source, unchanged synthetic target, parameter dictionary, numerical outputs, assertions and manifests. The clean protocol is supplied as an editable Word document and a matching PDF. The archive is a working reproducibility release; no public repository tag, OSF deposit, persistent DOI or retrospective registration entry is claimed. A public deposit requires verification of rights and a permanent identifier.

#### 14 Limitations

The reference analysis resolves the internal observation, population, incidence, mortality, vaccine-routing and time-horizon inconsistencies for this version. It still represents a small assumed system. The most material limits concern external observation compatibility, intervention evidence, natural history and programme resources, rather than numerical precision.

**Table 13. Remaining scientific limits on interpretation**

| Limit | Consequence | Required evidence or extension |
|---|---|---|
| Synthetic target and no independent observation set | Fit and same-model recovery do not establish field accuracy. | Governed surveillance outcomes, test and reporting information, and independent evaluation periods. |
| Maintained virtual age groups with no ageing | Age composition and vaccine eligibility are fixed, not projected. | Age progression, births, vaccination entry and observed demographic structure. |
| No asymptomatic, severe or diagnostic states | Survey infection, confirmed cases, admissions and clinical fatality cannot be matched directly. | Observation models and natural-history evidence for each intended outcome. |

| Limit | Consequence | Required evidence or extension |
| --- | --- | --- |
| Recovery-only treatment mechanism | Treatment mortality benefit and a valid clinical case-management allocation are absent. | Treatment-specific severity, eligibility, access, fatality and resource pathways. |
| Aggregate vaccine approximation | Dose completion, cohort protection and waning are not represented. | Dose and age cohorts with evidence-based waning and programme delivery. |
| Assumed vector kill curves and fixed vector total | Effects and relative benefit depend on uncertain mechanisms. | Local entomology, insecticide resistance, coverage use and intervention effect data. |
| Hypothetical programme prices and fixed horizon | Budget corner is conditional; no national cost-effectiveness recommendation follows. | Priced resource use, recurrence, base year, uncertainty, local opportunity costs and capacity. |
| Coarse assumed remaining lifetimes | Secondary health-loss proxy is not a complete DALY or standard age-specific YLL burden. | Age at death, normative life-table selection, disability and sequela inputs. |
| Finite parameter and candidate sets | Repeated-seed and ensemble stability do not prove a continuous global or robust optimum. | Broader structural scenarios, feasible interior searches and empirical joint uncertainty. |
| One homogeneous zone and no equity objective | District allocation, pregnancy, wealth and access disparities are unexamined. | Spatial and distributional models supported by appropriate data. |

These limits restrict the use of current results to explanation, debugging and conditional comparison. The model cannot justify reducing essential diagnosis or treatment. Its mean-field vaccine response and assumed prices should not override clinical trial evidence or the actual Ugandan vaccination programme. Future policy use requires replacement of the weakly supported inputs and independent assessment of the relevant outcomes.

### 15 Ethics governance and data protection

#### Current computational activity

The completed activity generated or analysed synthetic data and referred to publicly published aggregate information. It recruited no participants, contacted no patients and used no identifiable or individual-level health records. Participant consent and withdrawal procedures therefore do not apply to this computational activity. No ethics approval, exemption determination or national research registration is documented or claimed.

#### Future data governance

Any future access to restricted HMIS, DHIS2, individual or potentially re-identifiable data requires written decisions from the appropriate institution and data custodian about review, permissions and governance. Applicable Uganda research clearance and data-protection requirements must be assessed for the actual data and institutional arrangement [19,20]. Aggregate data alone do not establish exemption. Makerere’s committee is cited as an example of an institutional review body, without asserting affiliation or jurisdiction for this independent researcher [18].

Before restricted data are obtained, a governance plan must define authorised access, secure storage, retention, disposal, disclosure limits and responsibilities. No retention schedule or access agreement for future restricted data is invented here. Identifiable or restricted records must not enter external conversational assistants or services without an authorised governance arrangement.

##### Study responsibility and declarations

Jay Hiteshkumar Patel is the named investigator and is responsible for the study's scientific interpretation, source permissions and any submission. Funding, sponsor involvement and competing interests are not specified in the available author records, so this protocol makes no declaration that the study was unfunded or conflict-free. The intended journal and any additional contributor roles are likewise not documented.

AI assistance was used to reconcile equations, implement and execute the reference calculations, and edit and format the protocol. This work is not an independent clinical expert panel review, formal ethics decision or endorsement by an institution. The investigator remains responsible for checking the final content and for disclosing assistance under the receiving journal's applicable policy.

#### 16 Dissemination and future work

##### Dissemination

Dissemination comprises the clean retrospective protocol, the matching numerical reference package and any subsequently reconciled manuscript. A future public release should include the exact version, code licence, input and result manifests, supported data rights and a stable citation. Publication of an amended protocol should preserve the chronology and describe any change in estimands or results. The intention to use OSF in earlier records is not treated as proof of deposition.

##### Future scientific work

The next empirical study should first choose compatible clinical or infection outcomes and obtain their governed data. It should then specify the reporting and diagnostic observation process, use independently informed entomological constraints, and evaluate out-of-sample performance against simple baselines. Assessment criteria should be fixed before inspecting the evaluation period. The present retrospective work supplies a reproducible starting model, not that validation result.

Clinical case-management allocation requires a treatment severity and mortality mechanism. Programme vaccination analysis requires cohorts, dose completion, waning and matched costs. Economic evaluation requires resource quantities, local prices and base year, recurrent delivery, perspective and opportunity costs. Spatial and equity extensions should follow these prerequisites. No funded schedule or promised completion date is invented.

#### Reporting basis

The reporting basis is the Infectious Disease Modeling Reproducibility Checklist and its implementation framework [25,29], TRACE [28], ISPOR–SMDM transparency and validation guidance [26], and AdViSHE [27]. CHEERS 2022 informs the economic description [23]. These frameworks organise reporting and evaluation; they do not certify model validity by a score. Appendix C maps the present content and its specific limitations.

SPIRIT 2025 is designed for randomised trial protocols [22], and PRISMA-P for systematic-review protocols; neither is a compliance standard for this study. EPIFORGE applies if a future study develops and evaluates forecasts [24]. Current implementation counterfactuals are not forecasts. The protocol retains applicable modelling and economic reporting elements without asserting irrelevant trial or forecasting compliance.

### References

Numeric references are used throughout. Primary sources verified in the preceding source review and this revision are distinguished from illustrative assumptions in Table 3. Sources checked for this revision were accessed on 11 October 2026; no citation alone is treated as validation of an exact parameter.

### Appendix A Abbreviations and symbols

| Term | Meaning |
|---|---|
| UMAPS | Uganda malaria modelling framework named in the source project. |
| ACT; ITN; IRS | Artemisinin-based combination therapy; insecticide-treated net; indoor residual spraying. |
| HMIS; DHIS2; UMIS | Health Management Information System; District Health Information Software 2; Uganda Malaria Indicator Survey. |
| REC; UNCST | Research ethics committee; Uganda National Council for Science and Technology. |
| DE; NGM; RK4; LHS; PRCC | Differential evolution; next-generation matrix; fourth-order Runge–Kutta; Latin-hypercube sampling; partial rank correlation coefficient. |
| c; a; m | Children under five; people aged five and over; mosquitoes. |
| S; E; I; R | Susceptible; exposed; infectious; recovered states. |
| $N_h$; $N_g$; $\pi_g$; M | Standardised human total; fixed virtual group total; group share; vector-to-human reference ratio. |
| K; $\beta_0$; $\beta_m$ | Fitted $\beta_0$M composite; effective human-transmission coefficient; effective mosquito-transmission coefficient. |
| A; φ; $q_c$ | Seasonal amplitude; seasonal phase in days; single child risk multiplier. |
| λ; μ; σ; γ; ω; δ | Force of infection; natural mortality; incubation progression; recovery; loss of immunity; disease-death hazard. |
| ρ; $\rho_g$; $CFR_g$ | Nominal-share reporting input; age reporting probability; model infectious-exit disease-fatality probability. |
| κ; k; $T_g$ | Vector kill probability; coverage saturation input; gonotrophic cycle duration. |
| $c_{ITN}$; $c_{IRS}$; $c_{trt}$; $c_{vax}$ | Intervention target coverages; $c_{trt}$ denotes recovery enhancement in diagnostic runs only. |
| $f_{elig}$; $\varepsilon_{avg}$; v(t) | Eligible child fraction; assumed aggregate vaccine efficacy; force-of-infection reduction. |
| $P_{g,j}$; $Y_{g,j}$; $D_g$ | Model episode onsets; expected reported episodes; integrated ODE disease deaths. |
| $R_0$; Floquet multiplier | Unforced NGM reproduction index; annual seasonal linear-invasion multiplier. |
| C; B; ΔY; ΔD; ΔH | Illustrative programme cost; budget; reported episodes averted; ODE deaths averted; discounted YLL proxy averted. |
| YLL; YLD; DALY | Years of life lost; years lived with disability; disability-adjusted life year. No complete DALY is calculated. |
| pp; NMSE; RMSE; $R^2$ | Percentage point; normalised mean squared error; root mean squared error; coefficient of determination. |

### Appendix B Mathematical and numerical checks

The assertions below were executed against the fitted reference parameter vector on 11 October 2026. They assess the stated implementation and numerical behaviour. They are not a clinical validation or an independent expert endorsement. Full tolerances and exact values are retained in verification.json.

| No | Assertion | Executed result |
|---|---|---|
| 1 | Unforced NGM identity | Closed form and NGM: 6.378679572; difference below $10^{-10}$. |
| 2 | Intervention NGM identity | At ITN 50%, IRS 40%, recovery 70%, vaccine 70%: both 1.773962511. |
| 3 | Fixed group and vector totals | Maximum group or vector error below $1.4 \times 10^{-9}$ persons or mosquitoes. |
| 4 | Non-negative extreme trajectories | All tested states non-negative with half/double K and $\mu_m$ and high intervention coverage. |
| 5 | Zero allocation consistency | No-addition scenario and baseline reported totals agree exactly. |
| 6 | Vector-scale symmetry with fixed K | Doubling M at fixed K changes reported total by zero at displayed numerical precision. |
| 7 | Single child product equivalence | Rescaled historical factors give the same $q_c$ and human output. |
| 8 | Reporting propagation | Age probabilities preserve nominal average ρ and ratio 1.51. |
| 9 | Common baseline initial conditions | Maximum initial state difference is zero across tested scenarios. |
| 10 | Exact interval cumulative aggregation | One 720-day interval agrees with 24 × 30-day integrated totals. |
| 11 | Step-size convergence | 0.5 versus 0.25 days: relative reported-total difference $1.87 \times 10^{-9}$. |
| 12 | Independent DOP853 solver | Relative total difference $2.08 \times 10^{-9}$; maximum bin difference 0.000645 episode. |
| 13 | All caller parameters preserved | Custom waning, kill and efficacy inputs remain intact; input arrays are not mutated. |
| 14 | Treatment retained with positive vaccine coverage | Recovery enhancement changes vaccine-combination output; treatment is not dropped. |
| 15 | Competing-risk fatality identity | Derived infectious-exit disease-death probabilities equal 0.003 and 0.0005. |
| 16 | Horizon and eligible cohort costing | ITN 50% + IRS 40% + vaccine 70% costs US$8,656,800; eligible target 68,000. |
| 17 | Input rejection and budget feasibility | Five invalid input classes rejected; all main optimiser costs remain within US$2 million up to floating-point tolerance. |

### Appendix C Reporting crosswalk

This crosswalk identifies where the protocol reports the modelling and economic domains relevant to this study. An entry documents the corresponding content; it does not certify that an empirical input or validation requirement has been met.

| Framework domain | Protocol location | Reported content or limit |
|---|---|---|
| IDMRC model and software | 6–10; 13; Appendix D | Equations, units, parameter dictionary, solver, version, source map and dependencies. |
| IDMRC data and workflow | 5; 7; 10; Appendix D | Unchanged synthetic target, observation mapping, source roles, ordered analyses and hashes. |
| IDMRC reproducibility | 12–13; Appendices B and D | Executed calculations, seeds, assertions, source archive and stated public-release boundary. |
| TRACE purpose and concept | 3–6; 8–9 | Research questions, virtual population, model boundary and evidence status of inputs. |
| TRACE analysis and evaluation | 10–14; Appendix B | Calibration, scenarios, full finite-pool ensemble, noise repetitions, structural alternatives and checks. |
| ISPOR–SMDM and AdViSHE concept and inputs | 6–10; 14 | Assumption provenance and observation compatibility. Independent clinical expert assessment is absent. |
| ISPOR–SMDM and AdViSHE implementation and outcomes | 12–14; Appendix B | Independent solver and regression tests completed; external predictive evaluation remains outside present evidence. |
| CHEERS perspective and comparators | 6; 9; 11 | Additional programme-budget perspective, 720-day horizon, recurrence, cohort and hypothetical prices. No societal analysis or measured base year. |
| CHEERS effects and uncertainty | 11–12; 14 | Reported episodes, ODE deaths and labelled discounted proxy; cost/curve alternatives and assumed-parameter intervals. |
| CHEERS transparency and declarations | 13; 15–16 | Reproducibility and AI assistance disclosed; funding, conflicts and sponsor declarations not supplied by the author. |
| SPIRIT and PRISMA-P applicability | 16 | Randomised trial and systematic-review protocol standards are not claimed. |
| EPIFORGE applicability | 6; 16 | Current scenarios are counterfactual model experiments. No forecasting validation claim. |

### Appendix D Reproducibility package

The UMAPS_Reference_Analysis_v05.zip supporting archive identifies the reference analysis used for this protocol. Its contents are listed below. The historical dashboard is not the executable source of the current numerical tables. The source licence covers only the newly written reference routines.

| File or directory | Purpose |
|---|---|
| core.cpp | C++17 compartment derivatives, RK4, cumulative flows and phase-aligned baseline spin-up. |
| model.py | Complete parameter dictionary, validation, reporting propagation, scenario interface, NGM, Floquet and programme arithmetic. |
| run_analysis.py | Main calibration, scenarios, multi-seed optimisation, grid, profiles, round trip, ensemble, repeated noise and sensitivity. |
| checks.py | Seventeen numerical assertions, independent Python/DOP853 equations and alternative functional-form searches. |
| supplement.py | OAT, local derivative signs and exact feasible-vector ensemble diagnostics. |
| reproduce_all.py | Ordered entry point and output-hash verification for the complete reference workflow. |
| inputs/uganda_synthetic_monthly.csv | Unchanged 36-row historical target. Adult-labelled input means people aged five and over. |
| results/parameters.json; fitted_monthly.csv | Fitted parameter vector and corresponding targets and expected observations. |
| results/scenarios.json; optimisation.json | Common-state effects, full cost components, search seeds, coverage and expenditure shares. |
| results/ensemble_full.npz; assumed_parameters.csv | All portfolio-by-draw benefits and deaths, candidate vectors and 300 parameter draws. |
| results/noise_replicates.json; noise_summary.json | All 40 fitted noisy targets and 120 allocation searches, with between-target and within-target diagnostics. |
| results/lhs_full.npz; sensitivity.json; one_at_a_time.json | Complete global-sensitivity input/output arrays, PRCC and OAT response records. |
| results/verification.json; curve_alternatives.json | Executed assertions and alternative kill-curve outputs. |
| results/run_manifest.json; SHA256SUMS.json | Executed software environment, settings, seeds and file hashes. |
| requirements-lock.txt; environment.txt; README.md; LICENSE | Relevant package pins, compiler/platform record, reproduction instructions and reference-code licence. |
| authoring/build_protocol.py; authoring/assets/ | Document builder and two conceptual figure sources for the protocol. |

To regenerate the calculations, use Python 3.12 with the recorded dependencies and a C++17 compiler, then run python reproduce_all.py from the reference-code directory. The interface compiles the platform-specific core locally. Existing results can be checked with python reproduce_all.py --verify-existing. Generated binaries are not portable inputs. An exact rerun may produce tiny floating-point or search differences on another system; the documented tolerances and matching file hashes distinguish those from altered methods.

The historical synthetic input SHA-256 is:

```
e34443d00b7ffa618a0f2144f76c3c118905147ad6ca4d933b265724954f83db
```

The complete manifest records full source and result hashes rather than abbreviating their identity in the numerical tables. The new package keeps historical inputs intact and does not assert that original bulletin PDFs, a public DOI or the author’s unverified amended dashboard are included. Future deposits should cite this reference version and preserve the historical source chronology.

### Appendix E Amendments and deviations

| Date and version | Record | Consequences for the present protocol |
|---|---|---|
| 20 September 2026 Version 0.2 | Source-recorded earlier amendment; the original file was not available. | The project was described as retrospective. No prospective registration status is inferred. |
| 25 September 2026 Version 0.3 | Author's protocol records a manuscript v11 amendment pass. | Those claims remain historical. The accessible September dashboard is not declared the matching amended release. |
| 9 October 2026 Version 0.4 | Retrospective clarification of mathematical definitions, source roles and implementation differences. | Historical outputs and their limits were documented before the new reference analysis. |
| 11 October 2026 Version 0.5 | New reconciled reference equations and completed reanalysis. | K and $q_c$ replace separately unidentified factors; virtual age totals are maintained; infectious onsets, mortality flows and one reporting rule are used. |
| 11 October 2026 Version 0.5 | Common-state intervention and vaccine specification. | Scenarios preserve phase and baseline state; vaccine acts in $\lambda_c$; all parameters and recovery diagnostics propagate through one routine. Current effects supersede historical table values. |
| 11 October 2026 Version 0.5 | Programme and health-output specification. | 720-day prevention package counts recurrent IRS and the matched eligible vaccine stock. Unverified illness-cost inflation and adoption thresholds are removed; the discounted YLL proxy is labelled and calculated consistently. |
| 11 October 2026 Version 0.5 | Completed sensitivity and reproducibility calculations. | Five main seeds, a dense boundary grid, all 300 draws, 40 noise realisations, 500 LHS samples and 17 assertions are recorded with matching source and results. |
| Future version | Empirical or clinical extension requires new specifications and data. | District fitting, diagnostic/severity states, treatment fatality effects, vaccine cohorts, actual programme prices and external evaluation must be versioned and rerun rather than appended as completed claims. |

All analyses in version 0.5 were performed after access to the historical data and previous results. Changes in the incidence definition, demography, initial-condition estimand, vaccine mechanism and cost horizon alter numerical outcomes. They are substantive model amendments, not corrections to a printed decimal. The present protocol identifies the resulting reference study without rewriting the earlier source records or claiming prospective approval.
